# Genome-wide association study of thyroid-stimulating hormone highlights new genes, pathways and associations with thyroid disease susceptibility and age-of-onset

**DOI:** 10.1101/2022.12.22.22283779

**Authors:** Alexander T Williams, Jing Chen, Kayesha Coley, Chiara Batini, Abril Izquierdo, Richard Packer, Erik Abner, David J Shepherd, Robert C Free, Edward J Hollox, Nigel J Brunskill, Ioanna Ntalla, Nicola Reeve, Christopher E Brightling, Laura Venn, Emma Adams, Catherine Bee, Susan Wallace, Manish Pareek, Anna L Hansell, Tõnu Esko, Estonian Biobank Research Team, William Hennah, Balasubramanya S Rao, Frank Dudbridge, Louise V Wain, Nick Shrine, Martin D Tobin, Catherine John

## Abstract

Thyroid hormones play a critical role in regulation of multiple physiological functions and thyroid dysfunction is associated with substantial morbidity. Electronic health records were used to undertake the largest genome-wide association study of thyroid-stimulating hormone (TSH) levels, with a total sample size of 247,107. We identified 158 novel signals, more than doubling the number of known associations with TSH, and implicating 112 putative causal genes, of which 78 were not previously implicated. For the first time, we demonstrate that a polygenic score for TSH was associated with TSH levels in all ancestries in UK Biobank, and strongly predicted age of onset of hypothyroidism and hyperthyroidism in European ancestry participants. We developed pathway-specific genetic risk scores for TSH levels and used these in phenome-wide association studies to identify potential consequences of pathway perturbation. Together, these findings demonstrate the potential utility of genetic associations to inform future therapeutics and risk prediction for thyroid diseases.

## Introduction

Thyroid hormones are essential for energy metabolism and act on almost all cells. Thyroid dysfunction is associated with secondary cardiovascular, mental health, ophthalmic and other disease(1). Hypothyroidism has a high prevalence(2) and is most commonly due to autoimmune (Hashimoto) thyroiditis, in areas where iodine intake is sufficient.(1) Hyperthyroidism, prevalence 0.2%-1.3%, is most commonly due to autoimmune (Graves) disease or toxic nodular goitre.(1) Ageing, diet (including iodine deficiency), smoking status, genetic susceptibility, ethnicity, and endocrine disruptors are risk factors for thyroid diseases; defining genetic variants, genes, proteins and pathways associated with hypothyroidism and hyperthyroidism will inform a deeper understanding of the mechanisms of thyroid disease and inform prevention and treatment strategies.

Genome-wide association studies (GWAS) of quantitative traits have been particularly powerful and successful in identifying new drug targets(3-5). Most genetic associations for thyroid disorders were discovered in GWAS of thyroid-stimulating hormone (TSH) levels, a sensitive marker of thyroid function which is suppressed when levels of thyroxine (T_4_) and triiodothyronine (T_3_) are high and elevated when T_4_ and T_3_ levels are low. The largest GWAS to date, including 119,715 participants, brought the number of known genetic signals for TSH to 99, highlighting associations with hypothyroidism, hyperthyroidism and thyroid cancer(6).

Electronic health records (EHR) are increasingly utilised in genomic studies(7, 8). In the UK, primary care EHR have been recorded prospectively for more than 25 years. TSH is frequently measured in primary care because thyroid disease may present with non-specific symptoms or be asymptomatic. Through harnessing such TSH measures, our study included 247,107 participants, more than doubling the size of the largest study to date, increasing the number of genetic signals for TSH from 99 to 260. Using these 260 signals we then (i) tested the association between TSH-associated variants and disease; (ii) fine-mapped signals through annotation-informed credible sets; (iii) applied a consensus-based framework to systematically investigate and identify putative causal genes, integrating eight locus-based or similarity-based criteria; (iv) developed and applied a polygenic score (PGS) for TSH to show associations with susceptibility and age-of-onset of thyroid disease; (v) applied phenome-wide association studies (PheWAS) to individual variants, the PGS, and molecular pathway-specific genetic risk scores (GRS). Through evidence from the above, we aimed to define putative causal genes, and provide new insights into the mechanistic pathways underlying thyroid disorders and their relationship to other long-term conditions to inform relevant drug therapies.

## Results

In UK Biobank and the EXCEED study we undertook GWAS with TSH levels, using an inverse normal transformation and adjusting for age, genotyping array, sex and the first 10 principal components of ancestry (**Online Methods**). Across the two studies, we analysed 53,518,359 genetic variants in 127,392 European ancestry (EUR) participants and subsequently meta-analysed these GWAS summary statistics with those from the independent European-ancestry populations represented in the analysis by Zhou et al,(6) bringing the total sample size to 247,107 participants.

### TSH association with 260 variants

Using annotation informed fine-mapping (**Online Methods**), and a genome-wide significance threshold of P<5×10^−8^, we identified 260 independent signals for TSH at 156 unique genomic loci, of which 158 signals at 78 genomic loci are new. In addition to reaching P<5×10^−8^ in the meta-analysis, 226 of the 260 signals (128 of the 158 novel signals) showed consistent direction of effect and P<0.01 in at least two out of three large datasets available to us (UK Biobank(9), the analysis by Zhou et al(6), and an independent analysis of 40,948 individuals in Estonian Biobank, EstBB(10)), whilst 239 out of 260 signals were consistent if using a threshold of P<0.05 (**Supplementary Table 1**).

The SNP heritability of TSH was 16.6% (95% CI: 12.5%, 20.7%); together the 260 signals explain 22.6% of the TSH SNP heritability (**Online Methods**). The median number of variants per 95% credible set was 3, and 167 (64%) of credible sets had a putative causal variant with a posterior probability of association >50%.

### Identification of putative causal genes and causal variants

To better understand the functional relevance of our signals, we undertook comprehensive variant-to-gene mapping by integrating evidence from eight methods: (i) the nearest gene to the sentinel variant; (ii) the gene with the highest polygenic priority score (PoPS)(11); identification of (iii) expression quantitative trait loci (eQTL) or (iv) protein quantitative trait loci (pQTL) within the credible sets; (v) proximity to a gene for a thyroid-associated Mendelian disease (±500kb); (vi) an annotation-informed credible set containing a missense/deleterious/damaging variant with a posterior probability of association >50%; (vii) identification of a rare variant (±500kb of a TSH sentinel) association with hypo-or hyperthyroidism using whole-exome(12) and whole-genome sequencing(13) resources; and (viii) proximity to a human ortholog of a mouse knockout gene with a thyroid-related phenotype (±500kb).

We identified 112 putative causal genes satisfying ≥2 criteria, of which 30 were supported by ≥3 criteria (**Figure 1, Supplementary Table 2**), and compared these to 67 genes implicated typically by a single criterion in previous studies (**Supplementary Table 3)**(6, 14).

**Figure 1:**
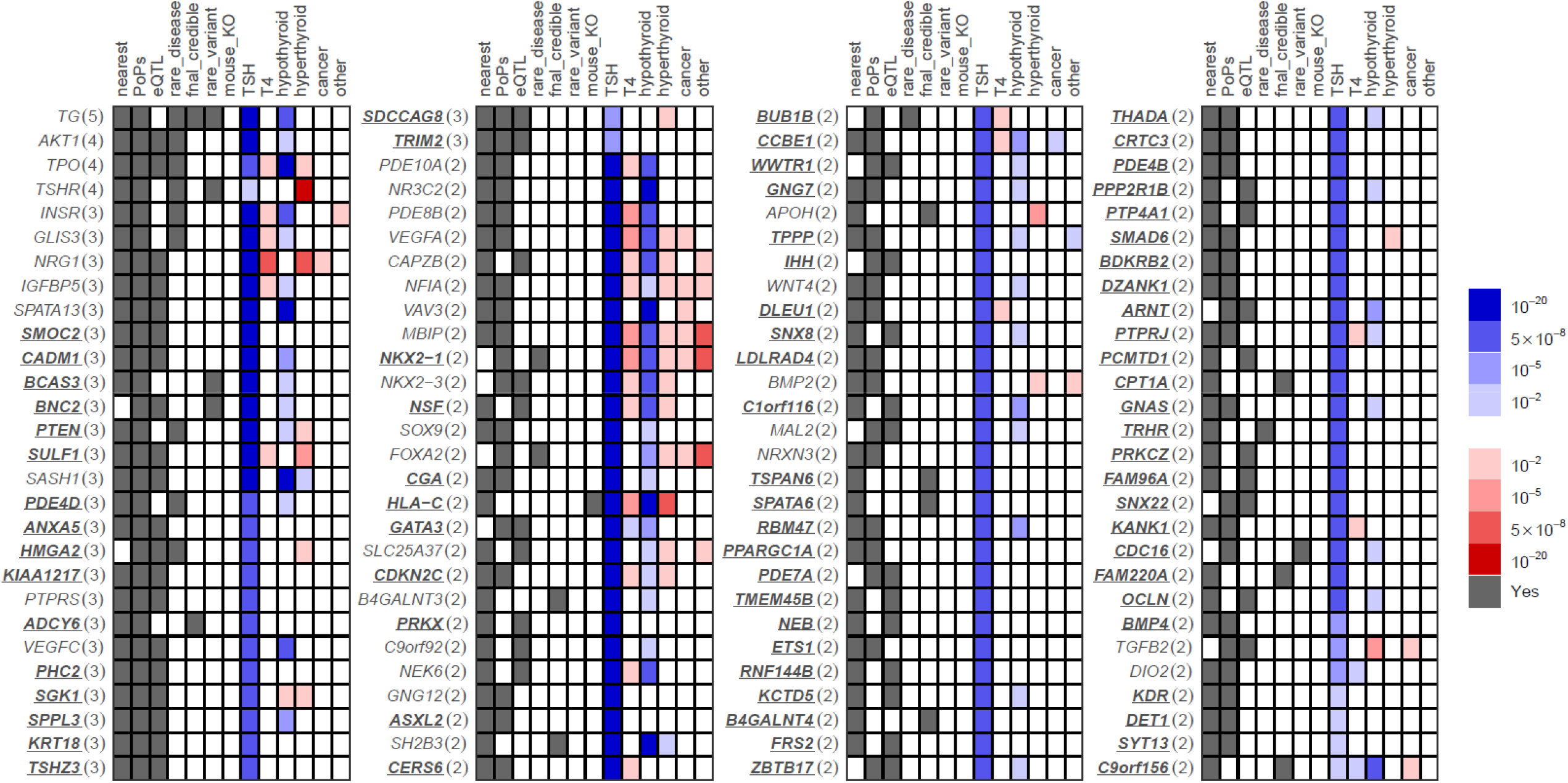
112 genes prioritised by two or more variant-to-gene criteria. The first seven columns indicate that at least one variant implicates the corresponding gene via the evidence for that column. The remaining six columns indicate the strength of association of the most significant variant implicating the corresponding gene as causal with respect to the TSH increasing allele, such that shades of blue represent associations with the other thyroid phenotypes that have the same direction of effect as the TSH association and shades of red represent an opposite direction of effect to the TSH association.

Of the 112 putative causal genes supported by ≥2 criteria, 78 genes have not been previously implicated in TSH level. The remaining genes were supported by additional criteria compared with the original reports, among which were 15 genes supported by additional signals (**Supplementary Table 2**). Among the 30 genes supported by ≥3 criteria, 18 were not previously implicated in TSH levels (***ADCY6, ANXA5, BCAS3, BNC2, CADM1, HMGA2, KIAA1217, KRT18, PDE4D, PHC2, PTEN, SDCCAG8, SGK1, SMOC2, SPPL3, SULF1, TRIM2, TSHZ3***, novel genes shown in bold) and 12 were previously reported genes supported by additional criteria compared with the original reports, among which were 6 genes also supported by additional novel signals (*TG, TSHR, GLIS3, IGFBP5, PTPRS, SPATA13*). The 30 genes supported by ≥3 criteria include genes involved in transcriptional regulation (***BNC2, HMGA2, PHC2, TSHZ3***, *GLIS3*), production, signalling or response to thyroid hormones (*TG, TPO, TSHR*, ***PDE4D***) or non-thyroid hormones (***ADCY6***, *INSR, NR3C2*), regulation of thyroid-relevant pathways (***HMGA2**, IGFBP5*), neuronal protection and neuropathies (***ADCY6, TRIM2***), angiogenesis (***SMOC2, SKG1***, *VEGFC, SPATA13*), AKT signalling (***PTEN, SGK1***, *AKT1, PTPRS*) and ciliogenesis (***SDCCAG8*)**.

To supplement understanding of the biological pathways and clinical phenotypes influenced by TSH-associated variants, we first tested associations with circulating free T_4_ levels, hypothyroidism, hyperthyroidism (**Figure 1, Supplementary Table 4**), thyroid cancer and other thyroid disease. Using DeepPheWAS(7), we then undertook PheWAS of selected individual variants which mapped to putative causal genes implicated by ≥3 criteria or by a single putative causal missense variant (posterior probability >50%; **Supplementary Figure 1**).

TSH signals implicating putative causal genes with ≥2 variant-to-gene mapping criteria show variable patterns of association with hypothyroidism, hyperthyroidism, thyroid cancer and other thyroid diseases. Among these are sentinel variants associated with hypothyroidism but not hyperthyroidism (implicating *AKT1, IGFBP5, INSR, GLIS3, SPATA13*, ***CADM1, BCAS3***, *SASH1*, ***PDE4D***, *VEGFC*, ***SPPL3, BNC2***, *PDE10A, PDE8B*, ***NR3C2***, *VAV3, SOX9, B4GALNT3*, ***CGA***, *C9orf92, NEK6*, ***NSF, CCBE1, GNG7, TPPP***, *WNT4*, ***SNX8, C1orf116, RBM47, KCTD5, PPP2R1B, PTPRJ, OCLN, C9orf156, GATA3, WWTR1***, *MAL2*, ***ZBTB17, ARNT, CDC16*)**, sentinels associated with hyperthyroidism but not hypothyroidism (implicating *TSHR, NRG1*, ***SDCCAG8, HMGA2***, *APOH, BMP2, SMAD6*), and sentinels associated with both hypothyroidism and hyperthyroidism with an opposite direction of effect (implicating *TPO*, ***PTEN***, *CAPZB, VEGFA, NFIA, MBIP*, ***HLA-C***, *SLC25A37, SLC25A37*, ***NKX2-1***, *NKX2-3, FOXA2*). However, tolerated *SH2B3* misssense variant, rs3184504 (allele T), associated with increased TSH, was associated with increased risk of both hypothyroidism and hyperthyroidism, and in our PheWAS with increased risk of other autoimmune disorders and pleiotropic associations with many traits (**Supplementary Tables 4 and 5**). Furthermore, the TSH-increasing allele of *SGK1* intronic SNP rs1743963, were associated with decreased risk of both hypothyroidism and hyperthyroidism and in our PheWAS, with increased calcium levels. *SGK1*, encoding serum glucocorticoid regulated kinase 1, is involved in regulation of ion channels and stress response.

Among relevant clinical phenotypes in the PheWAS is “secondary hypothyroidism”, defined by PheCode 244.1 which encompasses ICD codes for hypothyroidism due to surgery, ablation or medicaments used in treating hyperthyroidism. This code therefore represents consequences of treated hyperthyroidism (and is unrelated to central hypothyroidism, which was sometimes previously referred to as secondary hypothyroidism). We excluded these codes from our definition of hypothyroidism (**Figure 1**). When we explored single SNP associations with treated hyperthyroidism we found consistent directions of association with hyperthyroidism clinical codes and instances of associations, implicating genes *IGFBP5*, ***CADM1***, *SOX9, BMP2, TGFB2* and ***SYT13***, which were not detected by only studying hyperthyroidism codes in UK Biobank, consistent with earlier onset hyperthyroidism cases (**Supplementary Figure 2; Supplementary Table 6**).

Of the TSH sentinel variants implicating putative causal genes, a minority were associated with free T_4_ (*TPO, IGFBP5, INSR, GLIS3, NRG1, PDE10A, PDE8B, CAPZB, VEGFA, NFIA, MBIP*, ***HLA-C***, *NEK6*, ***CERS6, CCBE1, GNG7, PTPRJ, KANK1, C9orf156, NKX2-1***, *NKX2-3, GATA3*), and sentinels that were not associated with T_4_ included sentinels associated with hypothyroidism or hyperthyroidism (*TG, AKT1, TSHR, SPATA13*, ***CADM1, BCAS3***, *SASH1*, ***PTEN, PDE4D***, *VEGFC*, ***SGK1, SPPL3, SDCCAG8, HMGA2***, *NR3C2, VAV3*, ***CGA***, *SH2B3*, ***GNG7***, *APOH*, ***TPPP***, *WNT4*, ***SNX8***, *BMP2*, ***C1orf116, KCTD5, KDR***, *FOXA2*, ***WWTR1***, *TGFB2, MAL2*, ***ZBTB17, ARNT, CDC16***). Our findings suggest that the study of TSH levels is more sensitive approach to detecting genetic associations relevant to thyroid disease than the study of T_4_ levels.

Genes implicated by a single putative causal missense variant that was deleterious included ***SPATA6, ADCY6*** and *APOH*. Apolipoprotein H is involved in lipoprotein metabolism, coagulation and haemostasis. The G allele of the *APOH* missense deleterious variant, rs1801690 (minor allele frequency [MAF] 5.7% in EUR), was associated with reduced TSH, increased risk of hyperthyroidism, increased risk of congenital anomalies of endocrine glands and thyrotoxicosis in the UK Biobank DeepPheWAS analysis, as well as increased aspartate aminotransferase (AST) and alanine aminotransferase (AST) levels, increased height, reduced triglycerides, reduced carotid intima media thickness and, in FinnGen(15), reduced deep venous thrombosis risk. ***C9orf156*** (encoding TRNA Methyltransferase O) was implicated by a single putative causal tolerated missense variant, rs2282192 (T allele, frequency 28.8% in EUR), associated with increased TSH, increased hypothyroidism risk and decreased risk of nontoxic multinodular goitre and thyroid cancer as well as lower mean corpuscular volume and HbA1c.

Novel TSH signals associated with thyroid diseases also implicated relatively understudied putative causal genes, such as ***SPPL3*** and ***SDCCAG8. SPPL3*** encodes Signal Peptide Peptidase Like 3 involved in T cell receptor signalling, regulation of calcineurin-NFAT signalling and protein dephosphorylation. The ***SPPL3*** intronic variant rs2393717 G allele (frequency 47.3% in EUR) associated with increased TSH was associated with reduced hypothyroidism risk, increased tyrosine (a thyroid hormone precursor), as well as decreased C-reactive protein, increased insulin-like growth factor 1 (IGF-1), reduced height, whole body fat-free mass and reduced sex hormone binding globulin (especially in males), decreased gamma glutamyltransferase (GGT), increased alkaline phosphatase, reduced platelet count and eosinophils, increased cholesterol and with lipid composition traits. Mutations in ***SDCCAG8***, encoding the sonic hedgehog (SHH) signalling and ciliogenesis regulator, SDCCAG8, cause Bardet-Biedl Syndrome 16 (BBS16). Hypothyroidism and hyperthyroidism have been observed in commoner forms of Bardet-Biedl Syndrome(16). The TSH increasing allele, C (frequency 53.8% in EUR), of ***SDCCAG8*** intronic variant rs10926981 was associated with reduced risk of thyrotoxicosis and thyroid cancer, as well as reduced creatinine and increased eGFR.

As smoking is known to influence thyroid function, we tested whether adjustments for smoking attenuated sentinel SNP associations with TSH in UK Biobank. Adjustments for current smoking or pack-years resulted in betas <50% of unadjusted betas for eight SNPs, including variants implicating *DIO2, TSHR, C9orf156, DET1* and *DZANK1*.

### Druggable targets

For the 112 genes supported by ≥2 criteria, we surveyed gene-drug interactions using the Drug Gene Interaction Database (DGIDB). The protein products of these genes include targets for treatments to stimulate (thyrotropin [TSH]) or suppress (methimazole, targeting thyroid peroxidase, TPO) thyroid function, and drugs to treat thyroid cancer (e.g. the KDR inhibitor, vandetanib) as well as PDE4 inhibitors and AKT inhibitors utilised in immunoinflammatory conditions and cancers (**Supplementary Table 7**).

### Pathway analysis

Employing ConsensusPathDB(17), we tested biological pathways enrichment for the 112 putative causal genes supported by ≥2 criteria, highlighting signal transduction, particularly G protein (Reactome) and cAMP (KEGG) signalling, and the overlapping phosphodiesterases in neuronal function pathway (Wikipathways, including novel genes ***PDE4D, PDE7A, PDE4B, ADCY6***). The thyroxine production (Wikipathways) pathway included novel gene ***CGA***, encoding anterior pituitary glycoprotein hormones subunit alpha, which is common to TSH, chorionic gonadotropin (CG), luteinizing hormone (LH), and follicle stimulating hormone (FSH). New pathways of interest include VEGF hypoxia and angiogenesis (Biocarta), including ***ARNT, BDKRB2***, and ***KDR*** alongside *VEGFA* and *AKT1*, as well as opioid signalling, and platelet activation (**Supplementary Table 8**).

### Phenome-wide associations of pathway-specific TSH genetic risk scores

We hypothesised that partitioning a TSH genetic risk score (GRS) into pathway-specific GRSs according to the biological pathway(s) that each variant influences could inform understanding of mechanisms underlying TSH and thyroid disease, and possible consequences of pathway perturbation. Informed by the above prioritisation of putative causal genes and classification of these genes by pathway, we undertook PheWAS for TSH-weighted GRSs partitioned by each of 26 enriched pathways (FDR<5×10^−4^) after dropping redundant pathways (GRS correlation r^2^ <0.7).

We highlight examples of pathway-specific GRSs showing differing patterns of associations with thyroid and non-thyroid diseases (**Supplementary Table 9**). The GRS for higher TSH specific to the cAMP signalling pathway (KEGG, including novel genes ***GNAS, PDE4D, PDE4B, ADCY6, CGA***) was specific to increased risk of hypothyroidism; no associations (FDR<1%) with other traits were shown (**Supplementary Figure 3a**). A GRS for higher TSH specific to the activin receptor-like kinase (ALK) in cardiac myocytes pathway (Biocarta, including novel genes ***SMAD6, BMP4***) showed associations with reduced risk of nontoxic nodular and multinodular goitre, and simple goitre, as well as raised heel bone mineral density, standing height and whole-body fat-free mass and reduced FEV_1_/FVC (**Figure 2a**). GRSs specific to several pathways showed association to PheCode 244.1 capturing consequences of treated hyperthyroidism: the activin receptor-like kinase (ALK) in cardiac myocytes pathway, pathways in cancer (KEGG), factors and pathways affecting insulin-like growth factor (IGF1)-Akt signalling (Wikipathways), myometrial relaxation and contraction pathways (Wikipathways), FGFR3 signalling in chondrocyte proliferation and terminal differentiation (Wikipathways). The GRS for higher TSH specific to the platelet activation, signalling and aggregation pathway (Reactome, including novel genes ***GNG7, ANXA5, PRKCZ***) was associated with increased hypothyroidism risk, reduced nontoxic nodular goitre and simple goitre risk, raised urate, creatinine and reduced eGFR, reduced sex hormone binding globulin and testosterone, increased waist circumference, handgrip strength and whole body fat-free mass, osteochondropathies, increased triglyceride levels, and with lipid composition traits (**Figure 2b**).

**Figure 2:**
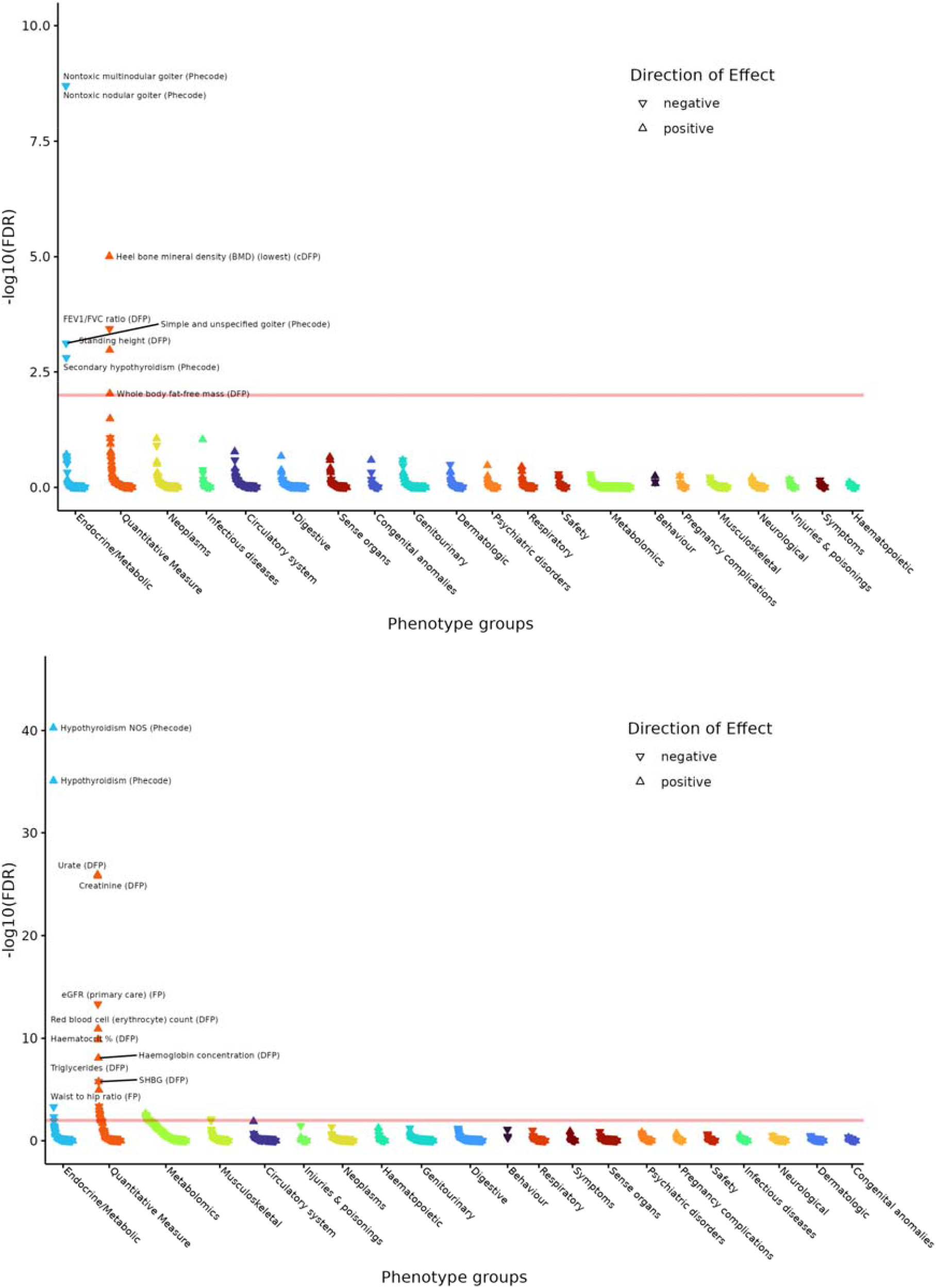
PheWAS for pathway-specific TSH-weighted GRS partitioned by: **(a, top)** activin receptor-like kinase (ALK) in cardiac myocytes pathway (Biocarta); **(b, bottom)** platelet activation, signalling and aggregation pathway (Reactome).

### Polygenic score associations

We constructed a polygenic score (PGS) for TSH using the summary statistics of approximately 1.12 million SNPs from our discovery GWAS (meta-analysed from UK Biobank, EXCEED and results from Zhou et al, with total sample size of 247,107 European-ancestry individuals) as the training dataset (**Online Methods, Figure 3: PGS performance across ancestries**).

**Figure 3:**
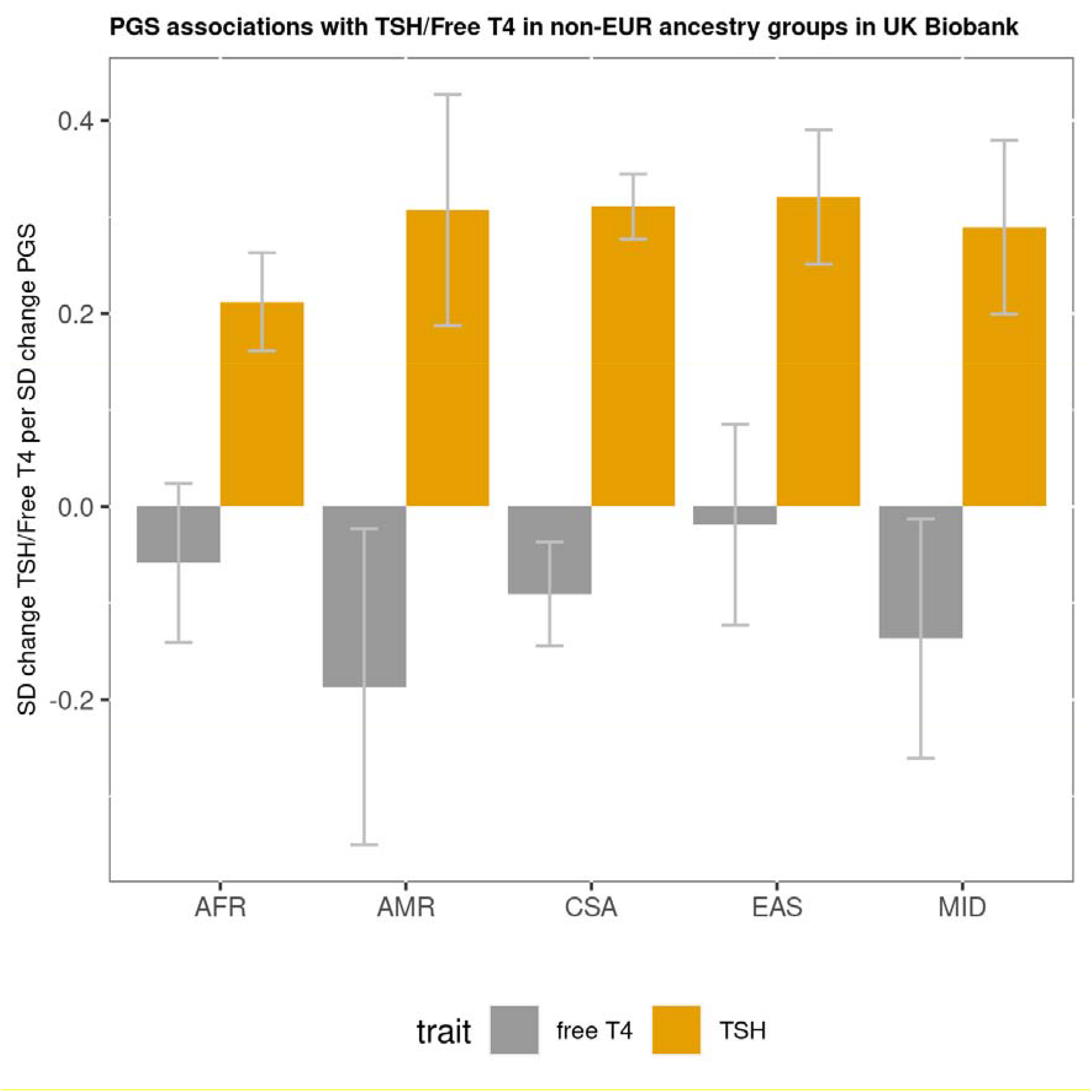
PGS performance across ancestries. Prediction performance of the TSH PGS across ancestry groups in UK Biobank shown as standard deviation (SD) change in TSH/free T4 per SD increase in the PGS. The ancestry groups were as defined by the Pan-UK Biobank initiative – AFR=African ancestry, AMR=admixed American ancestry, CSA=Central/South Asian ancestry, EAS=East Asian ancestry, MID=Middle Eastern ancestry. Error bars indicate 95% confidence intervals.

The TSH PGS showed distinct patterns of associations with relevant thyroid and non-thyroid phenotypes in our PheWAS (**Supplementary Figure 4**). Thyroid-relevant PGS associations included increased risk of hypothyroidism, lower risk of non-toxic (multi)nodular goitre, thyrotoxicosis, Graves’ disease, and thyroid cancer, and reduced tyrosine. Other PGS associations included increased FEV_1_/FVC, lower risk of chronic obstructive pulmonary disease (COPD), pneumonia, coeliac disease, common cancers and multi-site chronic pain, lower arterial stiffness, increased creatinine and urate, increased alkaline phosphatase and aspartate aminotransferase, increased eosinophils, decreased sex hormone-binding globulin, testosterone and IGF-1, decreased glucose (**Supplementary Table 10**) as well as altered lipid levels and composition. We found little or no attenuation of these PGS associations after adjustment for whether the individuals had ever smoked.

We then tested PGS associations across ancestries. Strong associations were shown with TSH levels in all ancestry groups tested (African, AFR; Admixed American, AMR; Central/South Asian, CSA; East Asian, EAS; Middle Eastern, MID; **Figure 3, Supplementary Table 11**). The TSH PGS was strongly associated with free T_4_ levels in E ancestry individuals (P=5.93×10^−58^), nominally associated with free T_4_ in the next largest ancestry group, CSA (P=0.0010, 1307 participants), and showed a consistent direction of effect in other ancestry groups (**Supplementary Table 11**).

To inform understanding of the relevance of the TSH PGS for disease, we subsequently tested disease susceptibility risk in all UK Biobank ancestry subgroups with at least 100 cases. In European ancestry UK Biobank participants the TSH PGS was associated with risk of hypothyroidism (P<1×10^−300^), hyperthyroidism (P=9.17×10^−166^), thyroid cancer (P=7.20×10^−10^) and other thyroid disease (P=1.35×10^−39^, **Supplementary Table 12**. In other ancestry groups, a consistent direction of association was shown with each of these traits, the largest case numbers being seen for hypothyroidism in CSA (862 cases, P=9.45×10^−21^, **Supplementary Table 12**). Results from a sensitivity analysis excluding individuals who were included in the discovery GWAS were consistent with these findings (**Supplementary Table 13**).

To further understand the clinical relevance of the polygenic score we examined risk of thyroid disease per decile of the PGS in European ancestry individuals. Individuals in the highest decile had 3.65-fold higher odds of hypothyroidism compared with those in the lowest decile, whilst those in the lowest decile had 4.21-fold and 2.18-fold higher odds of hyperthyroidism and thyroid cancer, respectively, and 3.40-fold higher odds of other thyroid disease compared with those in the highest decile (**Figure 4**). Given questions about how best to deploy and repeat testing for thyroid disease in asymptomatic patients and in patients with non-specific symptoms, we explored whether membership of a high or low risk decile for TSH PGS was associated with differences in age of onset of hypothyroidism or hyperthyroidism (**Online Methods**). Between individuals with median, highest and lowest deciles of the TSH PGS, clear differences were seen in age of onset of hypothyroidism (P<1.0×10^−300^, **Figure 5a**) and hyperthyroidism (P=4.30×10^−61^, **Figure 5b**). For example, a 5% prevalence of hypothyroidism was reached by age 51.1 years in the highest TSH PGS decile versus by 74.7 years in the lowest TSH PGS decile. Similarly a 1% prevalence of hyperthyroidism was reached by age 47.3 years in the lowest PGS decile compared with 71.2 years in the highest TSH PGS decile. Results from sensitivity analyses excluding individuals who were included in the discovery GWAS were consistent with the findings described here (**Supplementary Figures 5 and 6**).

**Figure 4:**
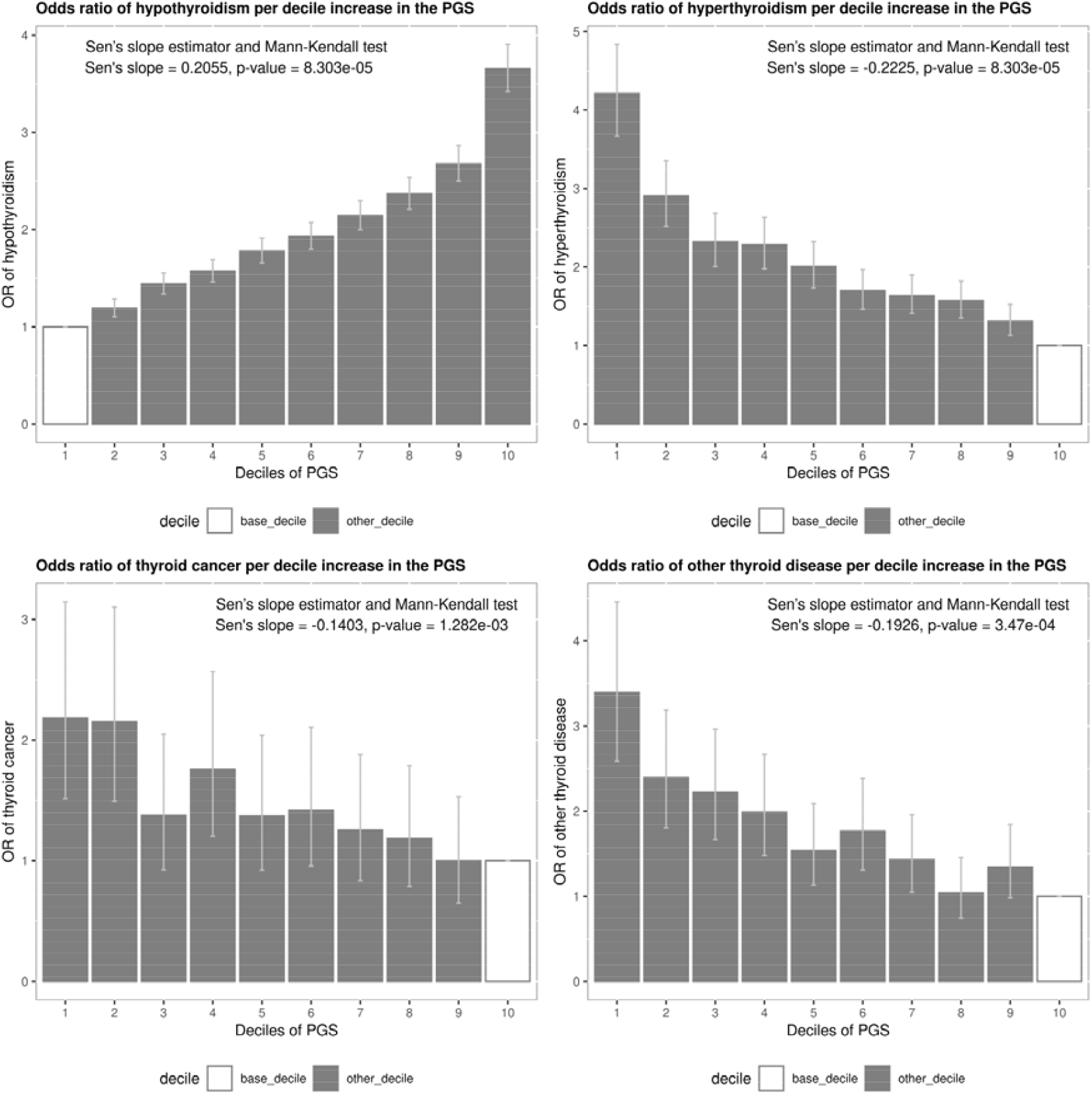
PGS performance across clinical diseases. Prediction performance of the TSH PGS for four clinical thyroid phenotypes – **(a, top left)** hypothyroidism, **(b, top right)** hyperthyroidism, **(c, bottom left)** thyroid cancer, and **(d, bottom right)** other thyroid disease. Error bars indicate 95% confidence intervals. The Mann-Kendall test is a test for monotonic trend.

**Figure 5:**
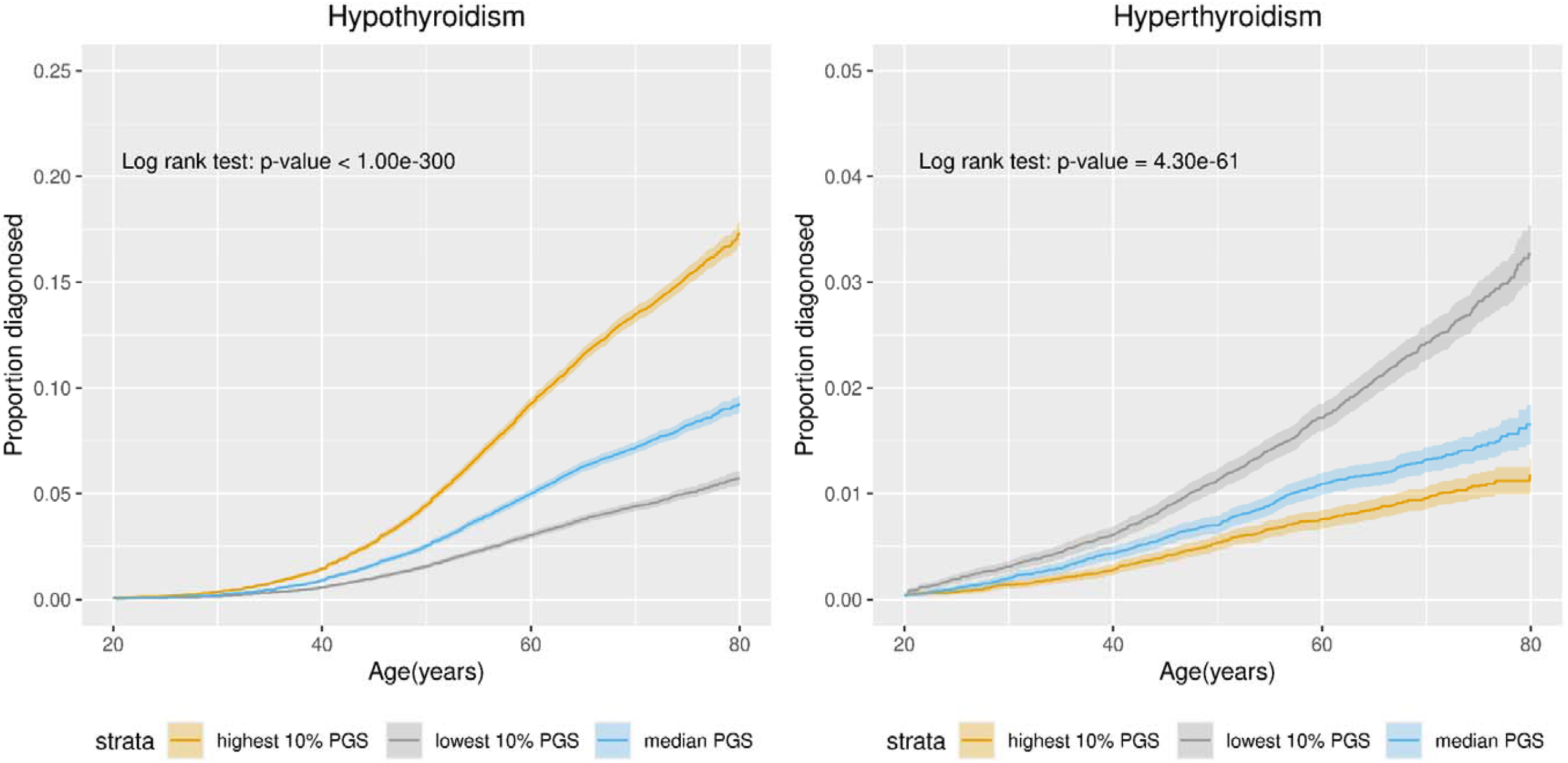
Age-of-onset analysis. Proportion of hypothyroidism **(a, left)** and hyperthyroidism **(b, right)** cases diagnosed by age stratified into lowest (grey), median (blue) and highest (yellow) decile for the TSH PGS. Shaded bands indicate 95% confidence intervals.

## Discussion

The large sample size of our study, achieved through utilising quality-controlled TSH measures from UK primary health care records, increased the yield of TSH genetic signals by over 2.5-fold, to 260. Through the most comprehensive initiative to identify putative causal variants and genes for TSH levels, we defined 112 high confidence genes implicated by multiple criteria. This is the first study to develop pathway-specific GRS for TSH levels and to use these in PheWAS, through our new DeepPheWAS platform(7), to investigate the potential consequences of intervening in relevant pathways. It is also the first to develop and use a polygenic score to predict age of onset of hypothyroidism and hyperthyroidism, showing marked differences in ages of onset of these conditions according to PGS deciles.

We implicate novel putative causal variants and genes, which alongside those previously reported(6, 14, 18), provide a more complete picture of relevant pathways and putative mechanisms. Pathways we highlight include signal transduction and cAMP signalling, as well as pathways not confidently implicated previously such as VEGF hypoxia and angiogenesis, AKT signalling and platelet activation. Our findings are consistent with signalling or response to thyroid or non-thyroid hormones (including IGF signalling), neuronal protection, angiogenesis and ciliogenesis influencing TSH levels and thyroid diseases.

Pleiotropic effects of aggregated TSH-associated variants have been previously shown through PheWAS. Partitioning TSH-associated variants by pathway provides a more nuanced understanding of the consequences of pathway perturbation on thyroid and non-thyroid disorders. We show contrasting patterns of phenotype association – for example highly specific associations for hypothyroidism (cAMP signalling) versus associations also with body composition, renal function and lipid traits (platelet activation pathways). As individuals may have high GRS for one or more pathways and low GRS for other pathways(19), individuals’ pathway GRS profiles may relate to patterns of comorbidities, and could have implications for treatment choices in thyroid diseases.

Here we adopted a powerful strategy for discovery of signals associated with thyroid diseases. We studied TSH as a quantitative measure within reference ranges, and detect novel signals which individually and in aggregate are associated with thyroid diseases. Not all TSH-associated variants showed association with free T_4_ levels, even those associated with thyroid disease, highlighting the value of TSH as a sensitive marker of thyroid disorders. In addition, thyroid function within the euthyroid range is associated with adverse outcomes(20) and thus TSH-associated variants that have not yet been overtly associated with thyroid disease remain highly relevant. GWAS of other quantitative traits – including those on the UK Biobank biomarker panel – have highlighted a number of targets leading to active drug development for related diseases(3-5). However, TSH has not yet been measured in UK Biobank samples. Thus, harnessing TSH levels measured in primary care in the EXCEED and UK Biobank studies more than doubled available sample sizes.

As with other contemporary genome-wide association meta-analysis, maximising power for discovery does not leave available datasets suitable for independent replication. In our study, 226 of the 260 independent sentinels (128 of the 158 novel signals) showed consistent direction of effect and P<0.01 in at least two of three large datasets (UK Biobank, EstBB, Zhou et al) in addition to reaching genome-wide significance in the meta-analysis. Not all SNPs were represented in the Zhou et al dataset due to contributing datasets using less dense imputation platforms than we were able to use in UK Biobank and EXCEED.

The PGS we developed for TSH utilised the full genome-wide association statistics to maximise predictive power. To our knowledge this is the first TSH PGS to be shown to be associated with TSH levels across all ethnic groups in UK Biobank. Power was more limited for testing other traits and diseases in the much smaller sample sizes available in non-European ancestries in UK Biobank. Nevertheless, we showed a consistent direction of association on T_4_ levels in all ancestries and association with hypothyroidism in South Asian participants (P=9.45×10^−21^, 862 cases). Understanding the genetic architecture of thyroid diseases within and across ancestries requires larger sample sizes in non-European ancestries, and an urgent global effort is required to include much more diverse populations in genomic studies than has been the case to date(21).

The PGS shows a strong association with age of onset of hypothyroidism and hyperthyroidism in European ancestry individuals. Universal screening for thyroid disease is not recommended(22, 23). Instead, case finding strategies are adapted to personal risk factors such as age, family history and relevant long-term conditions. Our findings raise the possibility of tailoring case finding strategies for thyroid disease according to a PGS for thyroid disease, especially if genome-wide data become available as part of the medical record. Further development and testing of a PGS would be required in independent, diverse populations, and alongside risk factors already employed in case finding.

Although we used TSH as the trait for signal discovery, there is potential for misclassification of thyroid diseases using electronic healthcare records in our investigating the clinical relevance of these signals. We undertook careful curation of clinical codes used to define clinical thyroid disease to test SNP associations. For example, we excluded cases of hypothyroidism resulting from treatment of hyperthyroidism. To avoid classifying cases incorrectly as hyperthyroidism or hypothyroidism we created a separate category “other thyroid disease” containing disorders such as goitre (where the clinical code was not explicit regarding thyroid status) and thyroiditis. In the latter group, we found that the direction of PGS associations were similar to those for hyperthyroidism, albeit with a smaller effect estimate.

In summary, we more than doubled the number of TSH-associated signals to 260, confidently implicated 112 priority genes, showed their relevance to thyroid diseases, and developed pathway-specific genetic risk scores which show differential patterns of pleiotropy of relevance in understanding co-morbidities and treatment choices. The PGS we developed predicts risk of onset and age of onset of hypothyroidism and hyperthyroidism, of potential utility in future case finding strategies, subject to further development and appropriate evaluation.

## Methods

### Cohort details

We analysed data from UK Biobank(24) and the Extended Cohort for E-health, Environment and DNA (EXCEED) study(25). UK Biobank is a cohort of approximately 500,000 individuals recruited from across the United Kingdom. Individuals aged between 40 and 69 years were recruited from the general population between 2006 and 2010. EXCEED recruited approximately 10,000 individuals primarily through local general practices in Leicester City, Leicestershire and Rutland. Recruitment started in 2013. Individuals invited to contribute to EXCEED were aged between 40 and 69 years. Both UK Biobank and EXCEED collected information at baseline concerning lifestyle and health outcomes, as well as providing linkage to electronic health records and genetic data.

### Phenotype

We captured all TSH results reported in the primary care data available in up to 230,000 individuals in UK Biobank and 8500 individuals in EXCEED utilising codes from Read version 2 and Read version 3 (Clinical Terms Version 3 or “CTV3”). We took an individual’s first non-missing TSH measurement to minimise the effect of thyroid function-altering medications on our phenotype as an individual is unlikely to have received these medications before their first thyroid function test. Where an individual’s first non-missing TSH measurement was 0, they were excluded from the analysis unless the individual also had a code in their primary care data for hyperthyroidism where a TSH measurement of 0 may be clinically feasible. In all other instances, we were unable to disentangle true 0 measurements from those that may have arisen due to, for example, an individual’s TSH being below the detectable range of the test apparatus used and, therefore, being entered into the primary care data as 0 or some other value such as “<0.05” which may have later been converted to 0. We excluded individuals with a TSH measurement <0.4 or >4.0 mU/L as has been done previously(14).

### Genome-wide association study

We applied an inverse normal transformation to the residuals from linear regression of the TSH phenotype against age (at time of measurement) and sex. This transformed phenotype was used for genome-wide association testing under an additive genetic model adjusted for age, genotyping array, sex and the first 10 principal components of ancestry using PLINK 2.0(26). We analysed individuals of European ancestry, as defined by the Pan-UK Biobank initiative(27), who were not more closely related than third-degree relatives using a KING software relatedness coefficient >0.0884 to indicate second-degree relatives or closer(28). In UK Biobank, we tested genetic variants with a minor allele count >20 and imputation score >0.5. In EXCEED, due to the smaller sample size, we tested genetic variants with a minor allele frequency >0.1% and imputation score >0.5.

We derived the LD Score regression intercept using LDSC(29) to estimate inflation in our test statistics due to confounding, such as by cryptic relatedness or population stratification. We estimated, separately, the LD Score regression intercept for the GWAS in UK Biobank and EXCEED. The UK Biobank test statistics were corrected for inflation (λ_LDSC_ = 1.05) prior to meta-analysis. The EXCEED test statistics were not corrected for inflation (λ_LDSC_ = 0.98).

We used METAL(30) to meta-analyse the results from the GWAS in UK Biobank and EXCEED and the previous largest GWAS of TSH(6). Since the EXCEED results were aligned to GRCh38, we ran LiftOver to map the results to GRCh37. Over 99.5% of the genetic variants tested in the GWAS in EXCEED were successfully mapped to GRCh37. Following meta-analysis, we estimated the LD Score regression intercept once more.

We estimated the proportion of variance explained by the sentinel SNPs using the formula:

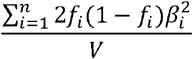

where *n* is the number of SNPs, *f*_*i*_ and *β*_*i*_ are the frequency and effect estimate of the *i*^*th*^ variant from our UK Biobank discovery analysis, and *V* is the phenotypic variance (always 1 as TSH was inverse-normal transformed).

### Signal selection

We selected 2 Mb loci centred on the most significant variant for all regions containing a variant with P<5×10^−8^. Loci within 500 kb of each other were merged for fine mapping.

PolyFun(31) and Sum of Single Effects (SuSiE)(32) was used to fine-map autosomal non-HLA loci utilising pre-computed functional prior causal probabilities based on a meta-analysis of 15 UK Biobank traits. The functional priors are proportional to per-SNP heritabilities estimated from the functional enrichments of 187 variant annotations from the baseline-LF 2.2.UKB model (Gazal S, 2018), including those relating to conservation, regulation, MAF and linkage disequilibrium, which were estimated using an extension of stratified-LDSC(33). Imputed genotype data from 10,000 randomly selected European individuals from UK Biobank was used as an LD reference. Loci for which PolyFun and SuSiE did not identify any credible sets, as well as HLA and chromosome X loci, were fine-mapped using the Wakefield method(34) with the prior *W* set as 0.04 in the approximate Bayes factor formula. 95% credible sets were generated for all loci, and variants with the highest posterior inclusion probability (PIP) per credible set identified (**Supplementary Table 14**).

We assessed whether signals were consistent (consistent direction of effect and P-value less than 0.01 or 0.05) in at least two of: (a) our results from UK Biobank, (b) summary statistics from the analysis by Zhou et al, and (c) an independent dataset from Estonian Biobank.

Estonian Biobank is a population-based biobank with 212,955 participants in the current data freeze (2022v2). All biobank participants have signed a broad informed consent form and information on ICD codes is obtained via regular linking with the national Health Insurance Fund and other relevant databases, with a majority of the electronic health records having been collected since 2004(10). The TSH concentrations were queried with LOINC code 3016-3 (“Thyrotropin in Serum or Plasma”, n=40,948), with values below 0.4mU/L and above 4.0 mU/L being excluded from the analysis. We used earliest possible measurement from each participant and analyses were restricted to individuals with European ancestry. Further details of genotyping and analysis are in the **Supplementary Note**.

### Novel signals

We searched PubMed and GWAS Catalog to identify applied studies focused on thyroid stimulating hormone and associations reaching P<5×10^−8^. These included the following sources: Gudmundsson *et al*. (35), Kwak *et al*. (36), Malinowski *et al*. (37), Medici *et al*. (38), Nielsen *et al*. (39), Popovic *et al*. (40), Porcu *et al*. (18), Taylor *et al*. (41), Teumer *et al*. (14), and Zhou *et al*. (6). We determined whether a signal was novel if its extent of linkage disequilibrium (LD) with nearby previously reported signals was <0.2 (R^2^).

### Epidemiological associations with clinical thyroid disease

We tested the association between our sentinel variants and free thyroxine (T_4_) or five clinical thyroid diseases: (i) hypothyroidism; (ii) hyperthyroidism; (iii) thyroid cancer; and (iv) other (non-cancer) thyroid disease. Using the UK Biobank primary care data, we extracted free T_4_ measurements that co-occurred with the corresponding individual’s first TSH measurement. To maximise potential cases for hypo- and hyperthyroidism, we utilised data available in primary care, secondary care and self-reported diagnoses (UK Biobank Data-Field 20002). For the remaining clinical disease phenotypes, we defined these using primary care alone (other thyroid disease), and cancer register data (thyroid cancer). To reduce the overlap in cases for the clinical disease phenotypes, we defined a case by their first diagnosis of hypothyroidism, hyperthyroidism, thyroid cancer and other thyroid diseases. The clinical codes used to define free T_4_ and the clinical thyroid diseases are presented in (**Supplementary Table 15)**. We used PLINK 2.0(26) to test the associations under an additive genetic model adjusted for sex, genotyping array, age at recruitment to UK Biobank and the first 10 principal components of ancestry.

These phenotypes were further tested for association with our polygenic score (described below).

To assess the robustness of the associations between our sentinel SNPs and TSH to smoking behaviour, we fit separate models that included current smoking status (UK Biobank data field 1239) and pack-years of smoking (UK Biobank data field 20161). We determined that an association was attenuated upon adjusting for smoking if the smoking-adjusted effect estimate was less than 50% its original magnitude.

### Identification of putative causal genes

To systematically prioritise putative causal genes for TSH associated signals, we integrated eight sources of evidence including: (i) the nearest gene to the sentinel variant; (ii) the gene with the highest polygenic priority score (PoPS)(11), a method based on the assumption that causal genes on different chromosomes share similar functional characteristics; identification of (iii) expression quantitative trait loci (eQTLs) or (iv) protein quantitative trait loci (pQTLs) within the credible sets; (v) proximity to a gene for a thyroid-associated Mendelian disease (±500kb); (vi) an annotation-informed credible set containing a missense/deleterious/damaging variant with a posterior probability of association >50%; (vii) identification of a rare variant (±500kb of a TSH sentinel) association with hypo- or hyperthyroidism using whole-exome(12) and whole-genome(13) sequencing resources; and (viii) proximity to a human ortholog of a mouse knockout gene with a thyroid-related phenotype (±500kb).

For (i), (v), (vii) and (viii), all 260 signals were used as input; for (ii), the 257 autosomal signals were used as input; for (iii), (iv) and (vi), all variants across the credible sets were used as input. Where there were two signals with the joint-highest posterior probability in their credible set, the one with the smallest P-value was used.

We catalogued previously reported genes (**Supplementary Table 3**) implicated by mapping genome-wide significant sentinels for thyroid traits using eQTL colocalization (P<1×10^−7^)(14) or DEPICT (FDR≤0.01, ref.(6)), to define whether the genes we implicated were novel.

#### Expression quantitative trait loci (eQTLs)

We used the SNP2GENE function implemented by FUMA(42) to facilitate the eQTL analysis. FUMA contains several eQTL datasets across a broad range of tissue types. We ran SNP2GENE requesting eQTL results from the GTEx v8 (thyroid, hypothalamus and pituitary tissues) and eQTLGen (blood, cis- and trans-eQTLs) datasets. We performed approximate colocalisation between our GWAS and eQTL signals by identifying whether the top variant in an eQTL signal was in one of our 95% credible sets (**Supplementary Table 16**).

#### Protein quantitative trait loci (pQTLs)

Two pQTL datasets were included in the pQTL analyses: deCODE Genetics(43), with data for 4,719 proteins measured by 4,907 aptamers, and the SCALLOP Consortium(44), including 90 cardiovascular proteins. The significance level for pQTL associations were set as in the original publications: P-value<1.8×10^−9^ for deCODE Genetics(43) and P-value<5×10^−8^ for the SCALLOP Consortium(44). We performed approximate colocalisation between our GWAS and pQTL signals by identifying whether the sentinel variant in a pQTL signal was in one of our 95% credible sets.

#### Polygenic priority score (PoPS)

We used a gene prioritization tool, PoPS(11), to calculate gene features enrichment based polygenic priority score(44) to predict genes for our TSH signals. The full set of gene features used in the analysis included 57,543 total features – 40,546 derived from gene expression data, 8,718 extracted from a protein-protein interaction network, and 8,479 based on pathway membership. In this study, we prioritized genes for all autosomal TSH signals within a 500kb (±250kb) window of the sentinel and reported the top prioritised genes in the region. If there was no gene prioritized within a 500kb window of the sentinel, we reported any top prioritized genes within a 1Mb window (**Supplementary Table 17**).

#### Nearby Mendelian disease genes

We selected rare Mendelian-disease genes from ORPHANET (https://www.orpha.net/) within ±500kb of a TSH sentinel that were associated with thyroid-related diseases. We implicated the gene if the string “thyro” (but not “parathyro”) was included in either the disease name or appeared frequently in human phenotype ontology (HPO) terms for that disease. We manually checked the diseases and HPO terms identified for relevance (**Supplementary Table 18**).

#### Nearby mouse knockout orthologs with thyroid related phenotype

We selected human orthologs of mouse knockout genes with thyroid related phenotypes, as listed in the International Mouse Phenotyping consortium (https://www.mousephenotype.org/) within ±500kb of a TSH sentinel. The thyroid related phenotypes included enlarged thyroid gland, abnormal thyroid gland morphology and increased/decreased circulating thyroxine level (**Supplementary Table 19**).

#### Functional annotation of credible sets

We annotated variants in the 95% credible sets using Variant Effect Predictor (VEP)(45). We implicated the gene if there was a variant with >50% posterior probability in the credible set that was also either a missense variant, annotated as “deleterious” by SIFT, annotated as “damaging” by PolyPhen-2 or had a CADD PHRED score ≥20.

### Rare variant analysis from whole exome and whole genome sequencing

We performed a lookup for rare variant associations with hypothyroidism or hyperthyroidism within ±500kb of a TSH sentinel using the following resources: (i) single variant and gene-based exonic associations from the AstraZeneca PheWAS Portal(12) (https://azphewas.com/); (ii) single variant whole-genome associations in 150,119 UK Biobank participants(13). For all tests, we used MAF<1% and P<5×10^−6^ (**Supplementary Table 20**).

### Pathway analysis

We used ConsensusPathDB(17) to test for enrichment of our prioritised genes in up to 31 pathway and gene set ontology databases. Pathways with FDR<5% are reported.

### Pathway-specific GRS

We selected 26 pathways that were enriched at FDR <5×10^−4^ for our 112 genes implicated by 2 or more lines of evidence (**Supplementary Table 8**). We created a weighted GRS (weights estimated from the TSH meta-analysis of UK Biobank and EXCEED) for each of the 26 pathways by including, for each gene in the pathway, the variant with the most significant P-value that implicates the gene in our variant-to-gene mapping (**Supplementary Table 2)**. Each of the 26 GRS were then checked for association with up to 1939 traits in the PheWAS.

### Polygenic score (PGS)

We applied PRS-CS-auto*(46)* to construct a polygenic score (PGS) using the summary statistics from our discovery GWAS (a meta-analysis of UK Biobank, EXCEED and results from Zhou et al (HUNT, Michigan Genomic Initiative [MGI], ThyroidOmics)) as the training dataset. PRS-CS-auto is a Bayesian approach, which automatically learns hyper-parameters from the training data; no validation dataset is required. We tested the association of this PGS trained from EUR ancestry group with TSH in non-EUR ancestry groups in UK Biobank, including AFR, AMR, CSA, EAS, MID. Associations were tested using a linear regression model, adjusted for genotyping array, age at TSH measurement, sex and the first 10 principal components of ancestry. We evaluated the association of the TSH PGS with susceptibility to hypothyroidism, hyperthyroidism, thyroid cancer, other thyroid diseases and thyroid eye disease in ancestry groups with more than 100 cases in UK Biobank. Associations were tested using logistic regression models, adjusted for genotyping array, sex and the first ten 10 principal components of ancestry. To further aid clinical interpretation, we divided individuals into deciles according to their PGSs and using logistic regression, investigated disease risk associated, comparing each decile to a reference decile. To evaluate the age-dependent PGS performance, we used the Kaplan–Meier method to generate a cumulative incidence plot and a log-rank test to test for differences between groups. In a sensitivity analysis, we assessed the possible impact of overfitting when testing the association between the PGS and binary disease phenotypes by excluding individuals used in the discovery TSH GWAS in UKB and closely related individuals (KING software relatedness coefficient >0.0884) from the testing datasets.

### Phenome-wide association study (PheWAS)

To identify pleiotropic associations with a wide range of phenotypes, we used DeepPheWAS(7), a flexible PheWAS framework which incorporates phenotypes not present in other PheWAS platforms for: (i) sentinel variants implicating genes supported by ≥3 variant-to-gene mapping criteria (**Supplementary Table 2**), variants in a credible set that were annotated as missense/damaging/deleterious/phred-scaled CADD score ≥20 that also had a posterior probability >50% (**Supplementary Table 21**), and low-frequency sentinel variants (MAF<1%, **Supplementary Table 1**); (ii) the PGS for TSH; (iii) pathway-specific genetic risk score (GRS).

### Druggability

To identify gene products that are the targets of drugs, we queried the Drug Gene Interaction Database (DGIDB) (https://www.dgidb.org) for the 112 putative causal genes supported by ≥2 variant-to-gene criteria. Genes were mapped to ChEMBL interactions and indications (MeSH headings).

## Data Availability

Genome-wide summary statistics will be deposited at GWAS Catalog (EMBL-EBI) upon acceptance of the manuscript.

## Acknowledgements

The research was partially supported by the NIHR Leicester Biomedical Research Centre and through an NIHR Senior Investigator Award to **M.D. Tobin**; views expressed are those of the author(s) and not necessarily those of the NHS, the NIHR or the Department of Health. The funders had no role in the design of the study.

This research was funded in whole, or in part, by the Wellcome Trust: Wellcome Trust Investigator Award (WT202849/Z/16/Z, MD Tobin) and Wellcome Trust Discovery Award (WT225221/Z/22/Z, MD Tobin and L.V.Wain). For the purpose of open access, the author has applied a CC BY public copyright licence to any Author Accepted Manuscript version arising from this submission.

**L. V. Wain** was supported by GSK/British Lung Foundation Chair in Respiratory Research.

**C. Batini** was supported by a UKRI Innovation Fellowship at Health Data Research UK (MR/S003762/1).

**C. John** held a Medical Research Council Clinical Research Training Fellowship (MR/P00167X/1).

EXCEED is supported by the University of Leicester, the NIHR Leicester Respiratory Biomedical Research Centre, by Wellcome [202849, https://doi.org/10.35802/202849] and by Cohort Access fees from studies funded by the Medical Research Council (MRC), BBRSC, NIHR, the UK Space Agency, and GSK. It was previously supported by MRC grant G0902313. This work is supported by BREATHE - The Health Data Research Hub for Respiratory Health [UKR_PC_19004] in partnership with SAIL Databank. We also thank all participants and staff who have contributed their time to the study.

Our analyses of UK Biobank and EXCEED used the ALICE and SPECTRE High Performance Computing Facilities at the University of Leicester.

We want to acknowledge the participants of the Estonian Biobank for their contributions. The Estonian Biobank analyses were partially carried in the High Performance Computing Center, University of Tartu.

We would like to thank the ThyroidOmics consortium, Nord-Trøndelag Health (HUNT) Study, and Michigan Genomics Initiative for depositing genome-wide summary statistics from previous studies in GWAS Catalog, and we are grateful to the staff who develop and maintain the GWAS Catalog.

## Ethical approval

The UK Biobank genetic and phenotypic data were analysed under UK Biobank Application 43027. UK Biobank has ethical approval from the UK National Health Service (NHS) National Research Ethics Service (11/NW/0382). EXCEED received ethical approval from the Leicester Central Research Ethics Committee (13/EM/0226).

The activities of the EstBB are regulated by the Human Genes Research Act, which was adopted in 2000 specifically for the operations of EstBB. Individual level data analysis in EstBB was carried out under ethical approval 1.1-12/624 from the Estonian Committee on Bioethics and Human Research (Estonian Ministry of Social Affairs), using data according to release application 6-7/GI/2013 from the Estonian Biobank.

Informed consent was obtained from all participants.

## Data availability

Access to UK Biobank and EXCEED datasets is available to bona fide researchers upon application (in accordance with the terms of ethical approval and participant consent). Genome-wide summary statistics will be made publicly available via the EMBL-EBI GWAS Catalog.

## Author Contributions

C.J. and M.D.T. supervised the study. A.T.W., J.C., N.S., M.D.T. and C.J. designed the study. A.T.W., J.C., K.C., C.B., A.I., R.P., E.A., and N.S. performed statistical analyses. A.T.W., J.C., K.C., C.B., A.I., R.P., E.A., E.J.H., W.H., B.S.R., F.D., L.V.W., N.S., M.D.T., C.J. analysed and/or interpreted the data. F.D. and L.V.W. provided methodological and statistical advice. A.T.W., J.C., N.S., M.D.T. and C.J. wrote the manuscript. All remaining co-authors contributed to data collection, management or analysis of the EXCEED study or the Estonian Biobank. All co-authors critically reviewed the manuscript.

