## Supplementary Material for "Genome-wide association study of thyroid-stimulating hormone highlights new genes, pathways and associations with thyroid disease susceptibility and age-of-onset"

Supplementary Tables and Figures for the manuscript entitled "Genome-wide association study of thyroid-stimulating hormone highlights new genes, pathways and association with thyroid disease susceptibility and age-of-onset".

### Supplementary Note

**Estonian Biobank (EstBB)**

All EstBB participants have been genotyped at the Core Genotyping Lab of the Institute of Genomics, University of Tartu, using Illumina Global Screening Array v3.0_EST. Samples were genotyped and PLINK format files were created using Illumina GenomeStudio v2.0.4. Individuals were excluded from the analysis if their call-rate was <95% or if sex defined based on heterozygosity of X chromosome did not match sex in the phenotype data. Before imputation, variants were filtered by call-rate <95%, Hardy-Weinberg *P*<1×10^-4^ (autosomal variants only), and minor allele frequency <1%. Genotyped variant positions were in build 37 and were lifted over to build 38 using Picard. Phasing was performed using the Beagle v5.4 software. Imputation was performed with Beagle v5.4 software (beagle.22Jul22.46e.jar) and default settings. The dataset was split into batches of 5,000. A population-specific reference panel consisting of 2,695 whole-genome sequencing (WGS) samples was utilised for imputation and standard Beagle hg38 recombination maps were used. Based on principal component analysis, samples that were not of European ancestry were removed. Duplicates and monozygous twin were identified using KING 2.2.7, and one sample was removed out of every pair of duplicates.

Association analyses in Estonian Biobank were carried out for all variants with an imputation quality score >0.4 under an additive model as implemented in REGENIE v3.0.3 with standard quantitative trait settings. Linear regression was carried out on inverse normalised TSH concentration values and adjusted for current age, age², sex and 10 principal components as covariates, analysing only variants with a minimum minor allele count of 2.

**Extended Cohort for E-Health, Environment and DNA (EXCEED)**

Samples were genotyped using the UK Biobank Axiom array. Prior to imputation, individuals were excluded if their call rate was <97% or if genetic sex and phenotypic sex did not match; variants were excluded if the call rate was <95%, Hardy-Weinberg *P*<1×10^-6^ or minor allele frequency <1%. Imputation was conducted using the TOPMed Imputation Server (phasing: Eagle v2.4, imputation: Minimac4, reference panel: TOPMed r2).

Association analyses in EXCEED were conducted for all variants with an imputation quality score >0.5 and a minor allele frequency >0.1% under an additive genetic model adjusted for age, sex and the first 10 principal components of ancestry using PLINK 2.0.

**UK Biobank (UKB)**

Genotyping and imputation for UK Biobank samples are described in detail in the following source: Bycroft C, Freeman C, Petkova D, et al. The UK Biobank resource with deep phenotyping and genomic data. Nature. 2018 Oct;562(7726):203-209. doi: 10.1038/s41586-018-0579-z

Association analyses in UK Biobank were conducted for all variants with an imputation quality score >0.5 and a minor allele count >20 under an additive genetic model adjusted for age, genotyping array, sex and the first 10 principal components of ancestry using PLINK 2.0.

### Supplementary Figures

**Supplementary Figure 1**: Single variant PheWAS

a) **rs115315671 (implicating *ADCY6*).**
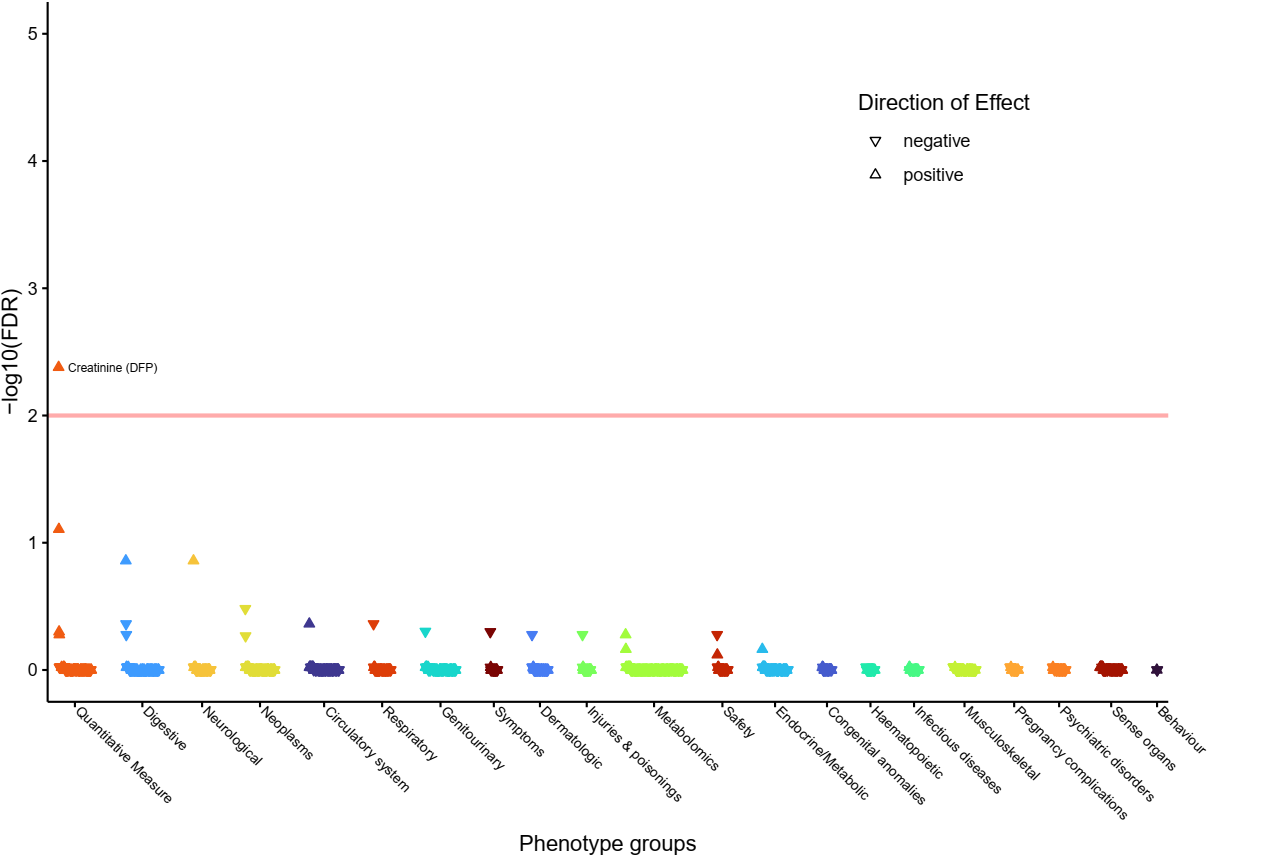

b) **rs2494734 (implicating *AKT1* and *ZBTB42*).**
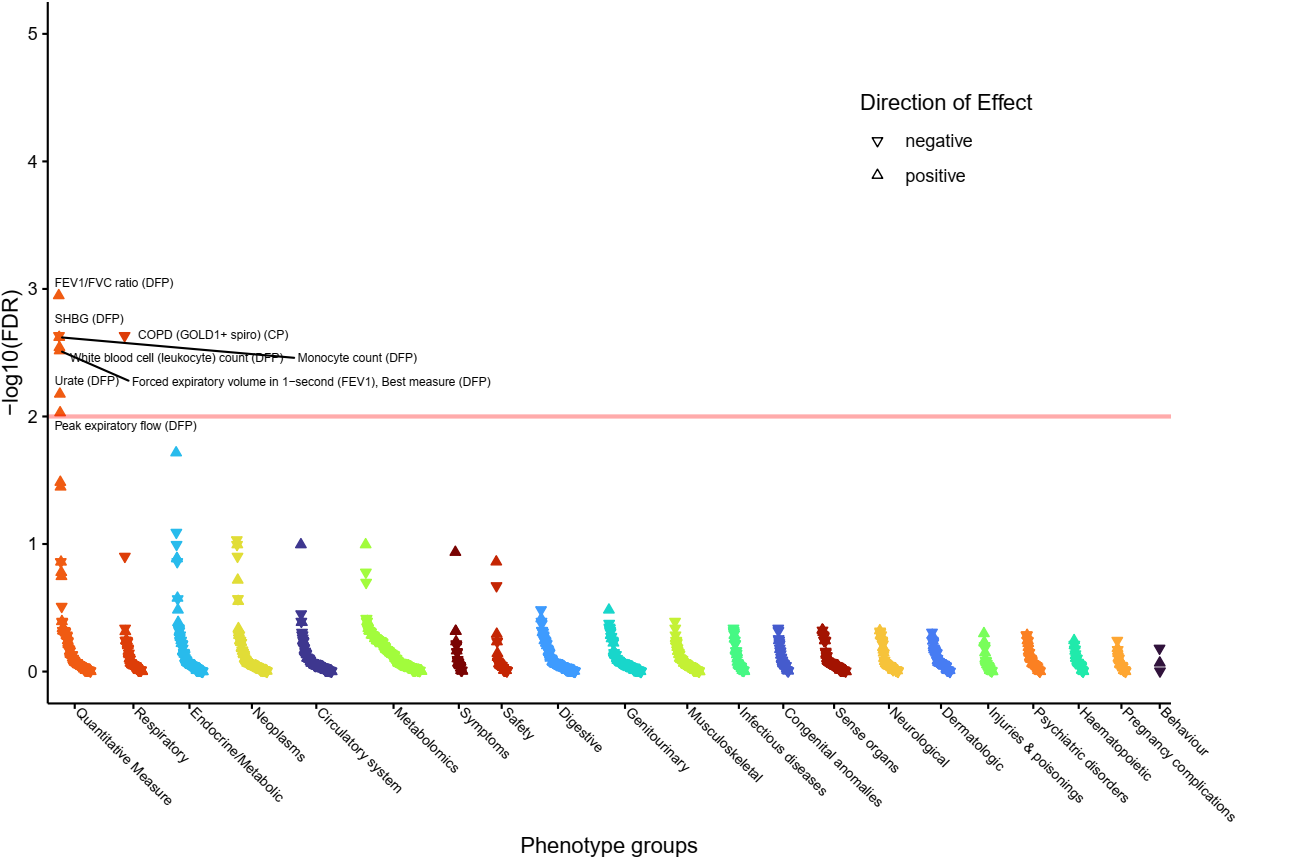

c) **rs17450274 (implicating *ANXA5*).**
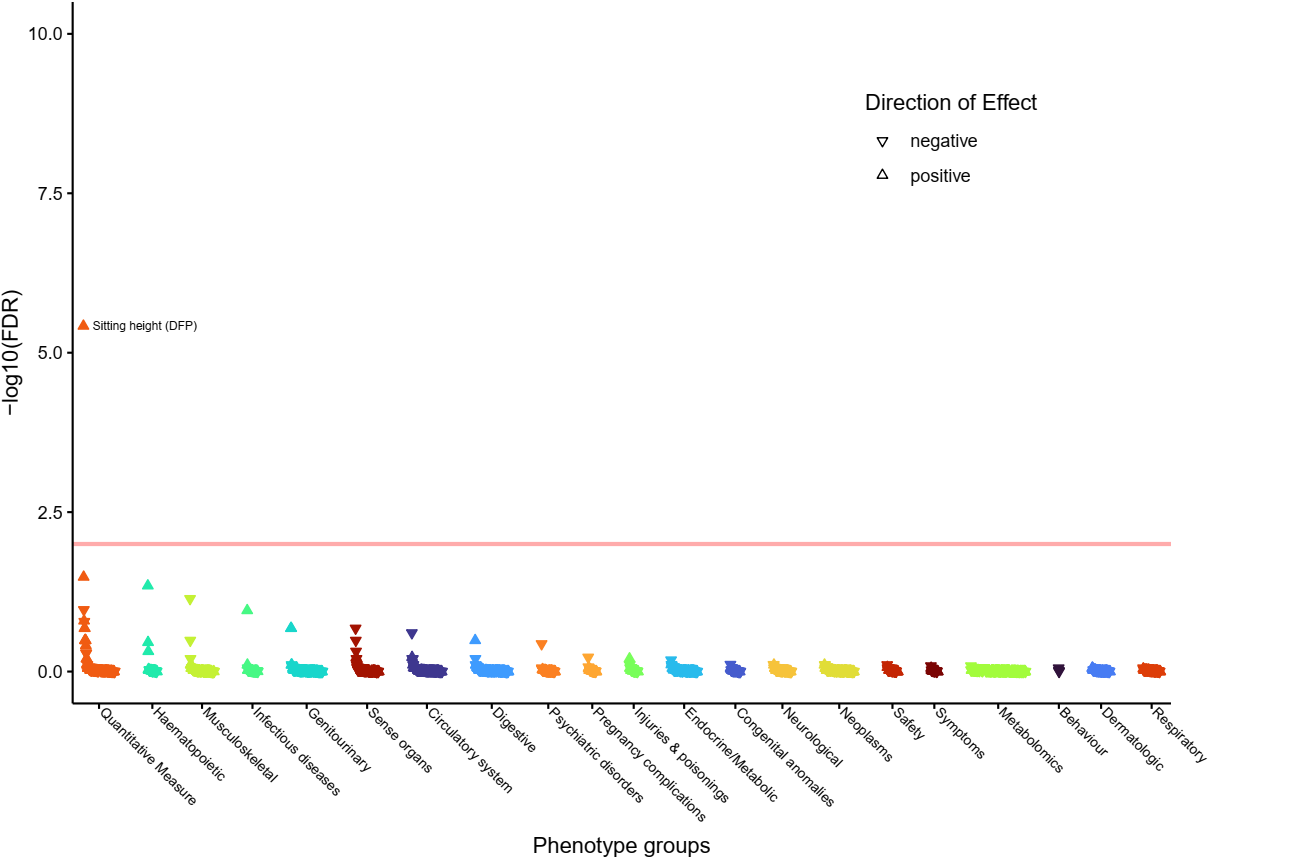

d) **rs11605461 (implicating *CADM1*).**
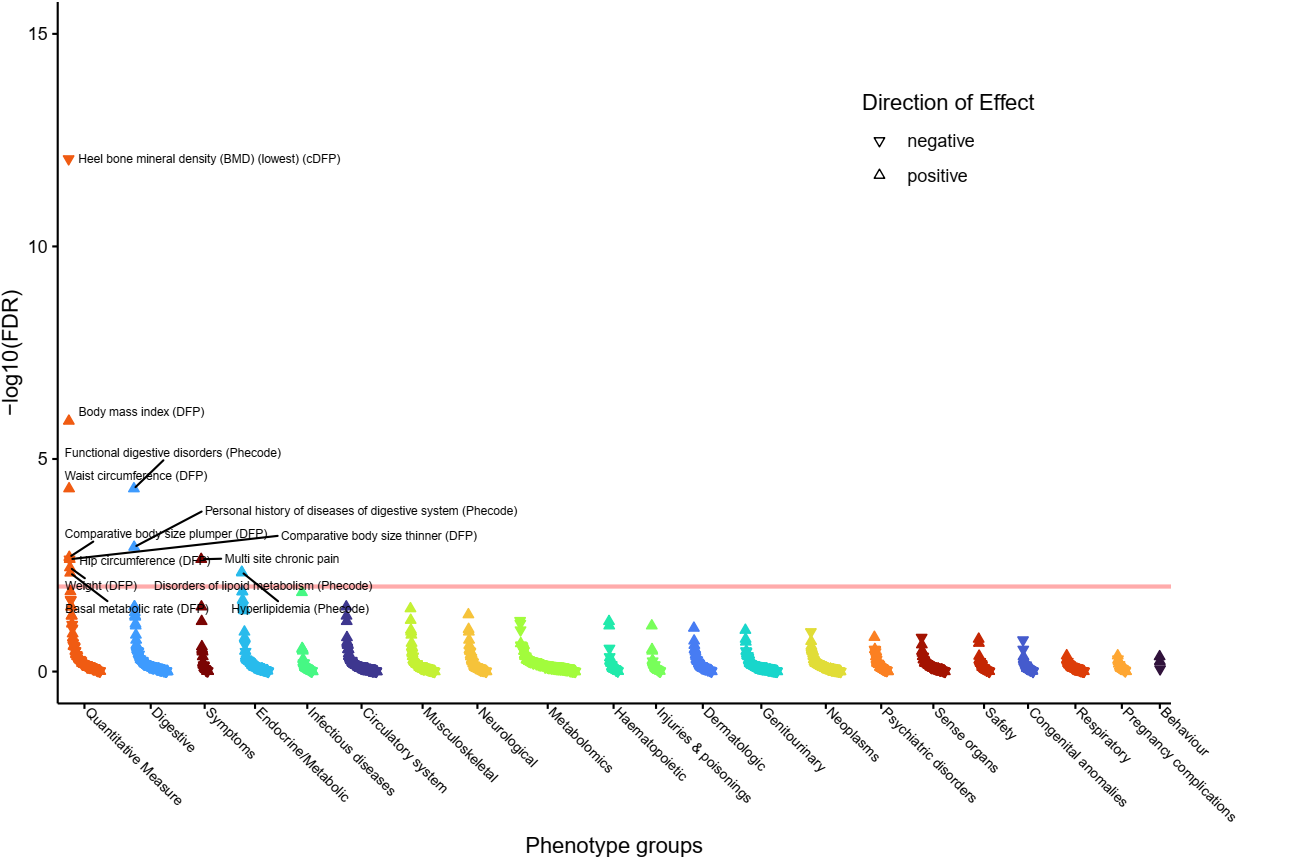

e) **rs149363012 (implicating *CCDC77*, *WNK1* and *B4GALNT3*).**
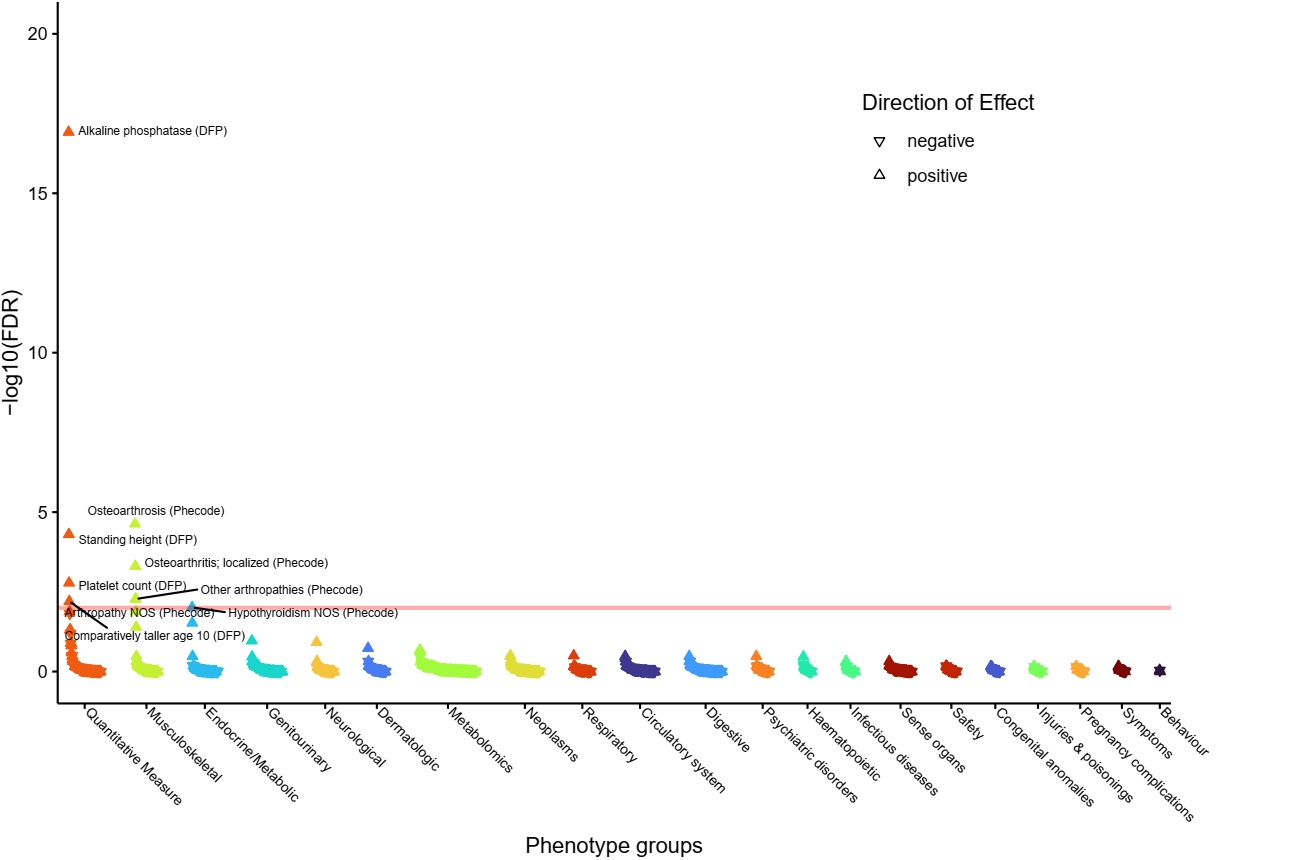

f) **rs3124747 (implicating *DBH*, *C9orf96* and *STKLD1*).**
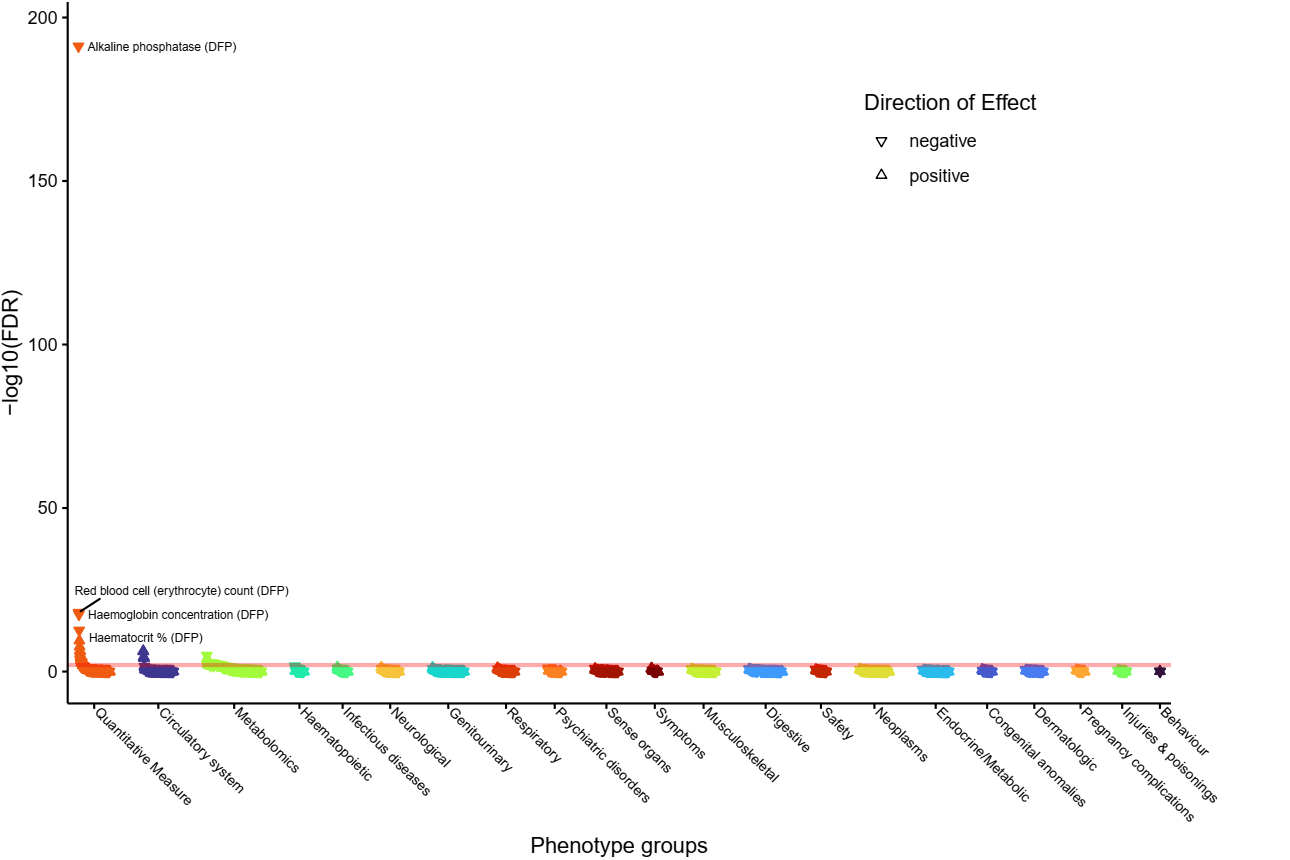

g) **rs139411099 (implicating *DIO2*).**
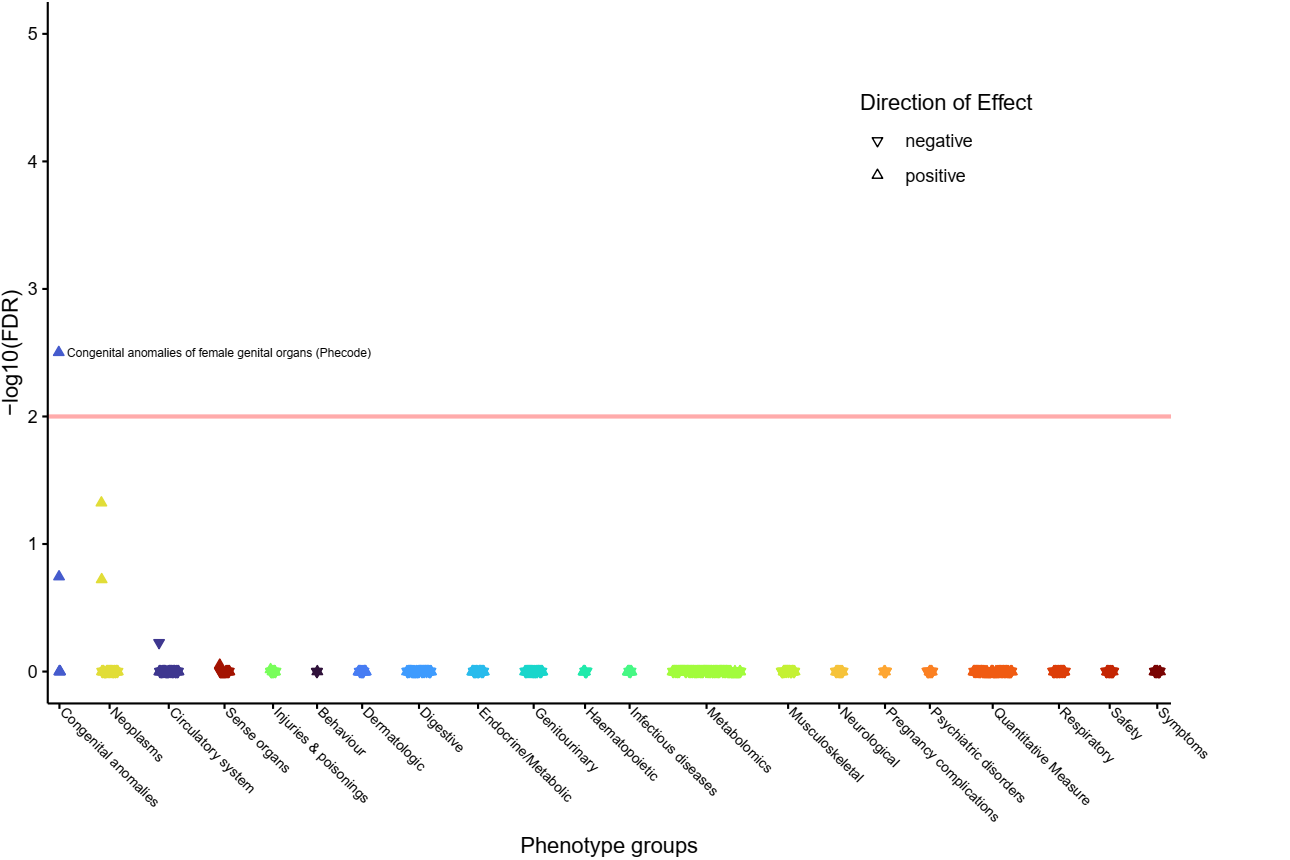

h) **rs2282192 (implicating *FOXE1*, *ANP32B* and *C9orf156*).**
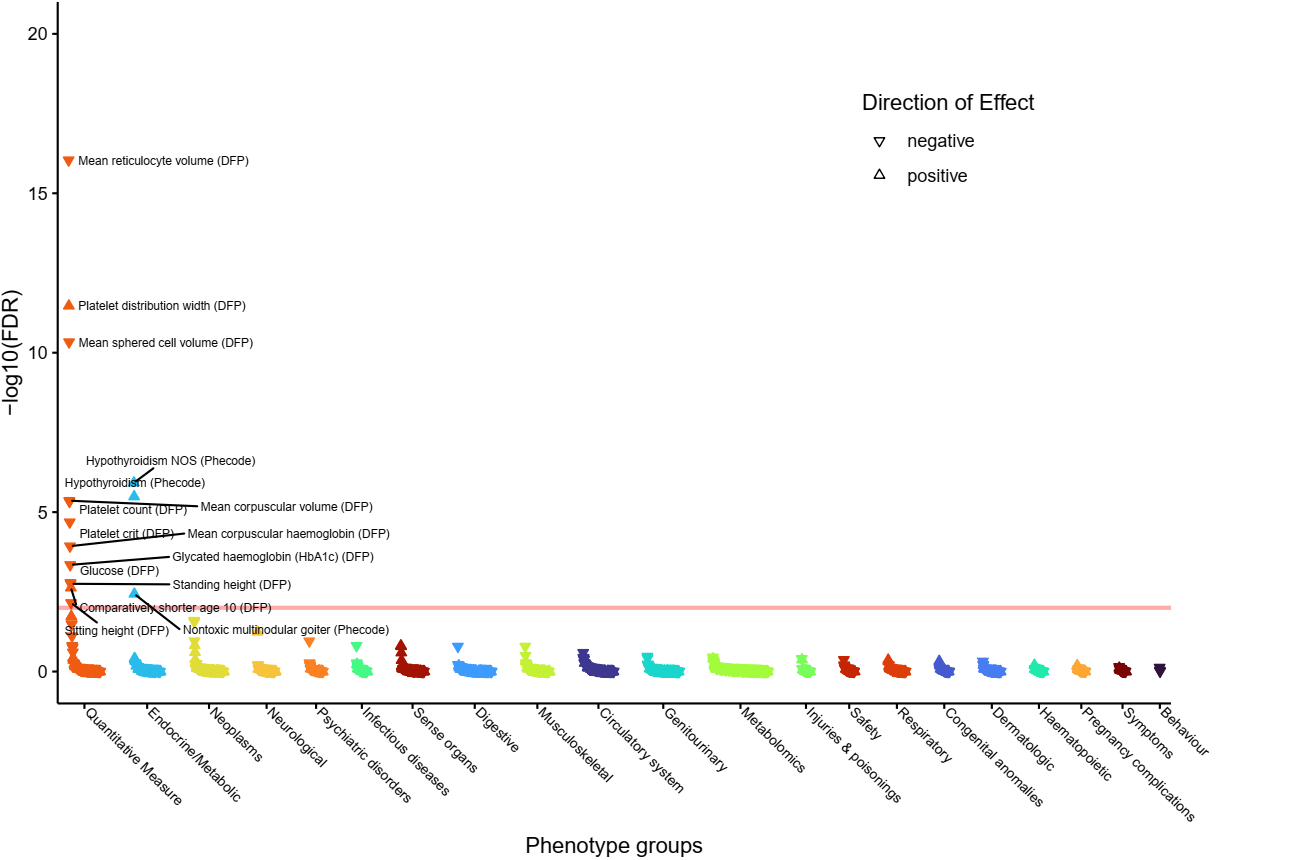

i) **rs10814915 (implicating *GLIS3*).**
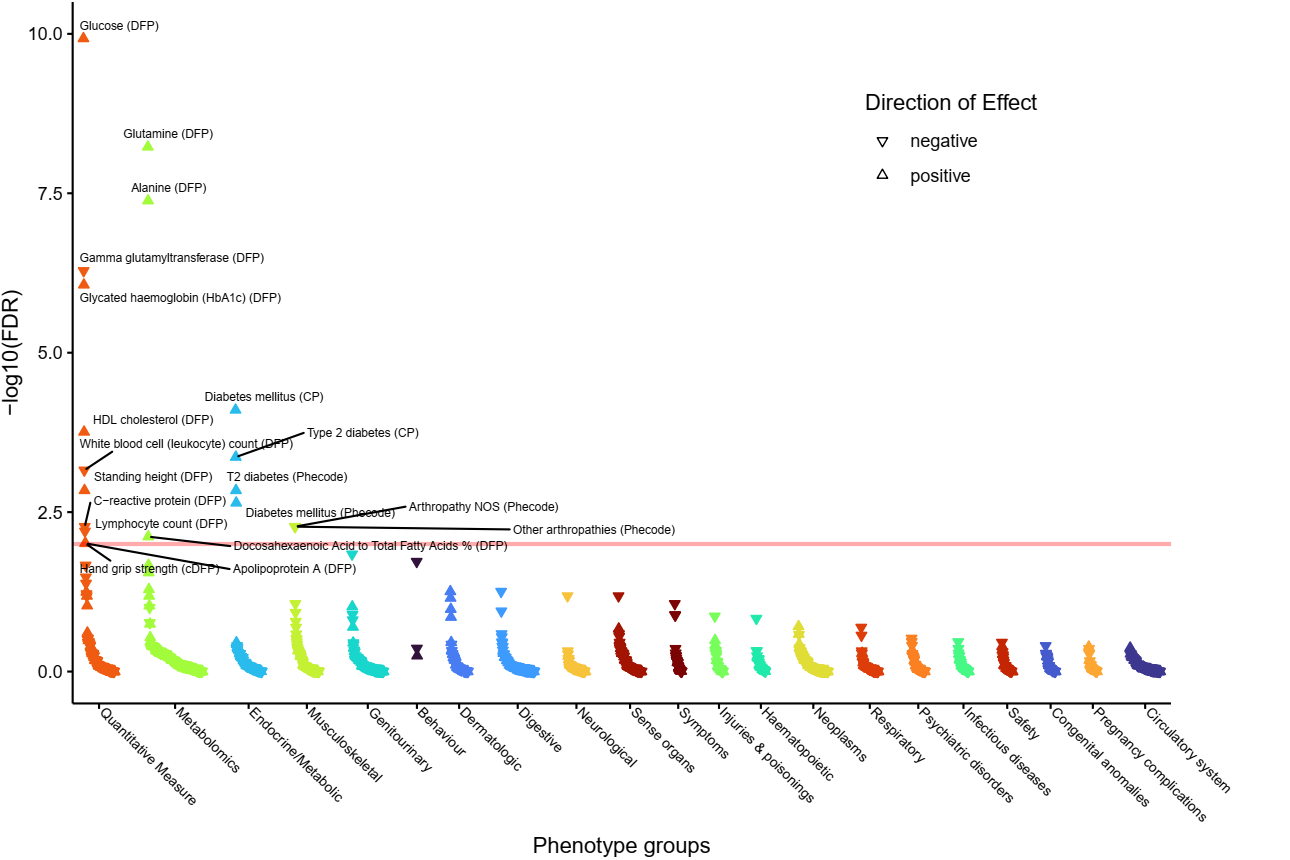

j) **rs1861628 (implicating *IGFBP5*).**
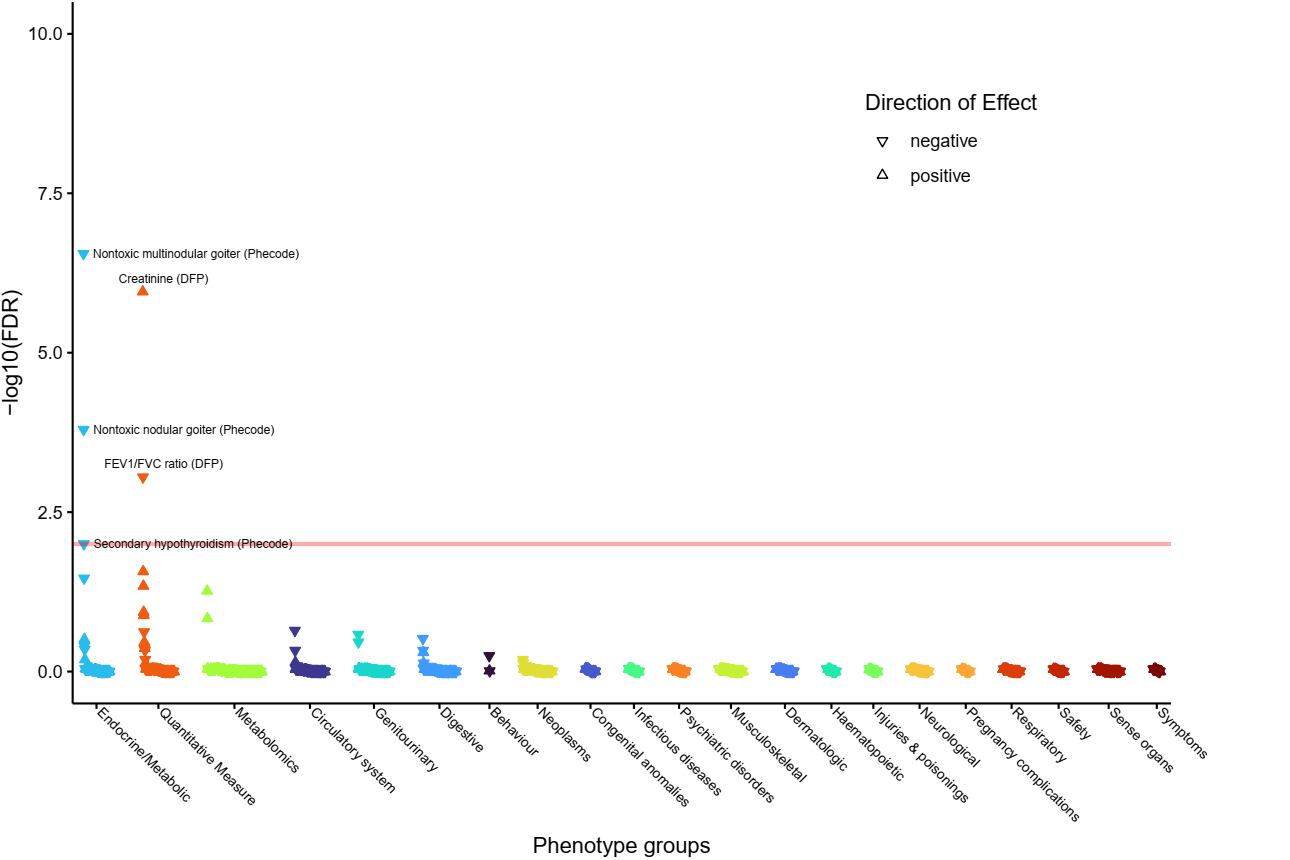

k) **rs1861630 (implicating *IGFBP5*).**
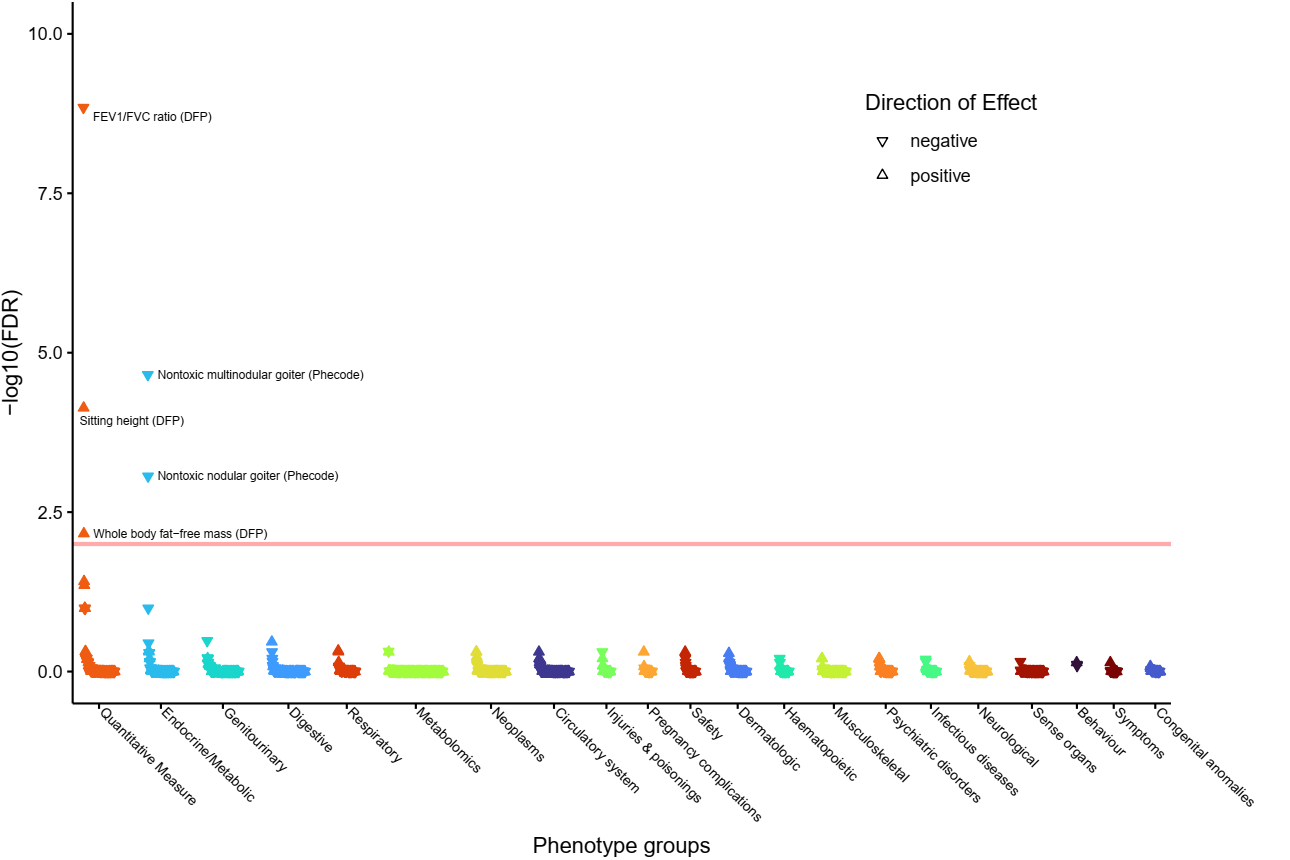

l) **rs6711608 (implicating *IGFBP5*).**
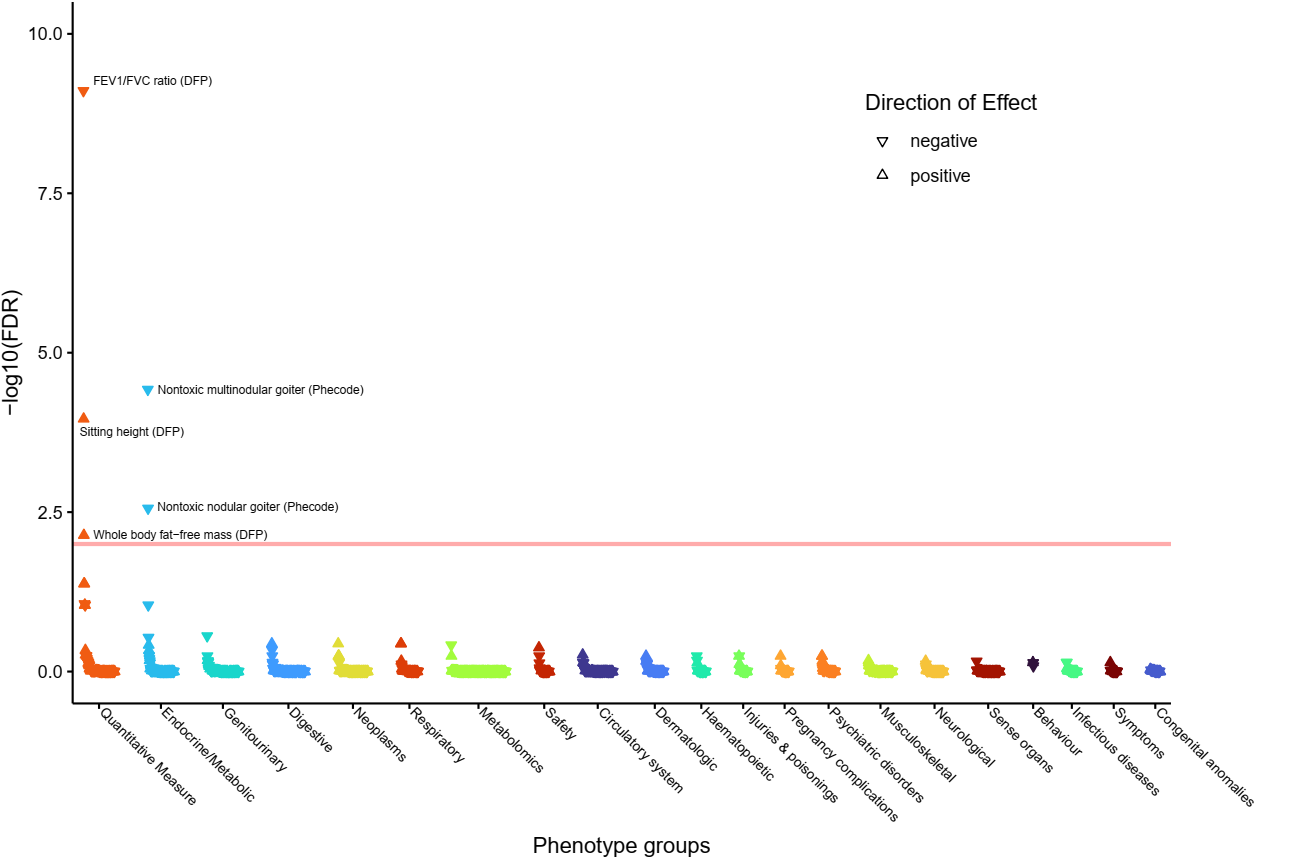

m) **rs57866041 (implicating *IGFBP5*).**
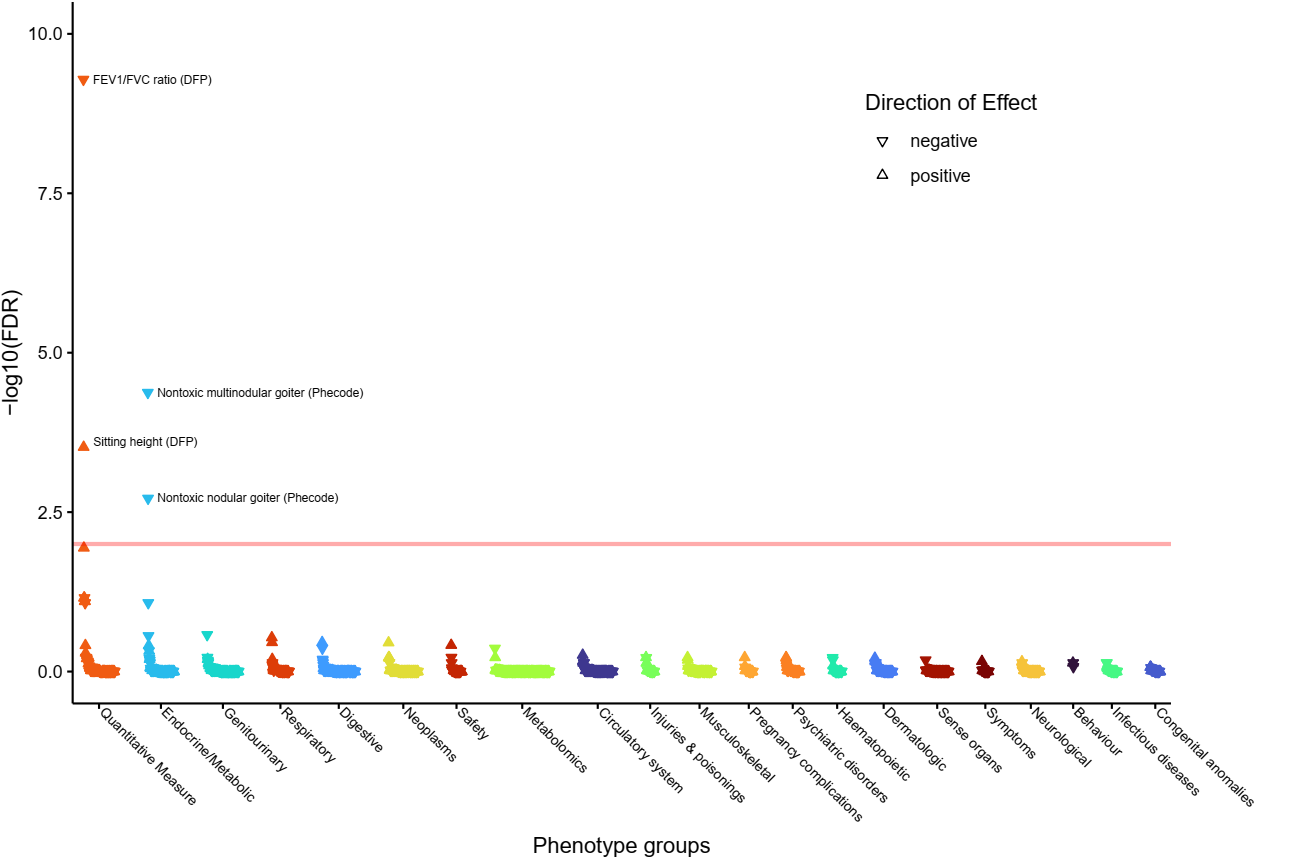

n) **rs2229738 (implicating *IGHMBP2*, *CPT1A* and *SCD*).**
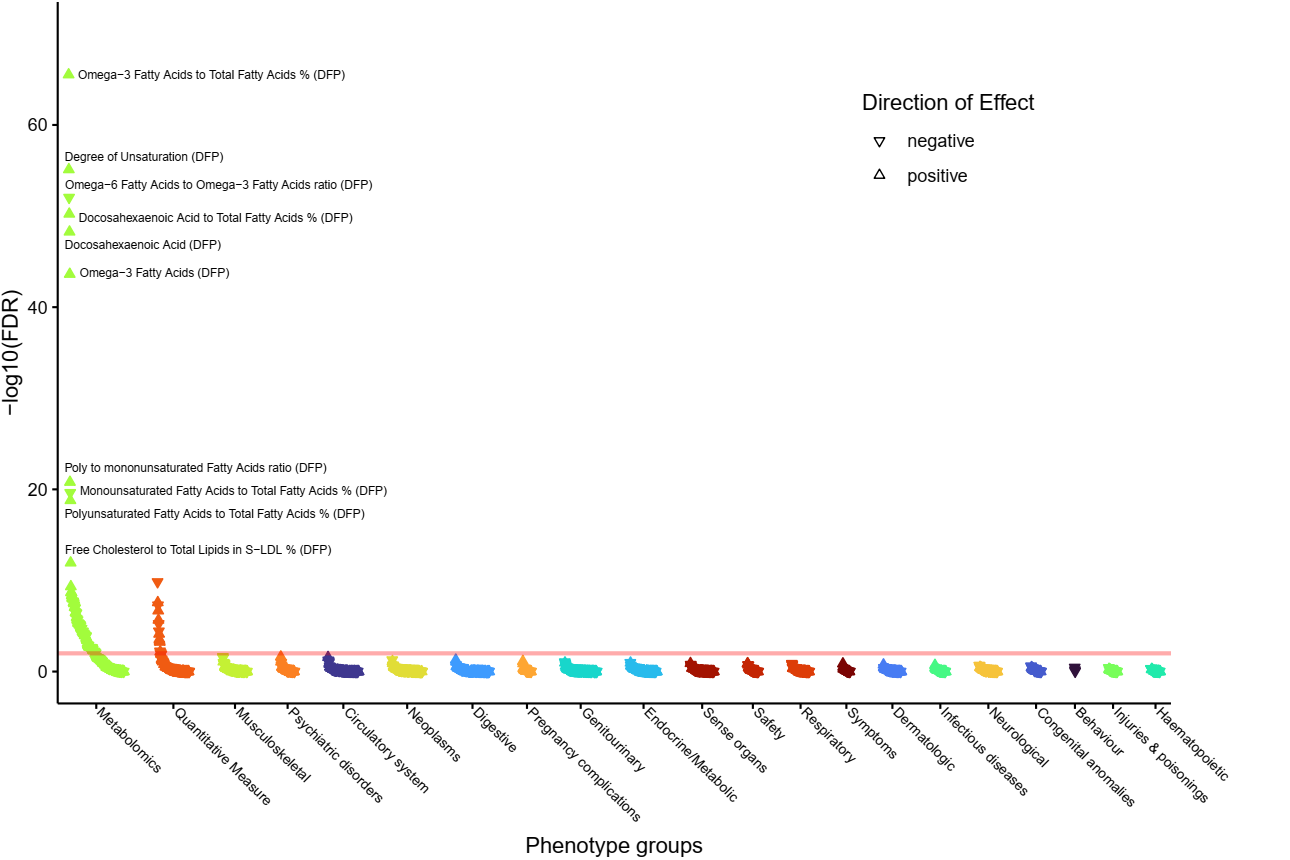

o) **rs4804416 (implicating *INSR* and *ACTL9*).**
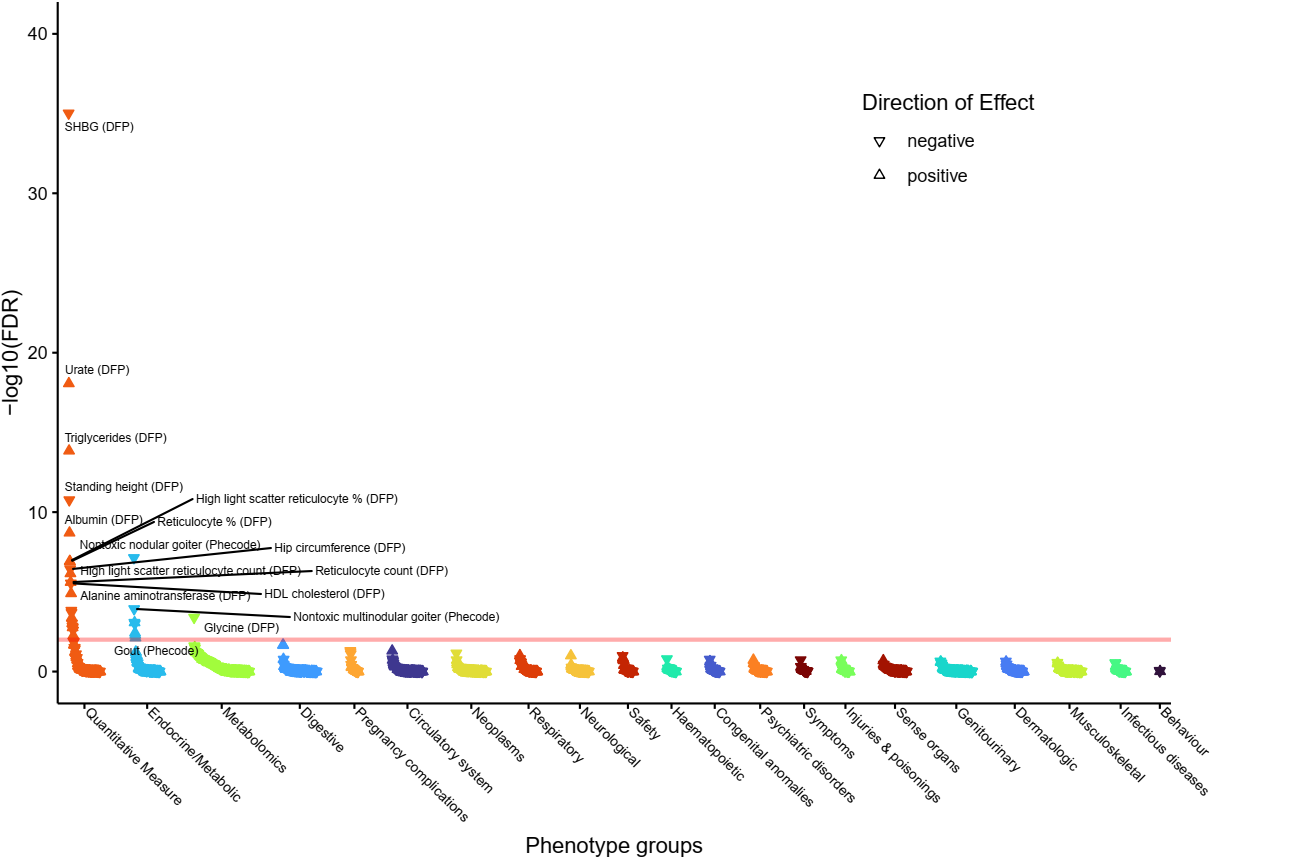

p) **rs7306508 (implicating *KRT18*).**
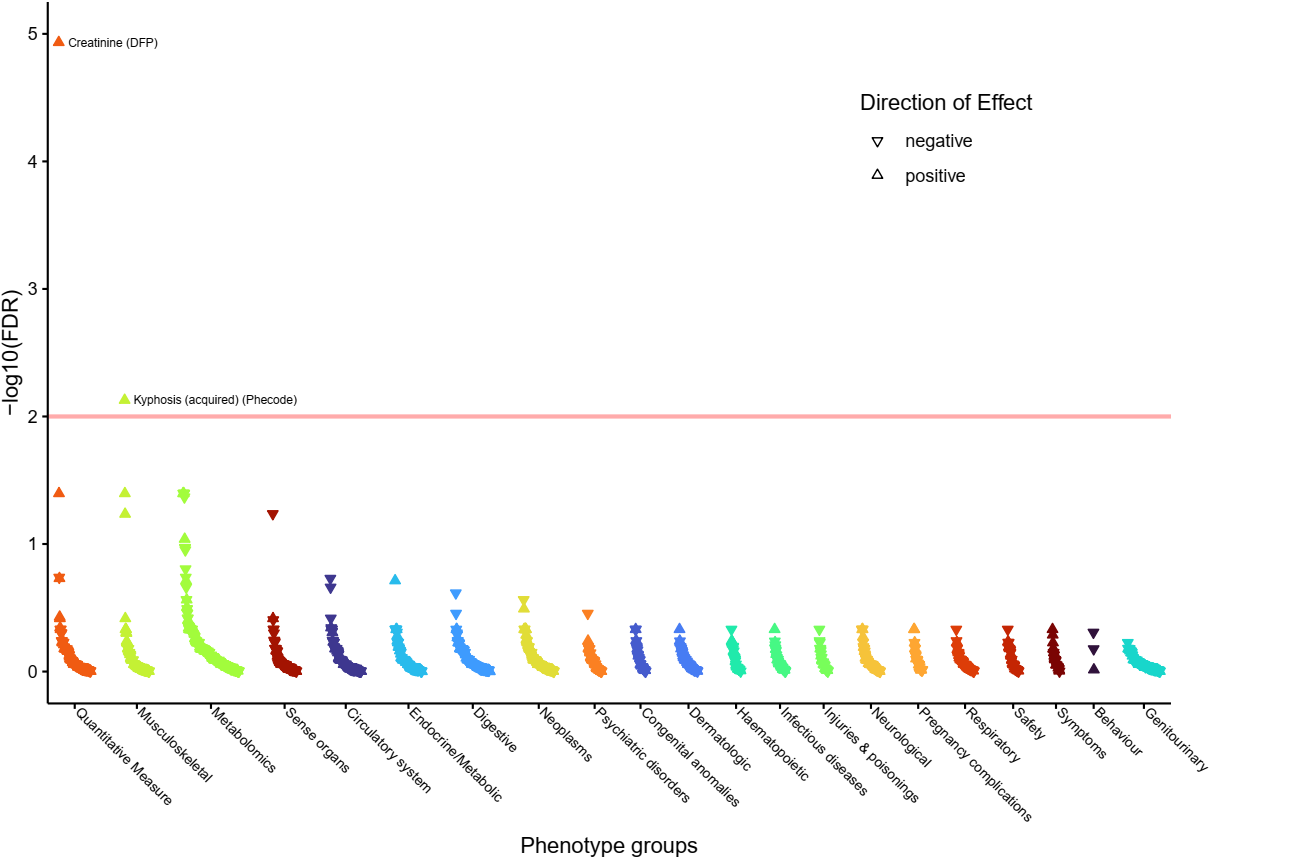

q) **rs2393717 (implicating *MLEC* and *SPPL3*).**
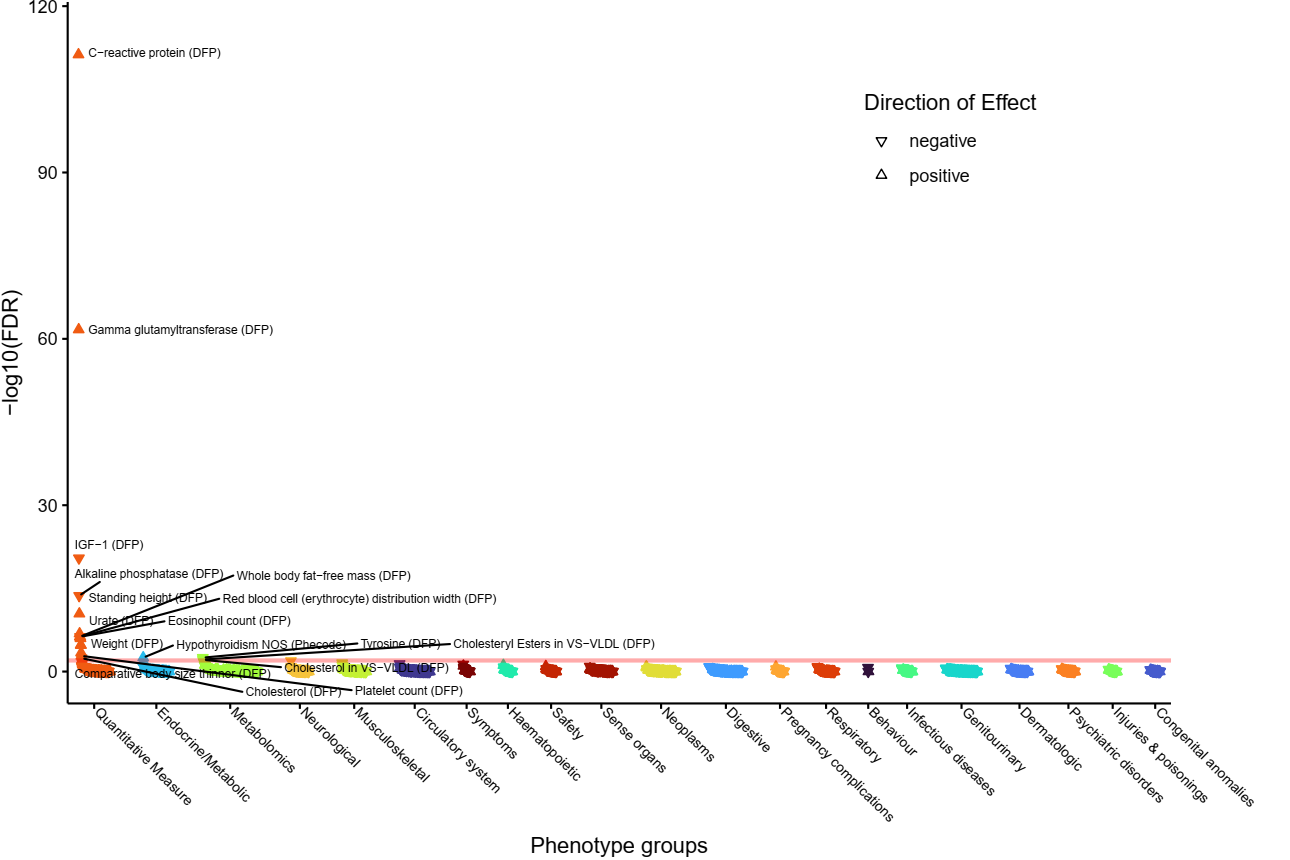

r) **rs36213229 (implicating *NRG1*).**
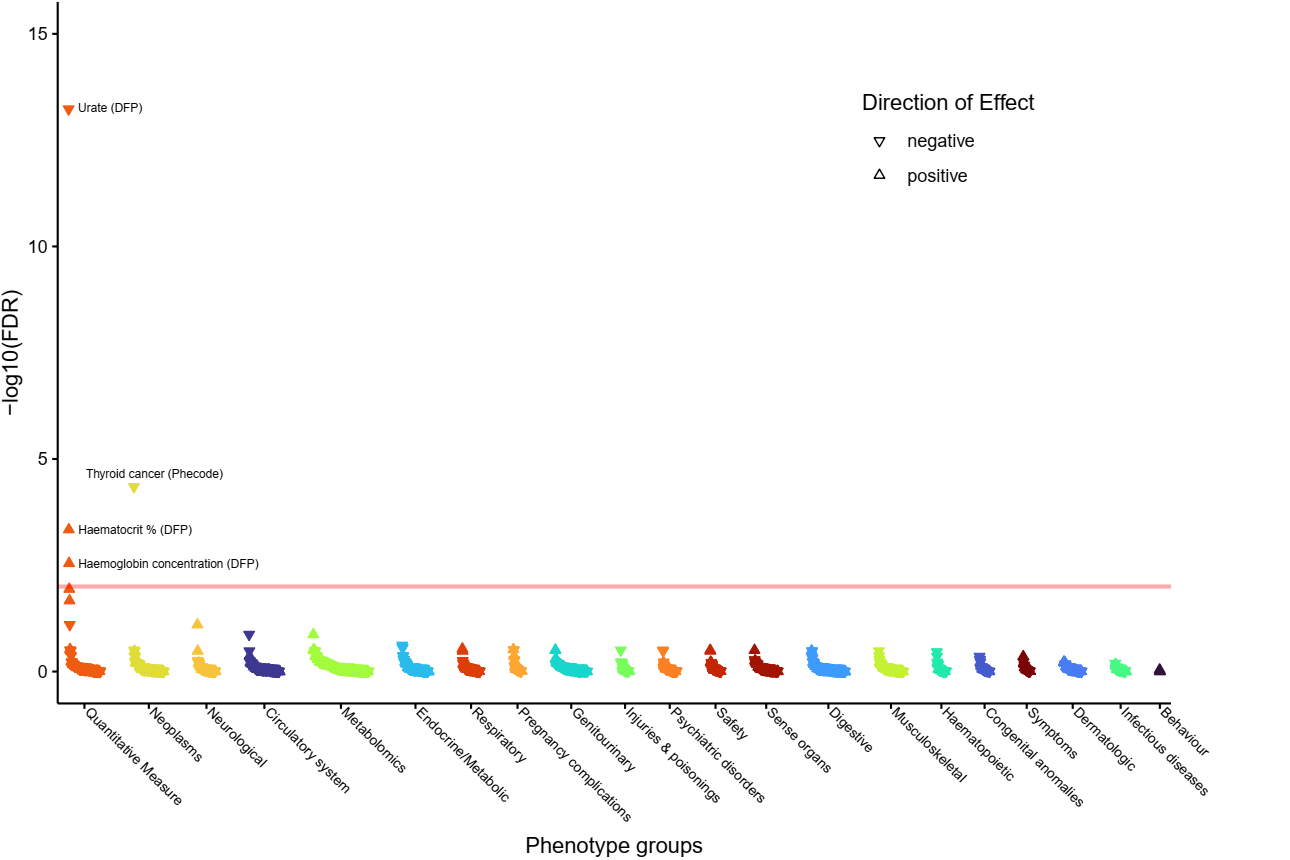

s) **rs68030583 (implicating *NRG1*).**
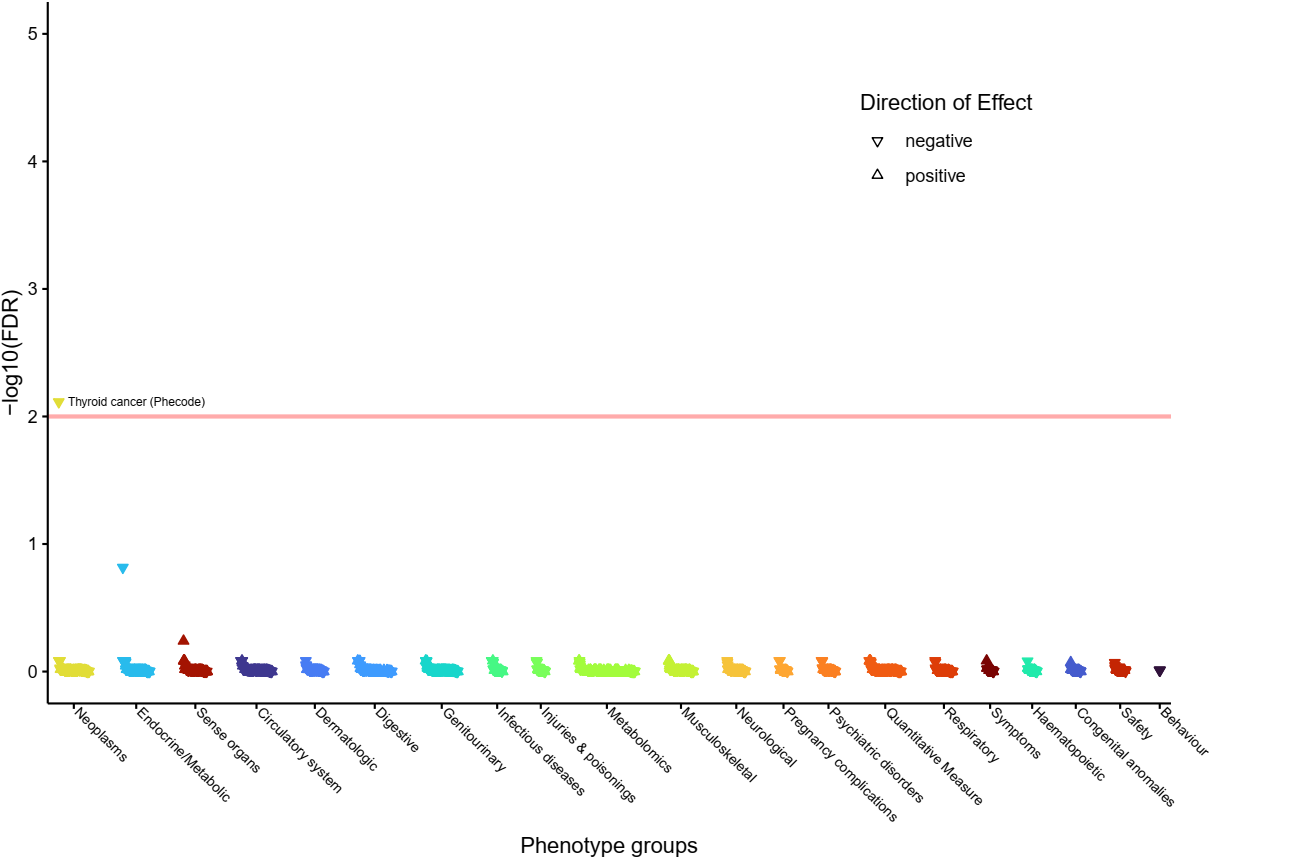

t) **rs11554674 (implicating *PHC2*, *ZNF362* and *A3GALT2*).**
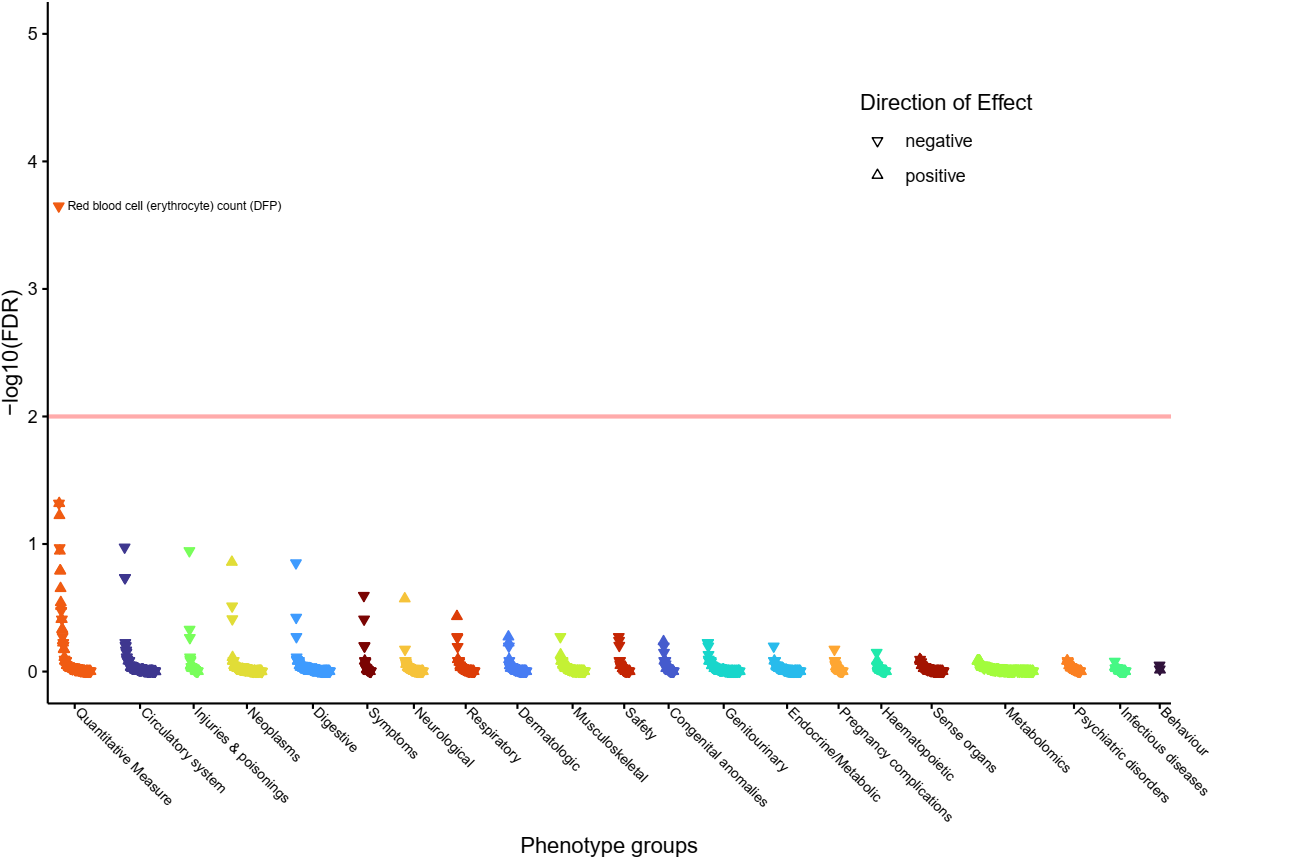

u) **rs1801690 (implicating *PRKCA* and *APOH*).**
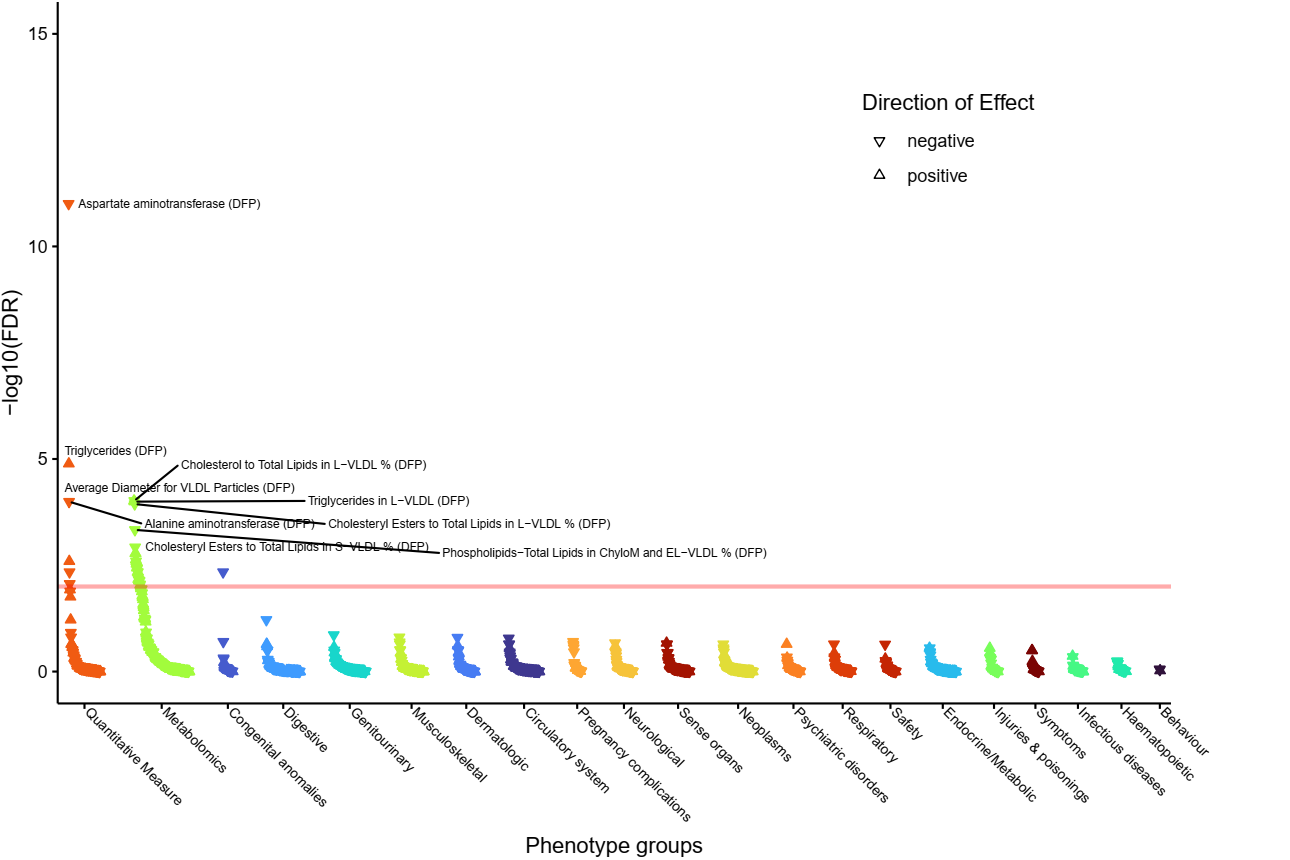

v) **rs9497965 (implicating *SASH1*).**
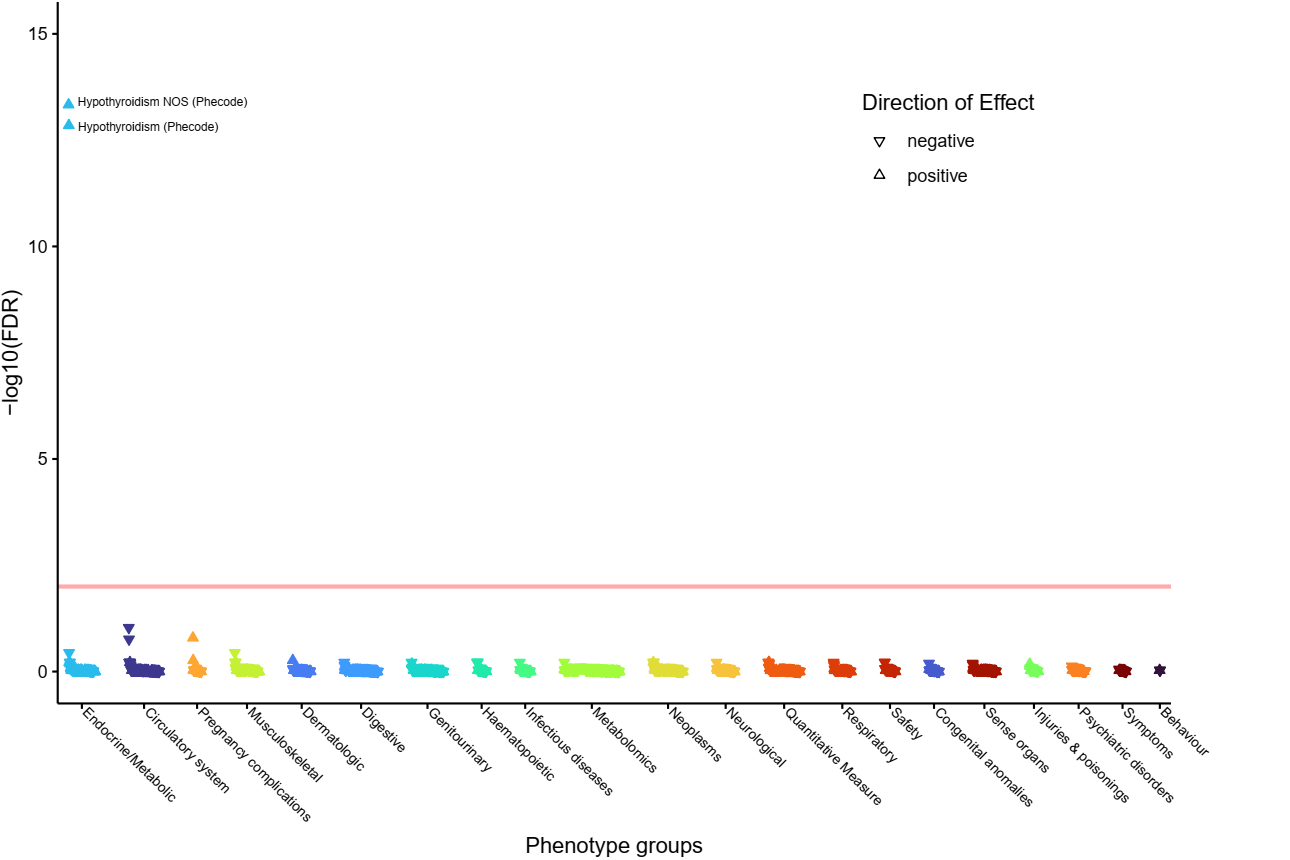

w) **rs10926981 (implicating *SDCCAG8*).**
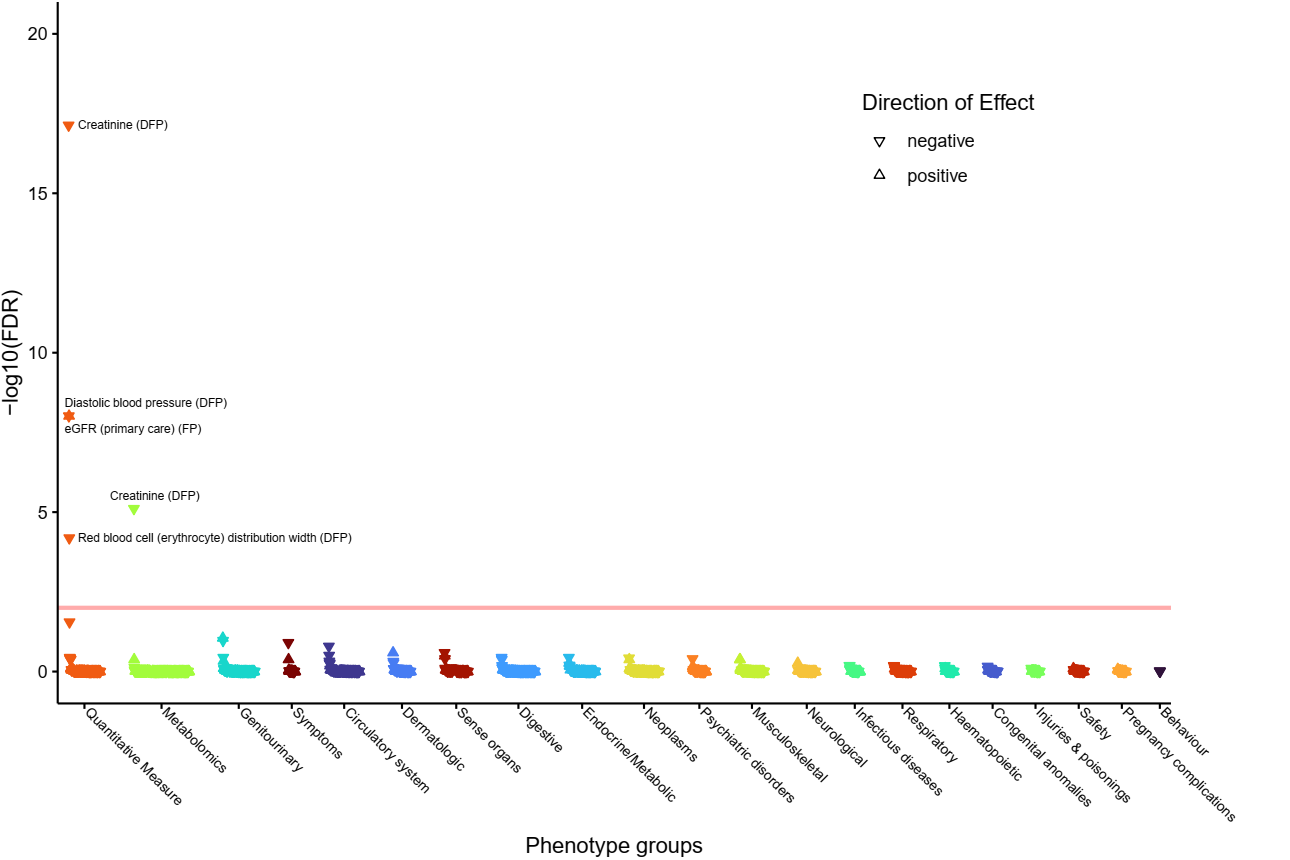

x) **rs1743963 (implicating *SGK1*).**
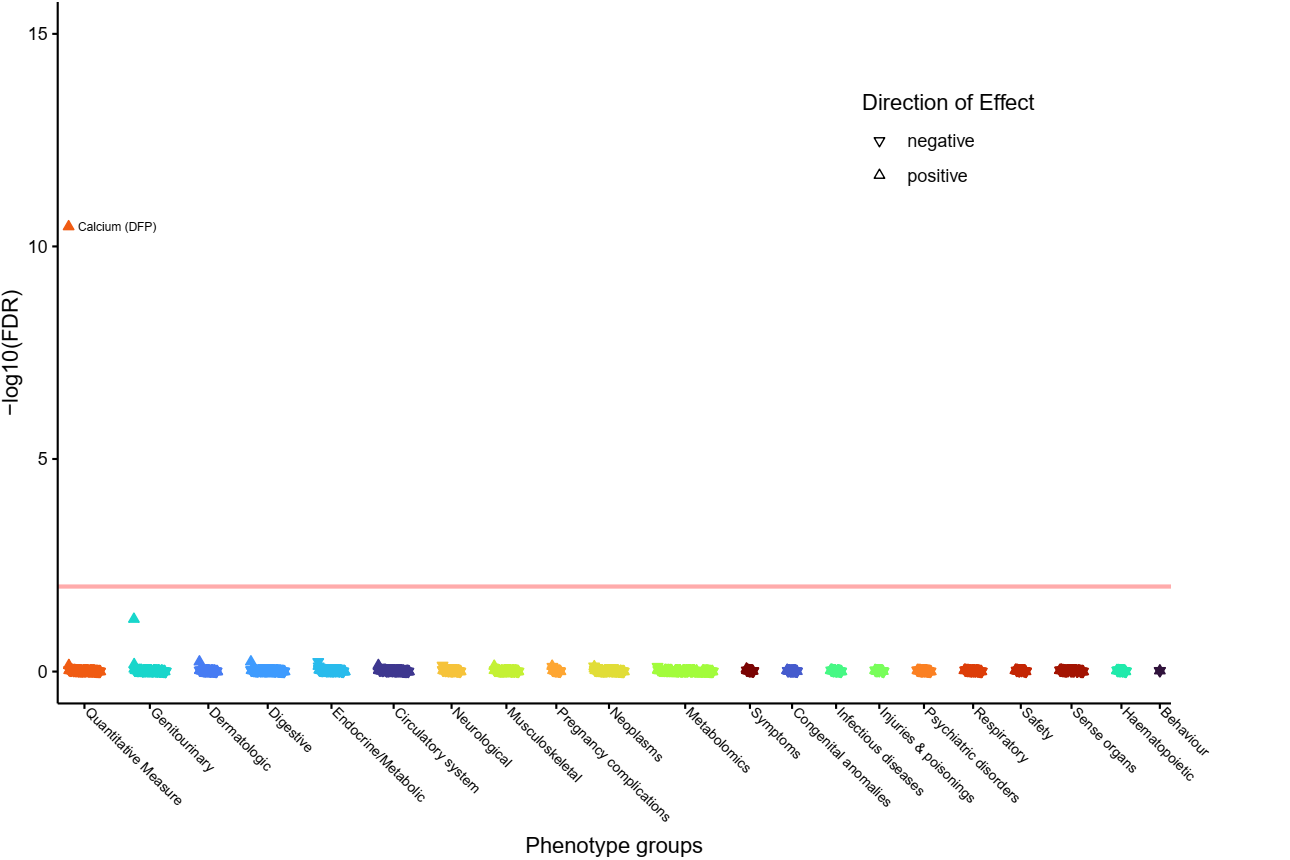

y) **rs3184504 (implicating *SH2B3*).**
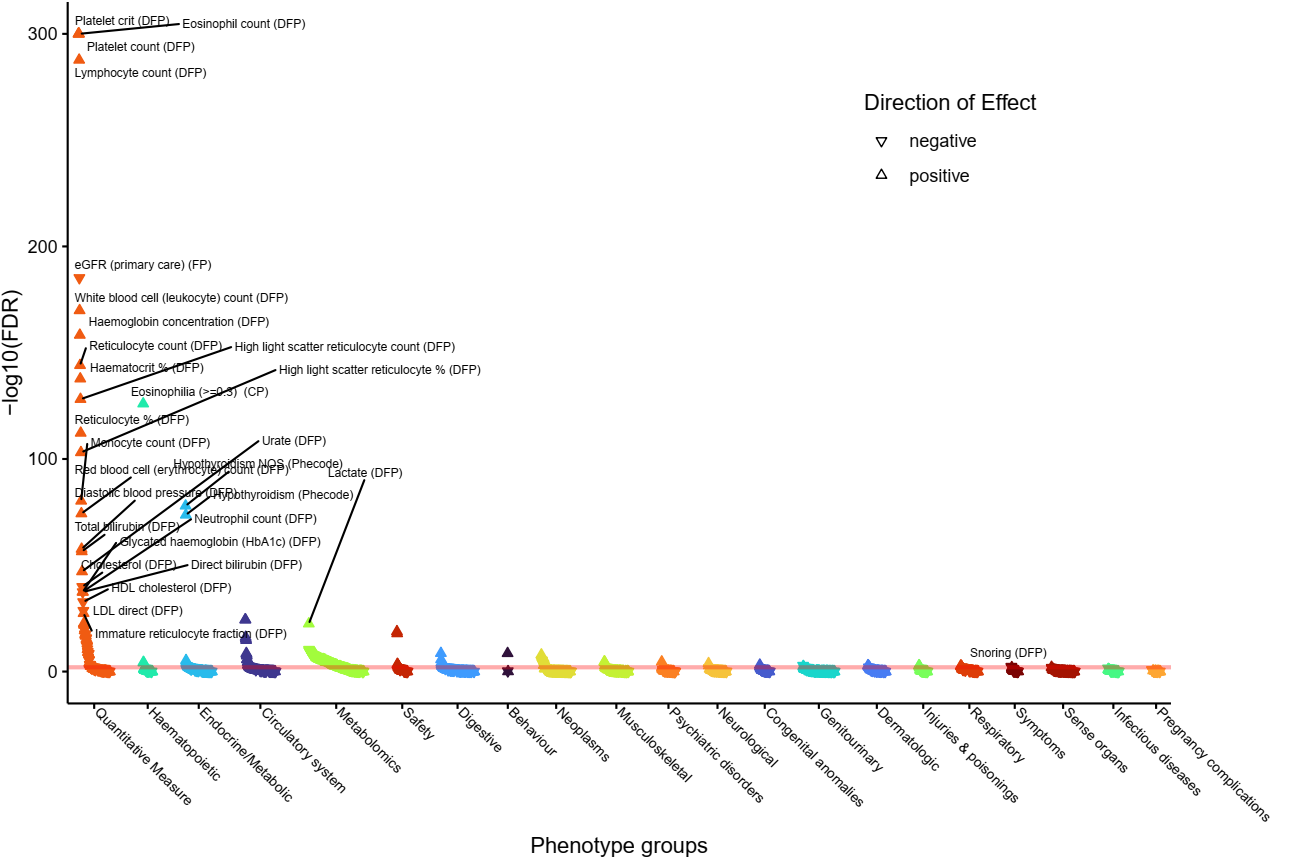

z) **rs751171 (implicating *SMOC2*).**
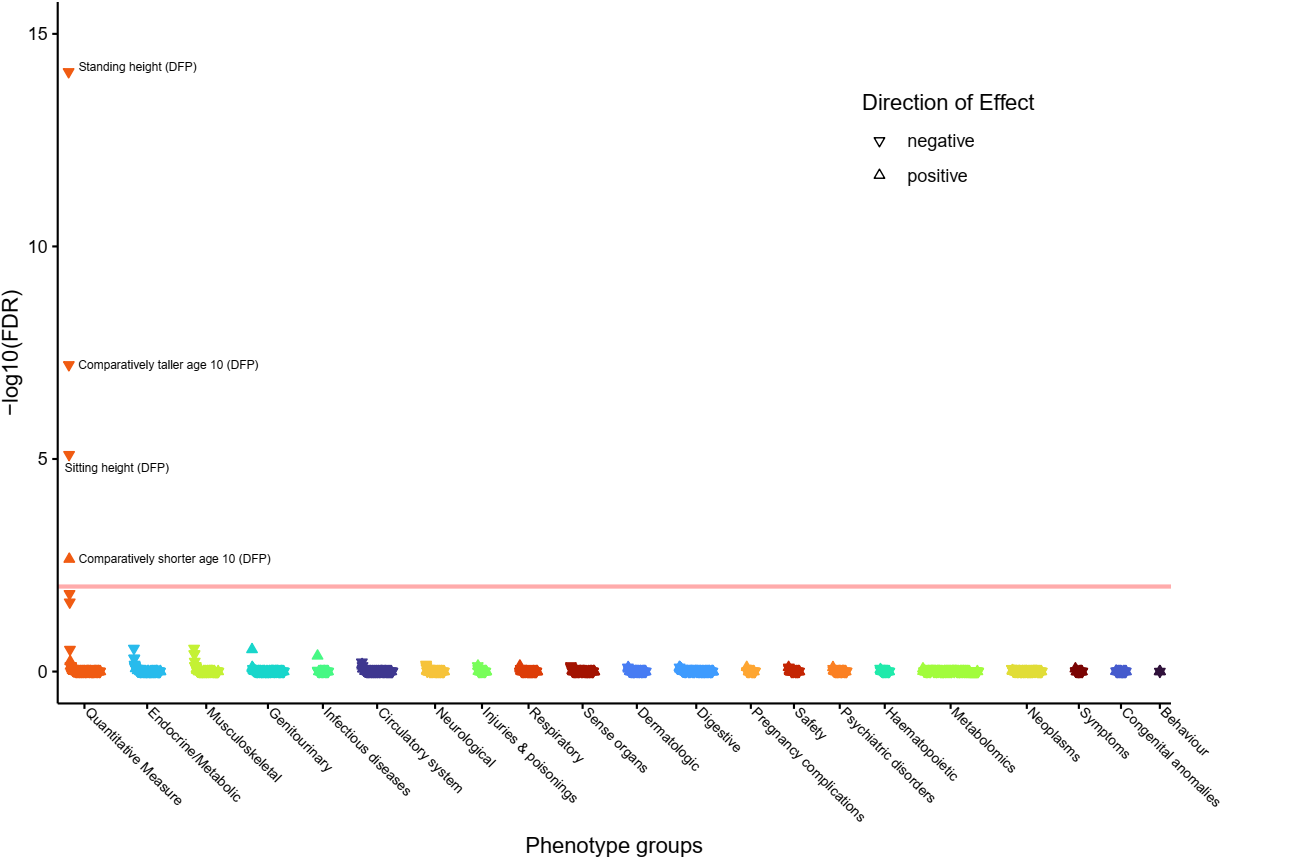

aa) **rs9507279 (implicating *SPATA13*).**
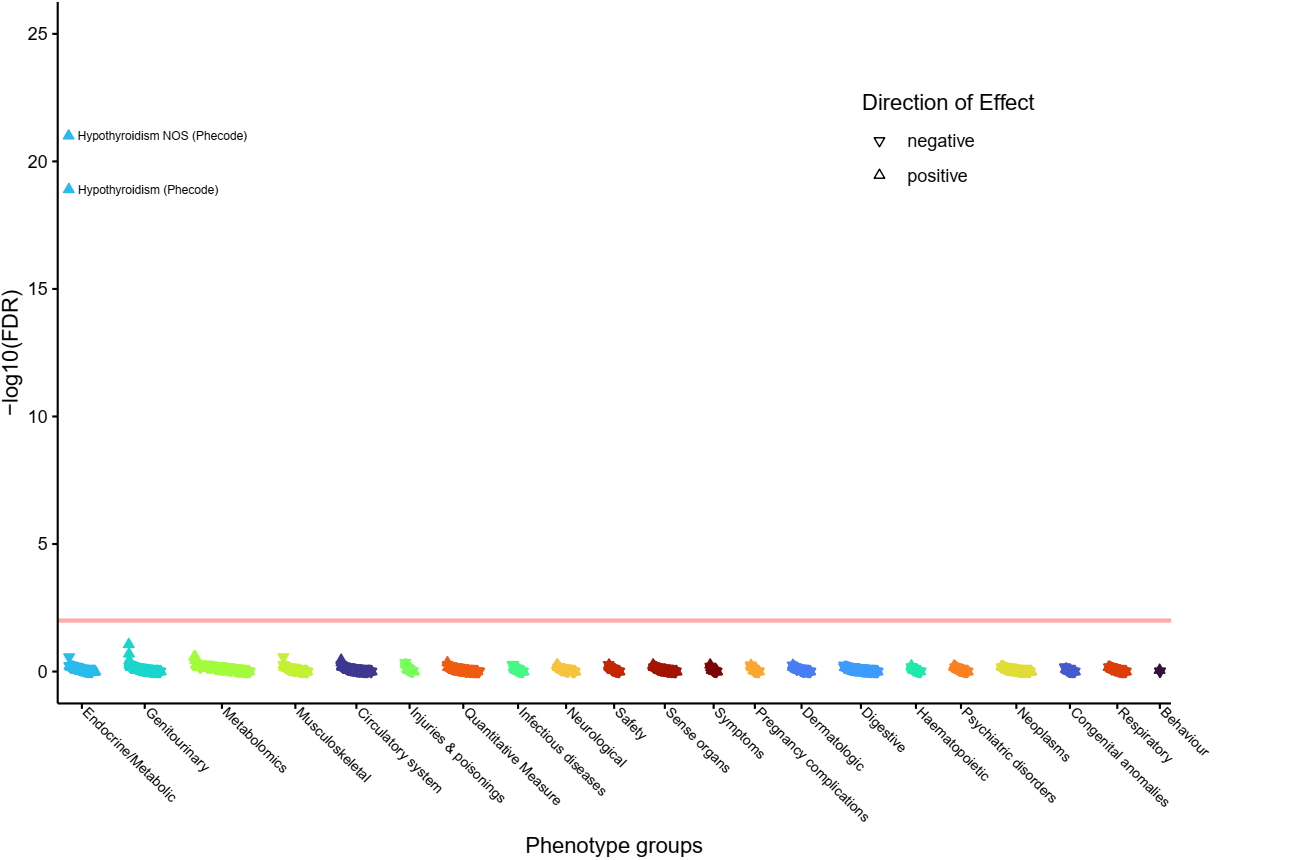

bb) **rs17364832 (implicating *SPATA13*).**
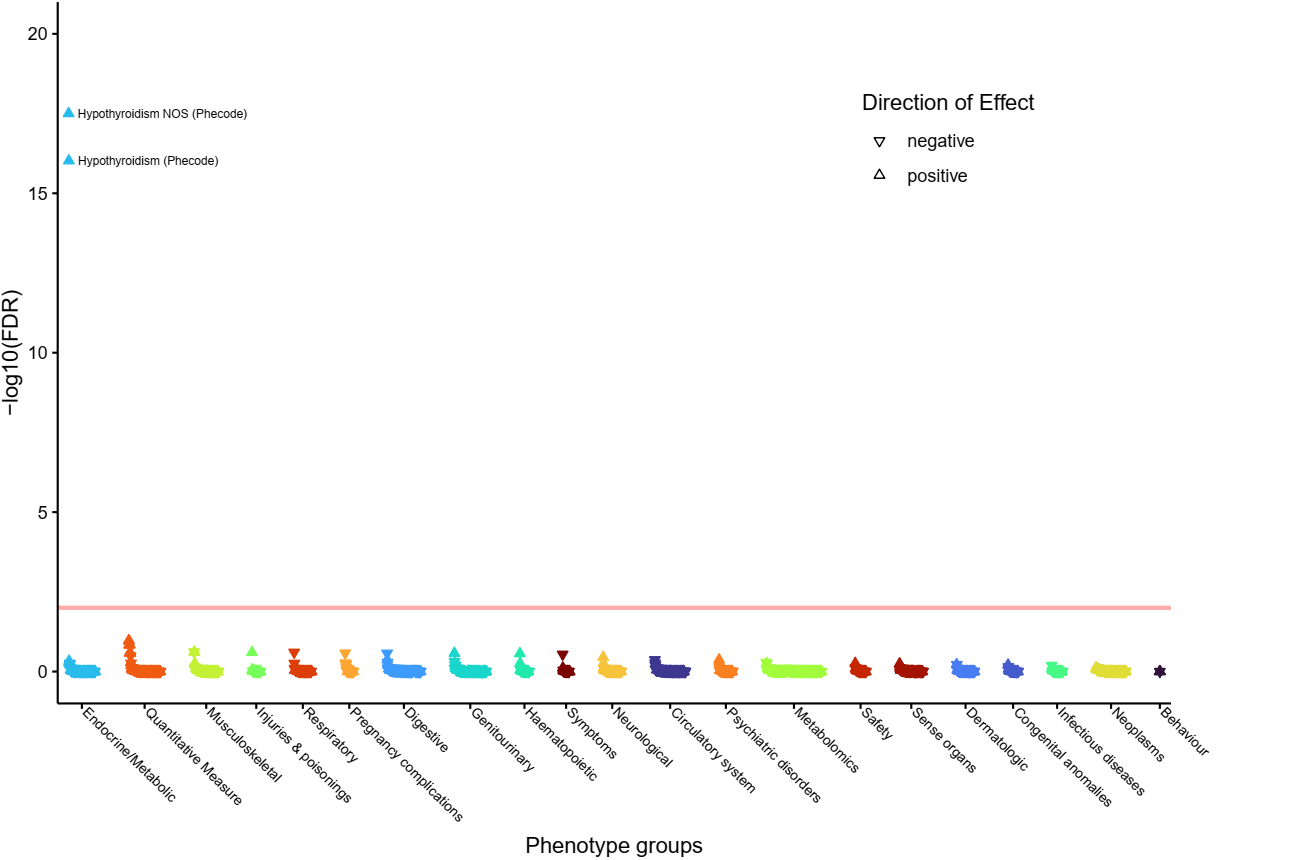

cc) **rs10113355 (implicating *SULF1* and *LOC100505739*).**
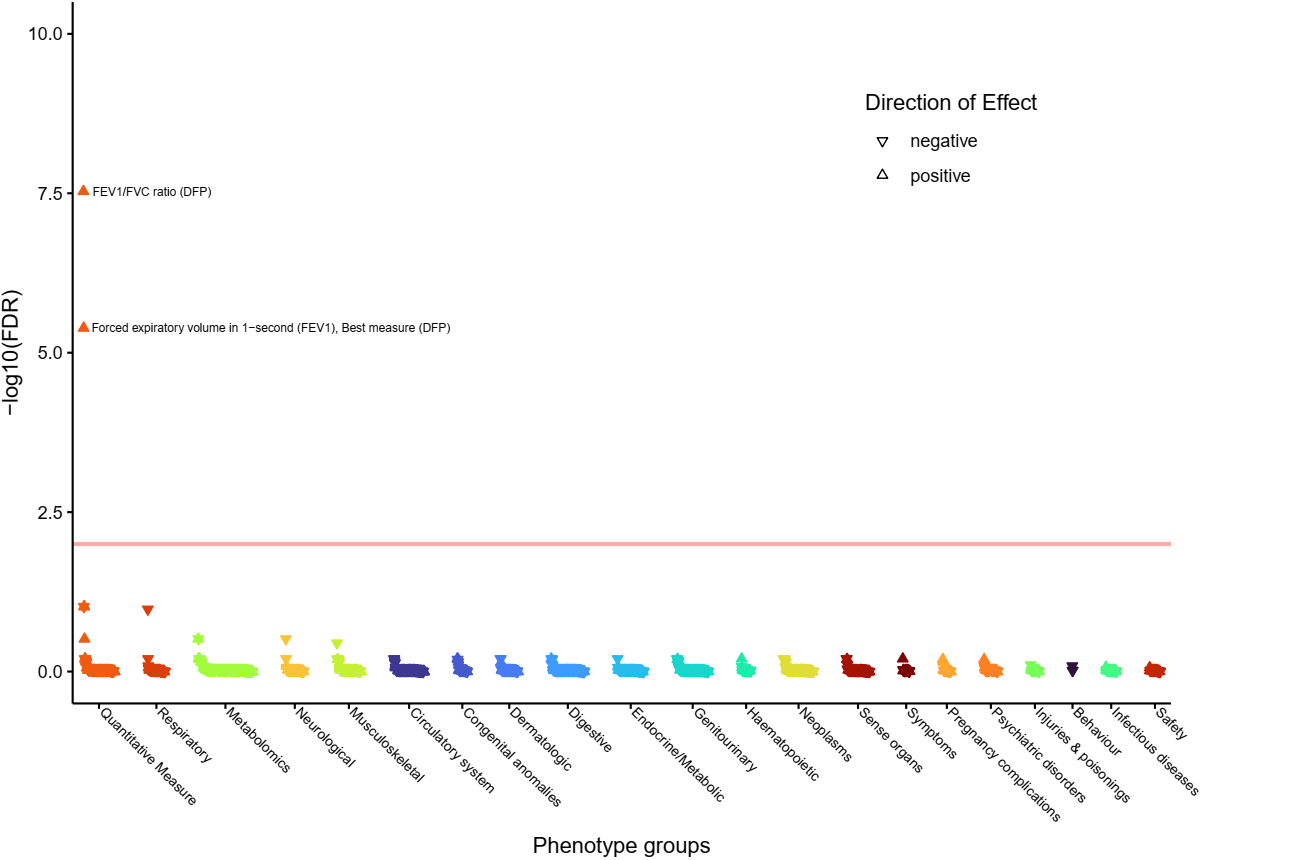

dd) **rs2069556 (implicating *TG* and *CCN4*).**
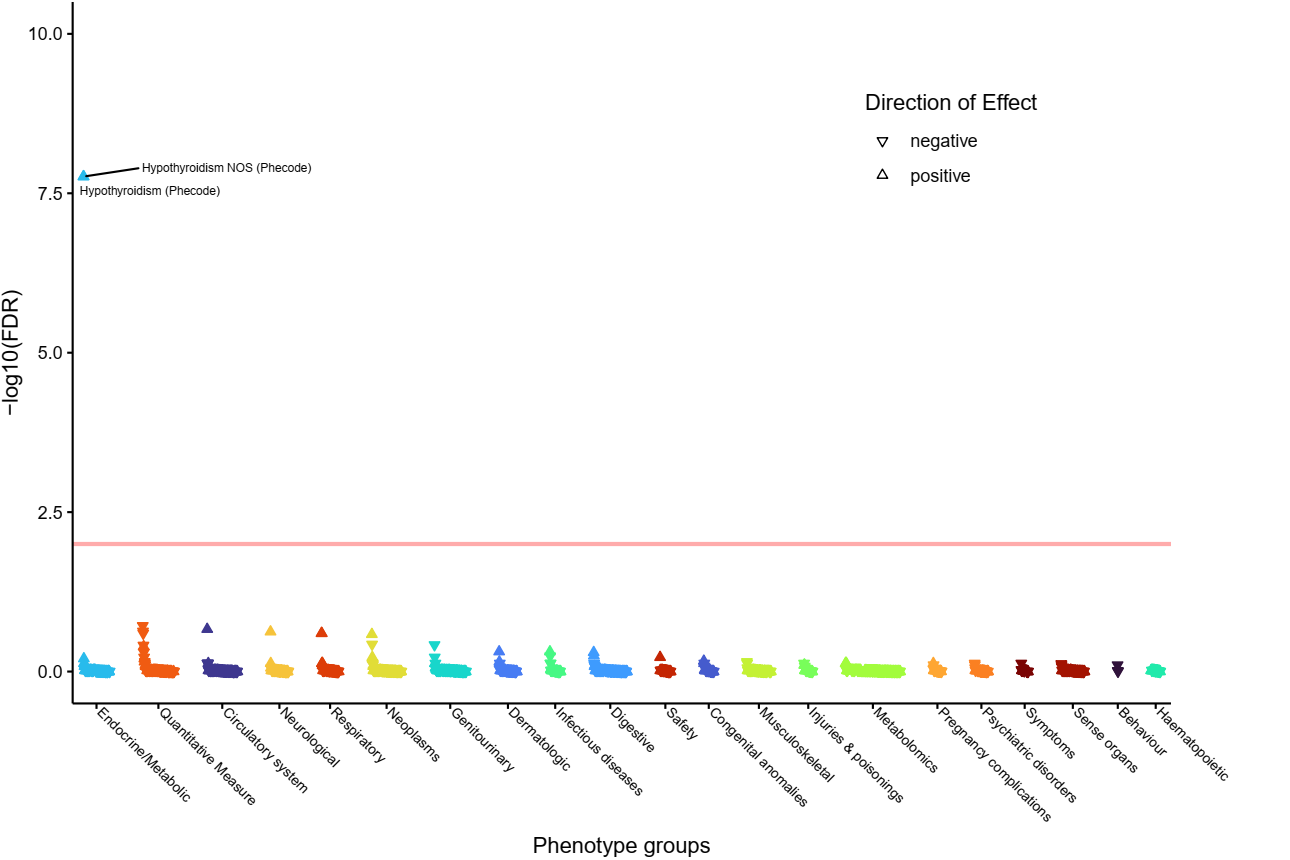

ee) **rs2069568 (implicating *TG* and *CCN4*).**
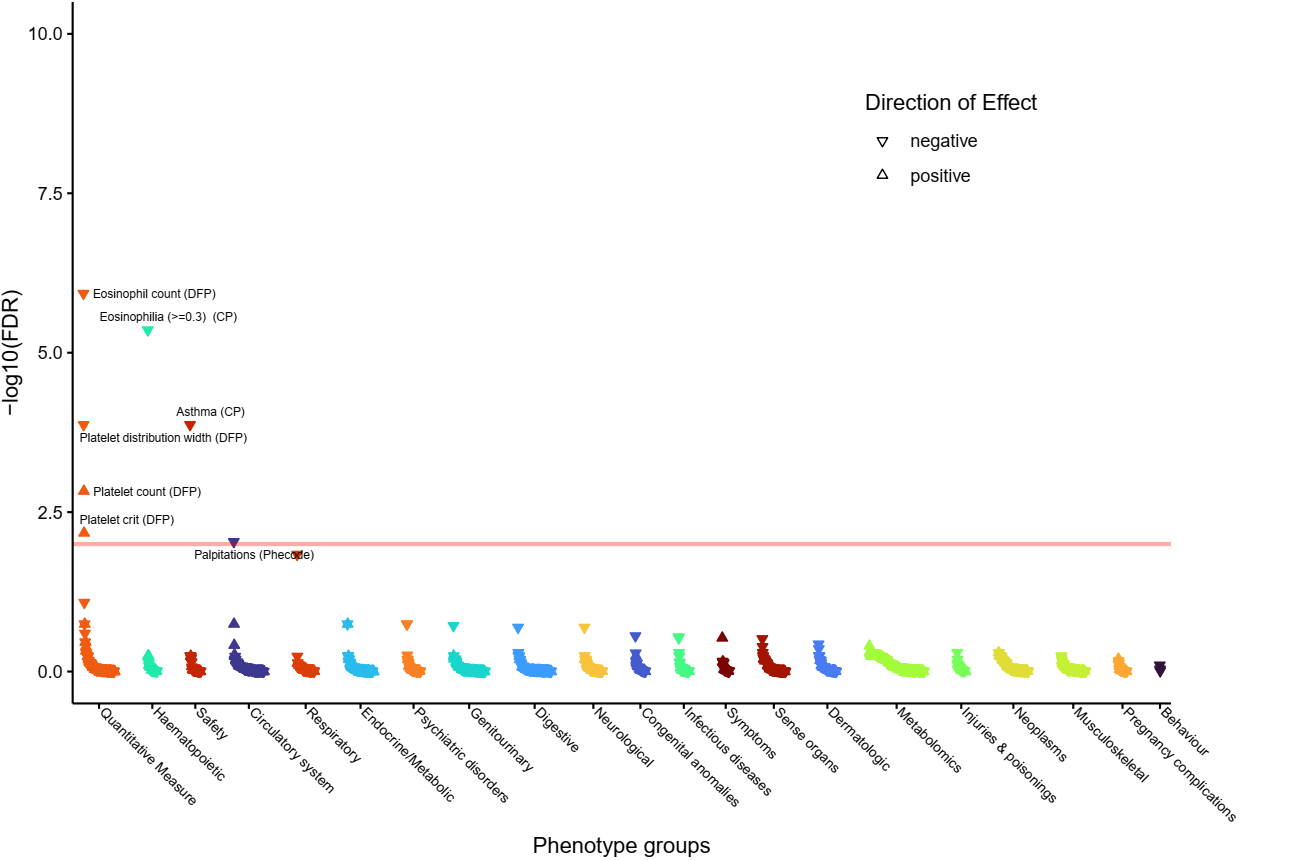

ff) **rs114322847 (implicating *TG* and *CCN4*).**
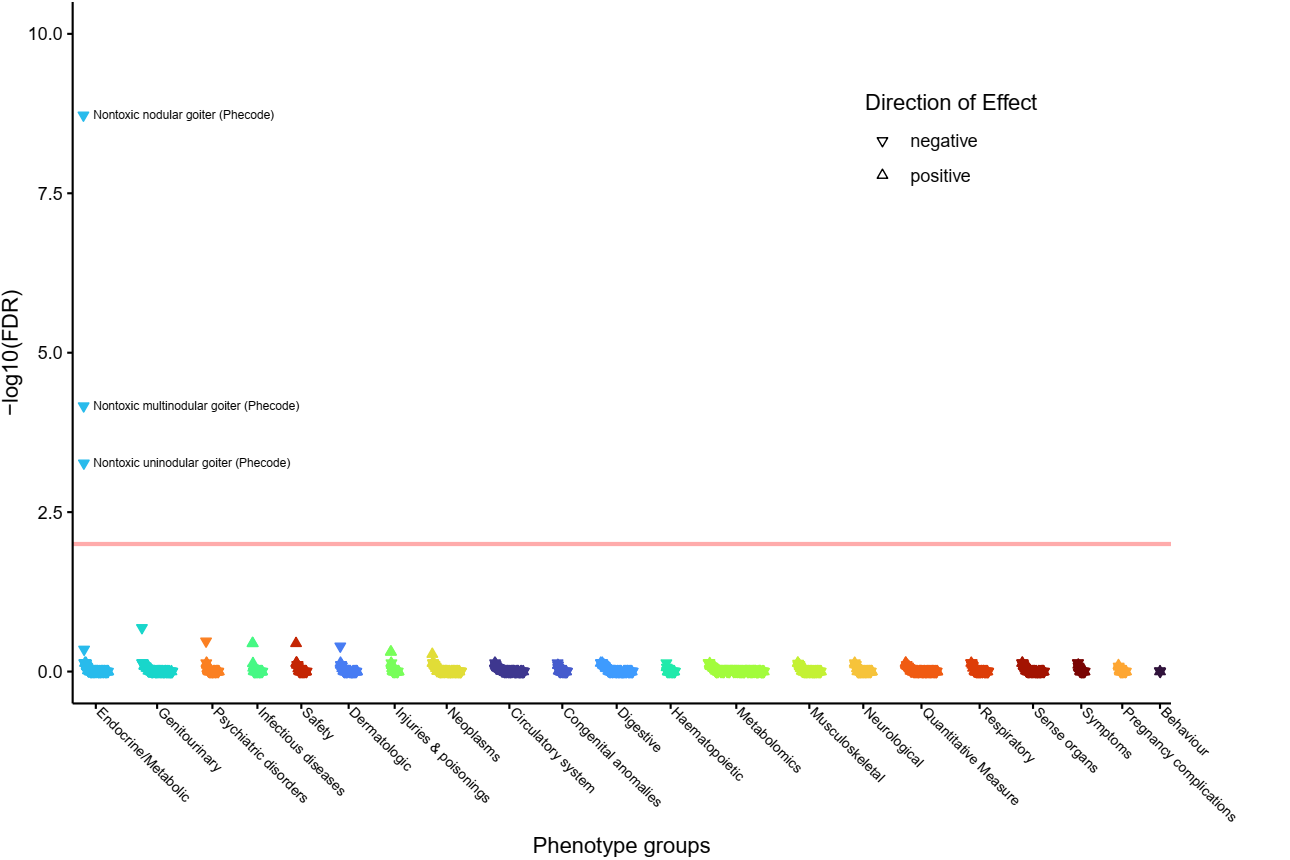

gg) **rs519709 (implicating *TGFB2* and *MIR548F3*).**
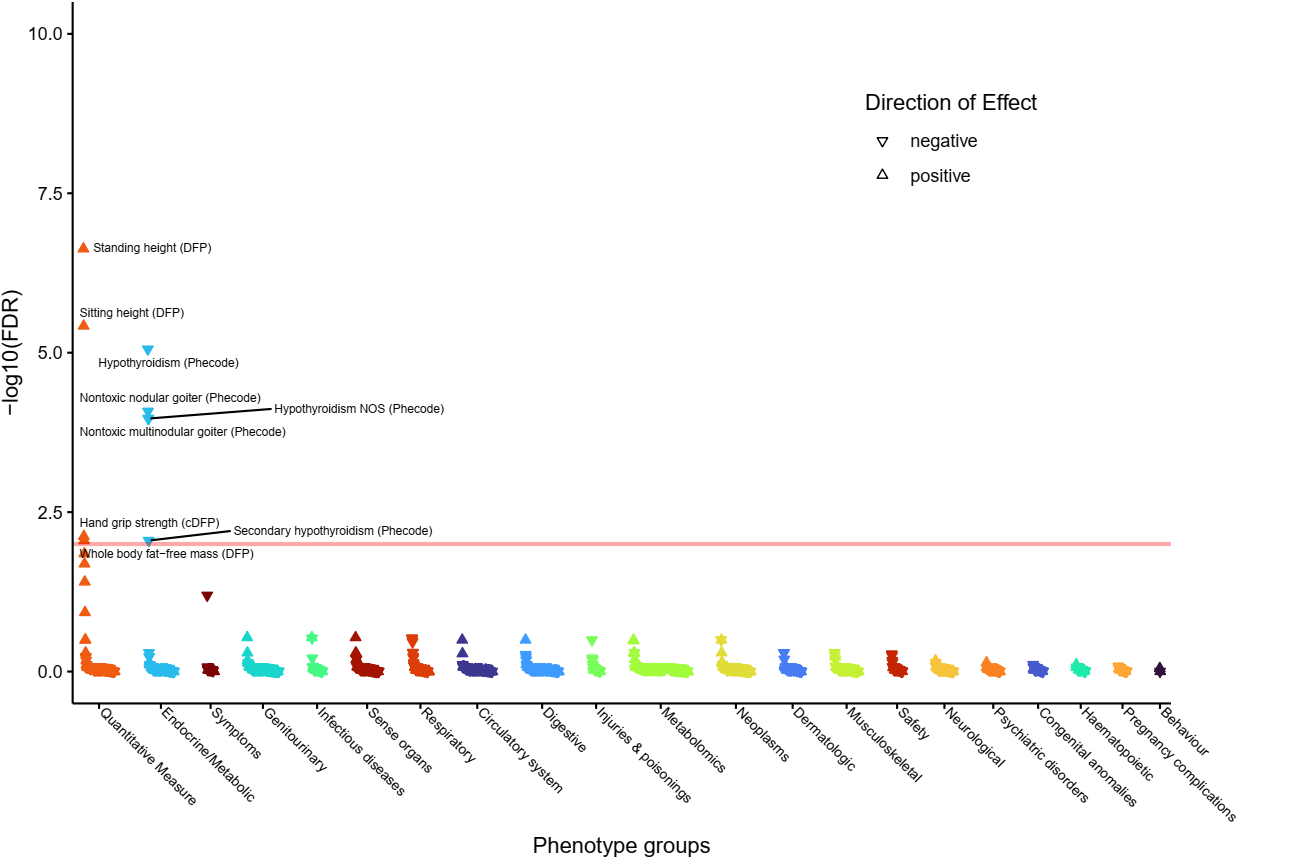

hh) **rs11694732 (implicating *TPO*).**
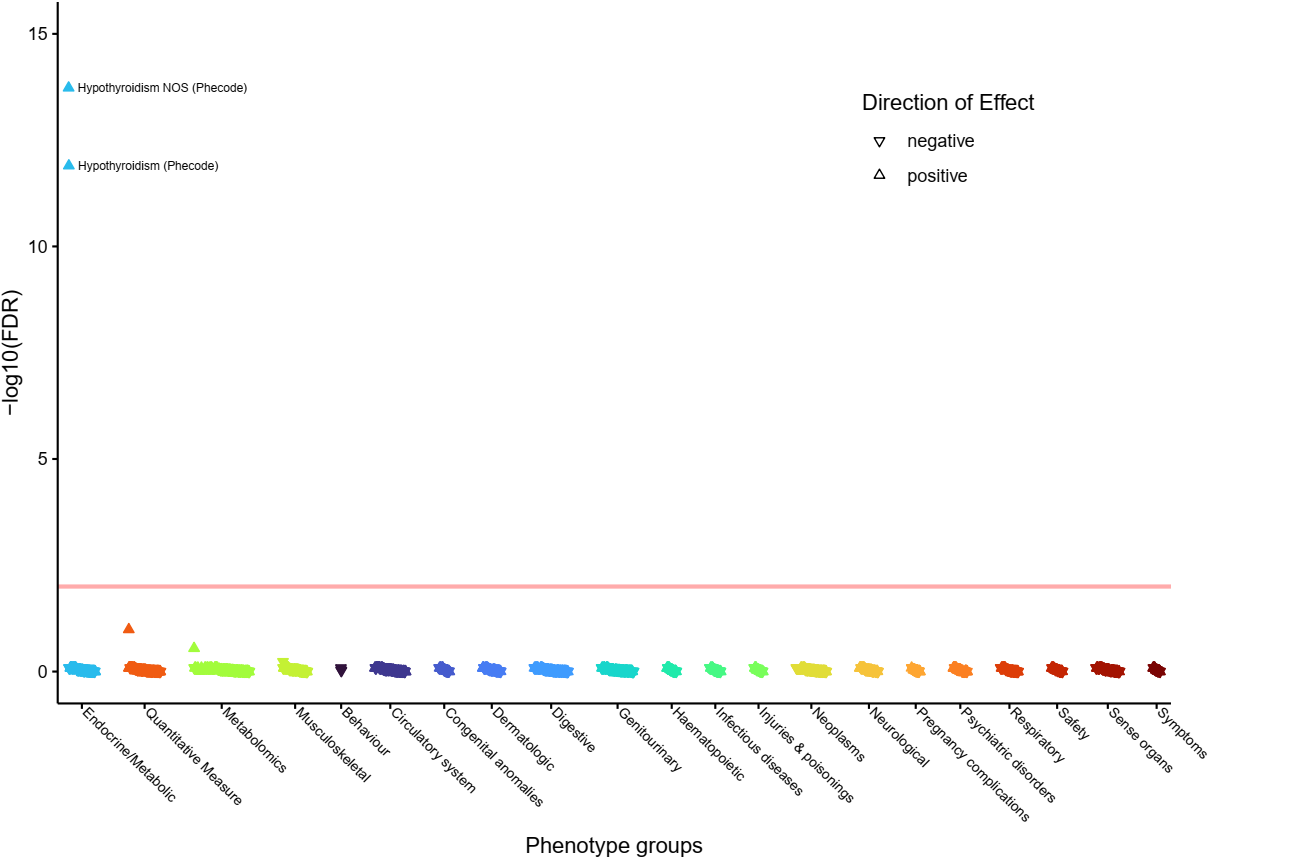

ii) **rs3742721 (implicating *TSHR* and *CEP128*).**
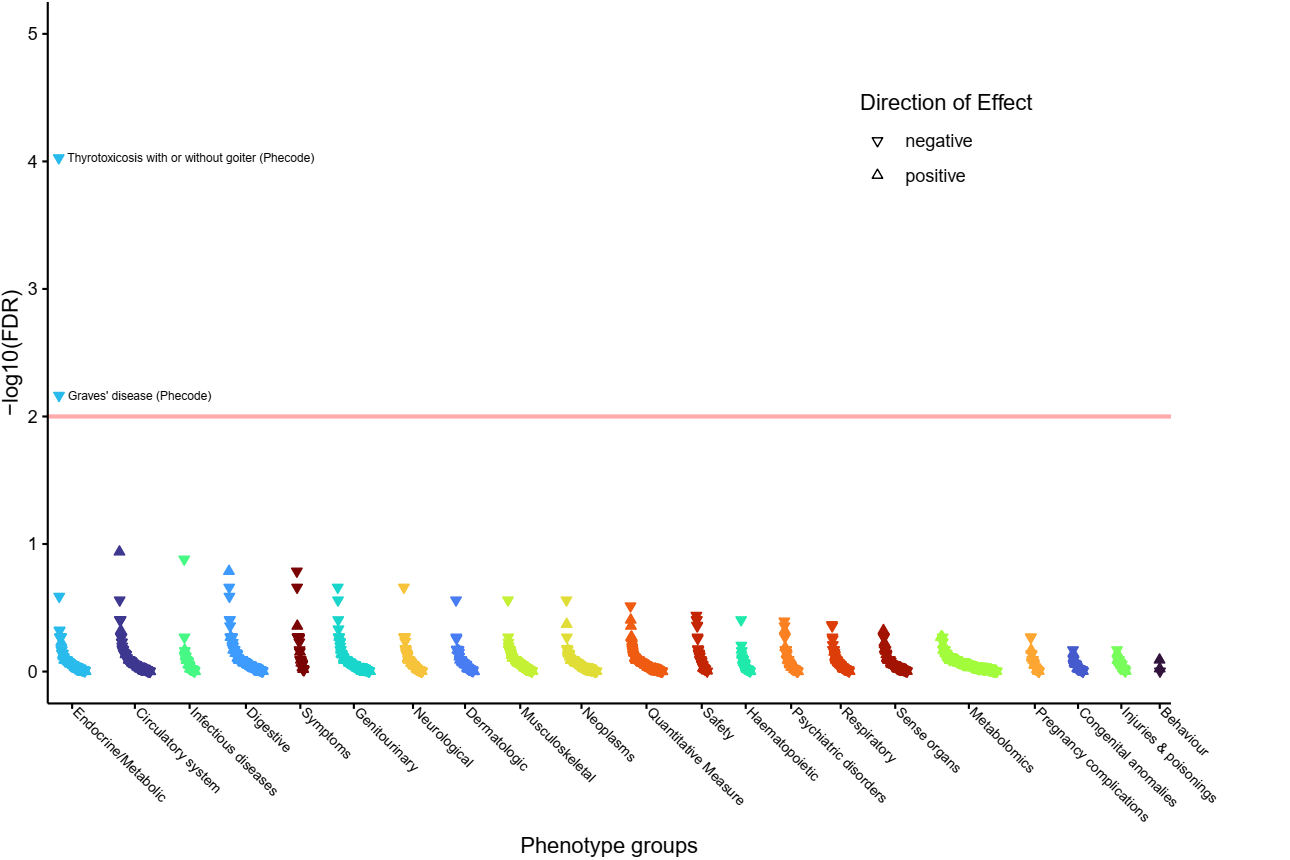

jj) **rs6574611 (implicating *TSHR* and *CEP128*).**
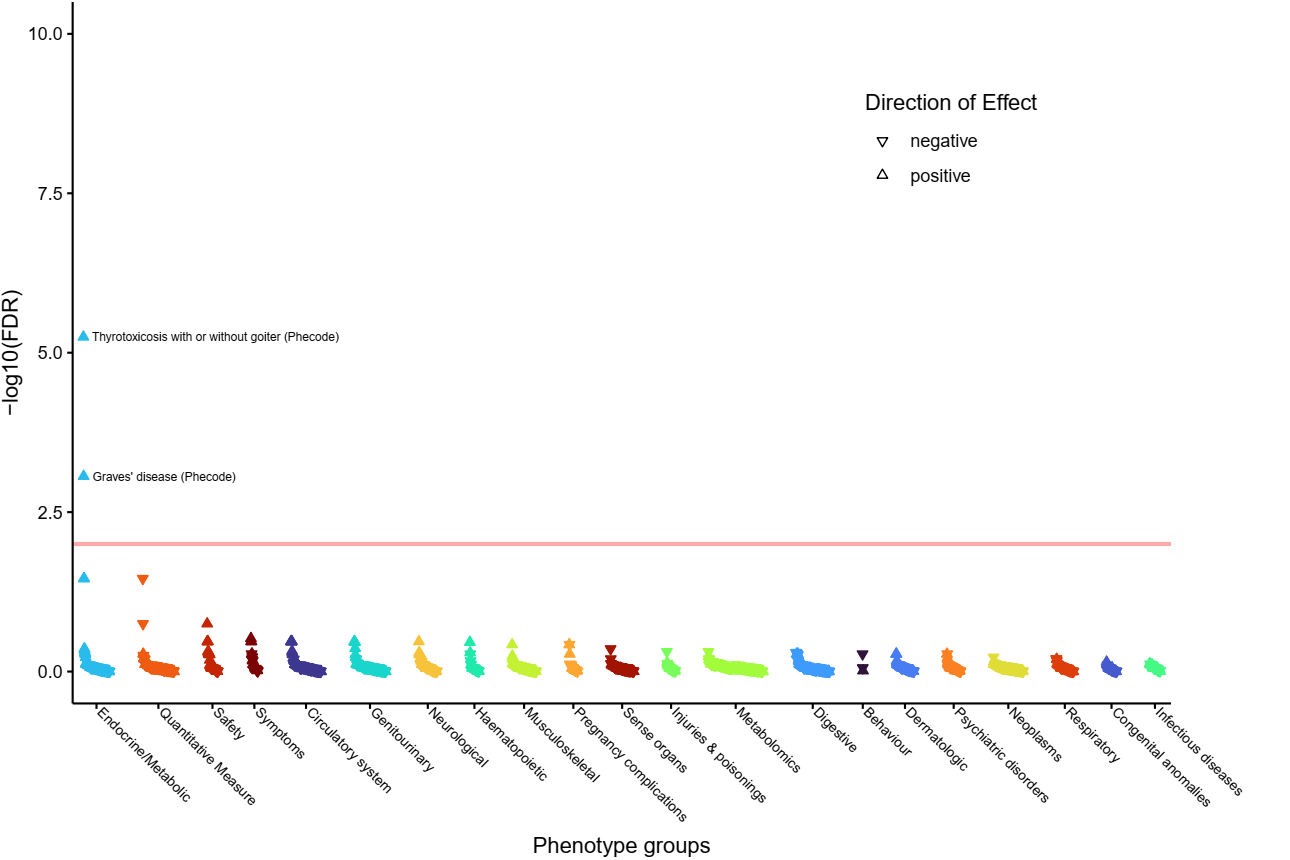

kk) **rs2110696 (implicating *TSHR*).**
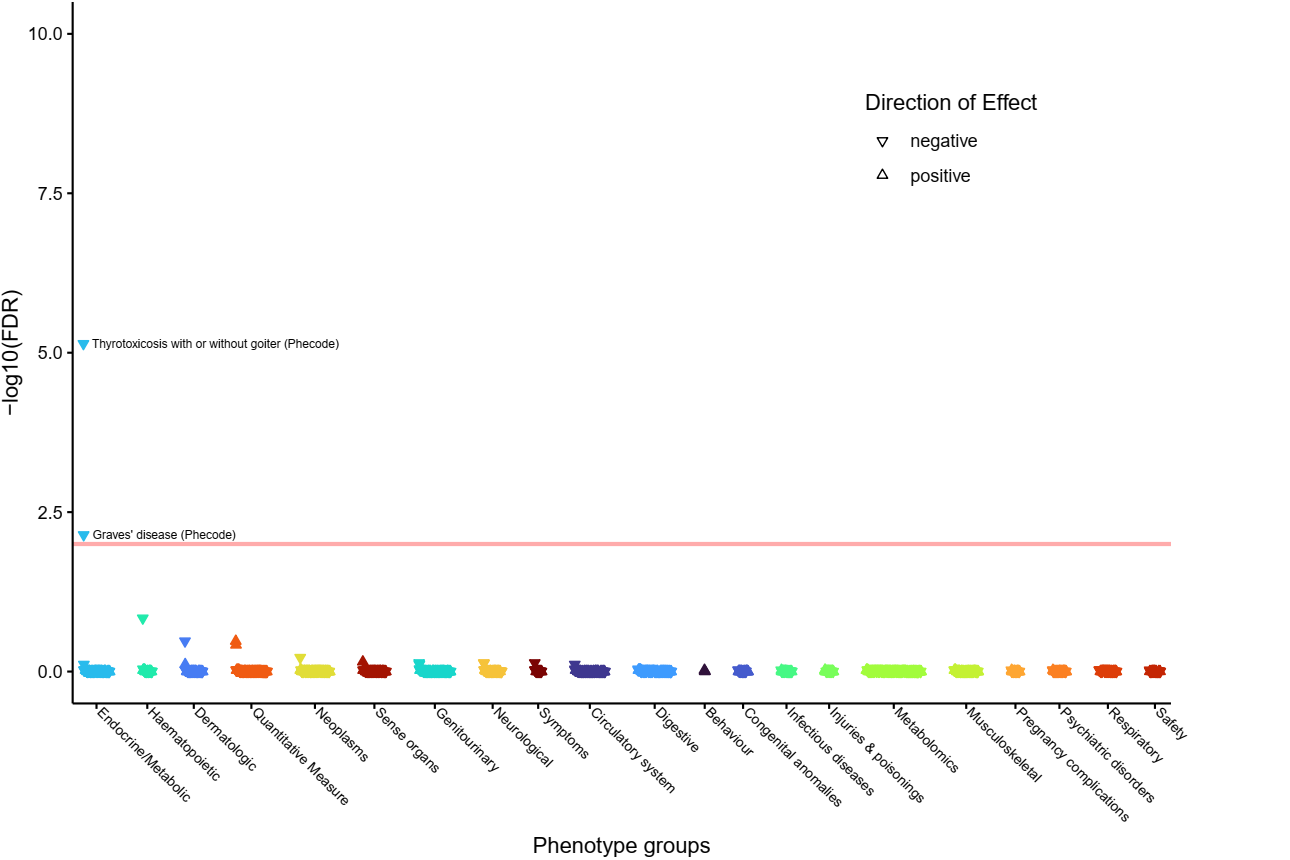

ll) **rs1802288 (implicating *TSPAN6*).**
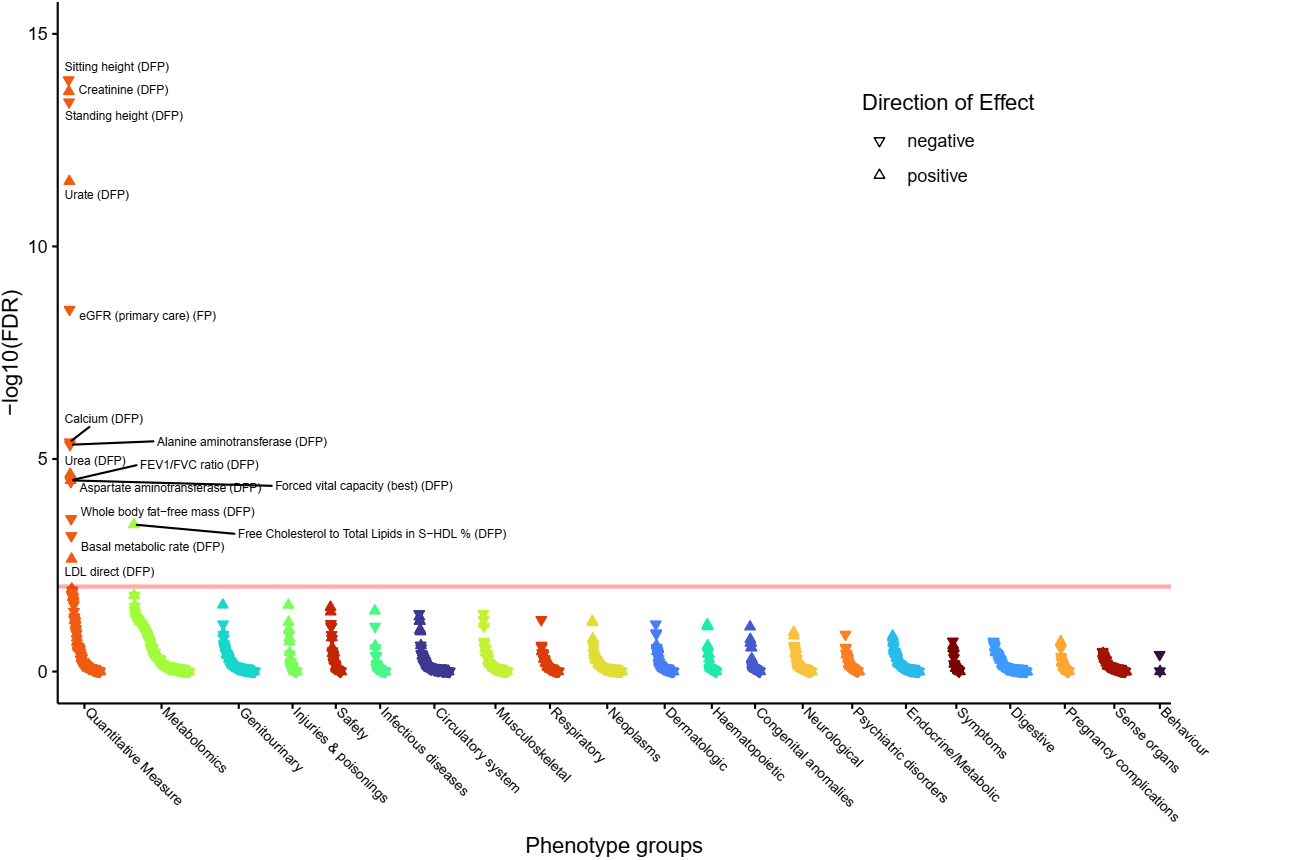

mm) **rs3755972 (implicating *VEGFC*).**

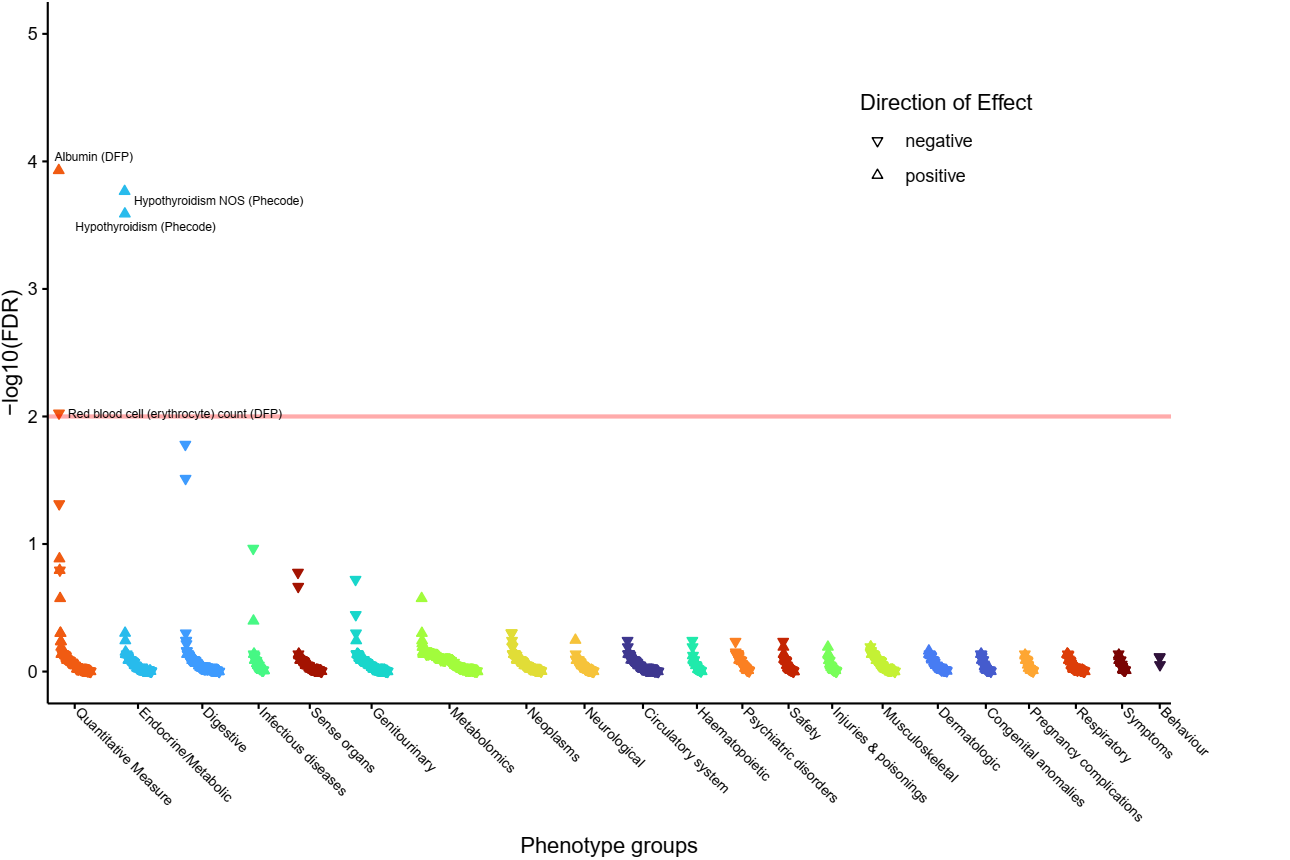

Supplementary Figure 2**:** 112 genes prioritised by two or more variant-to-gene criteria. The first seven columns indicate that at least one variant implicates the corresponding gene via the evidence for that column. The remaining seven columns indicate the strength of association of the most significant variant implicating the corresponding gene as causal with respect to the TSH increasing allele, such that shades of blue represent associations with the other thyroid phenotypes that have the same direction of effect as the TSH association and shades of red represent an opposite direction of effect to the TSH association.

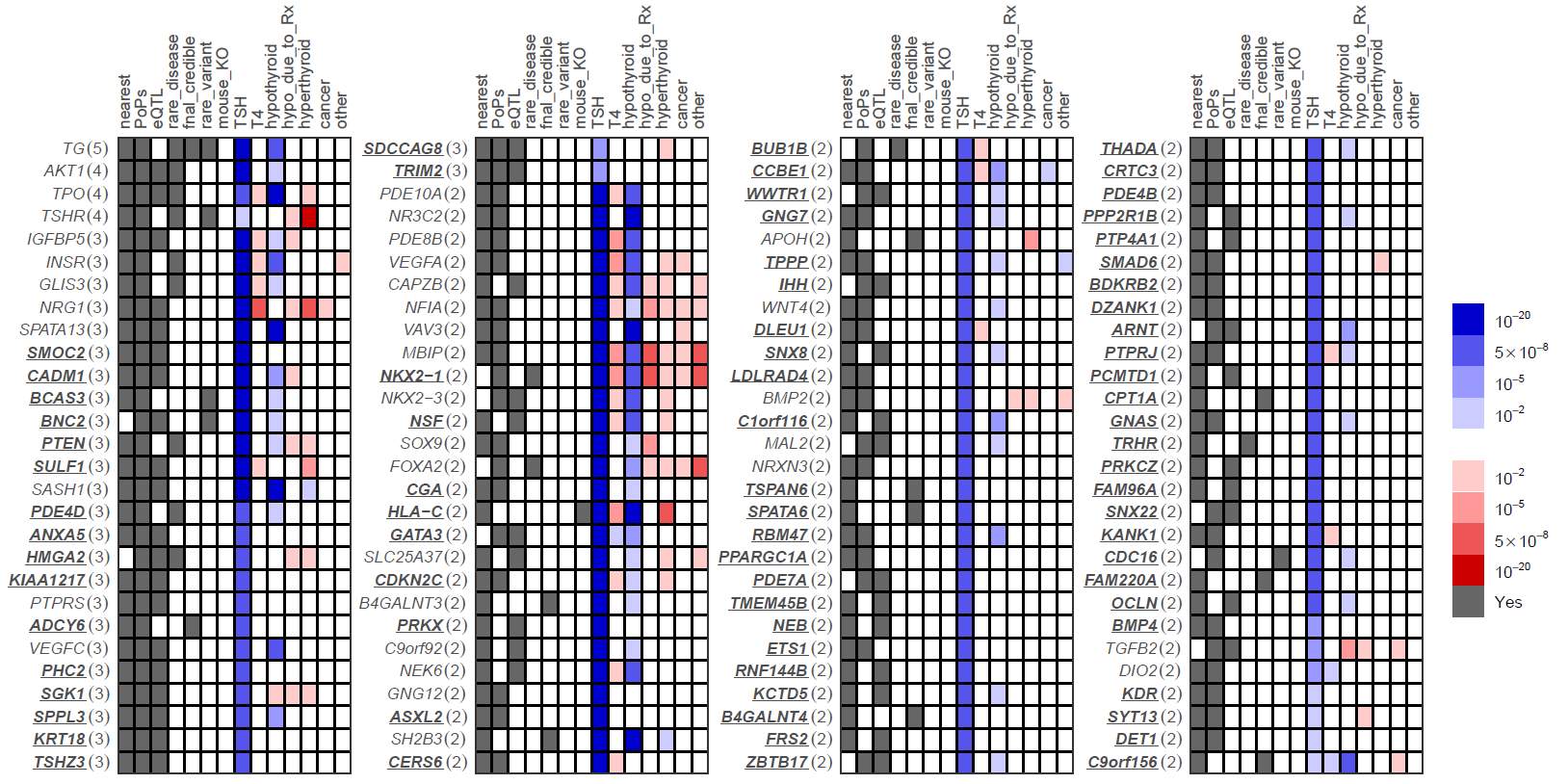

Supplementary Figure 3: Pathway-specific polygenic score phenome-wide association studies (PheWAS)

a) **cAMP (KEGG) signalling pathway**

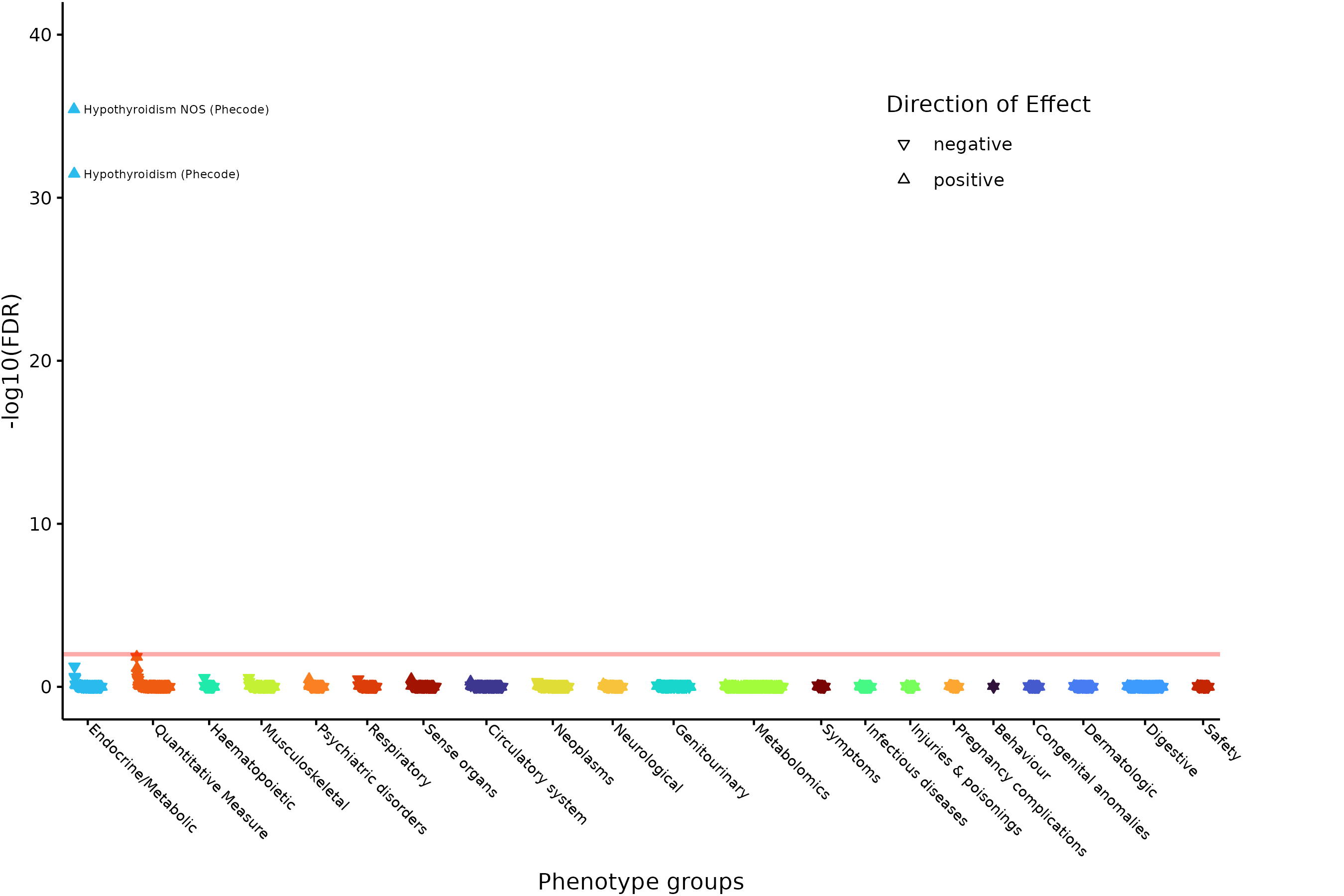

b) **ADORA2B mediated anti-inflammatory cytokines production pathway.**

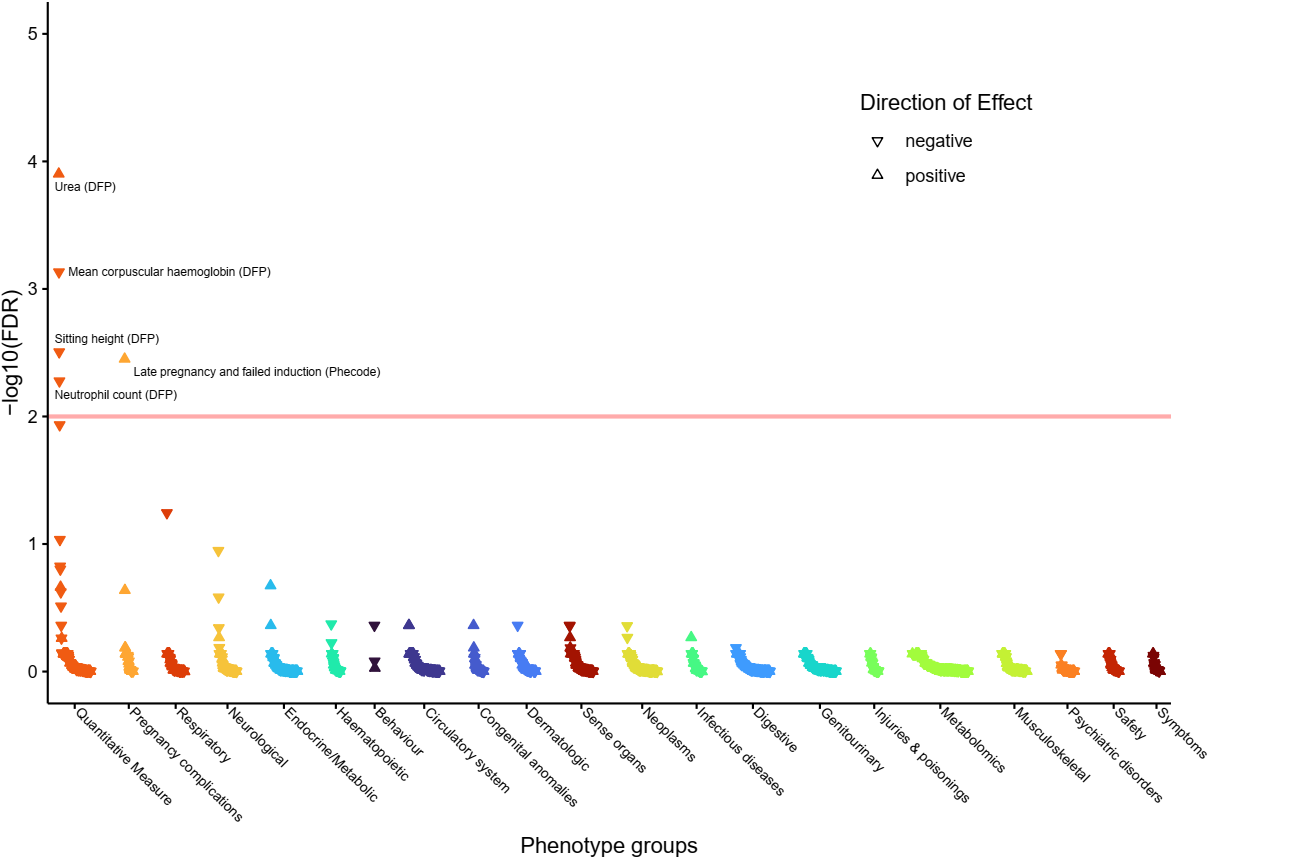

c) **Apoptosis-related network due to altered Notch3 in ovarian cancer pathway.**
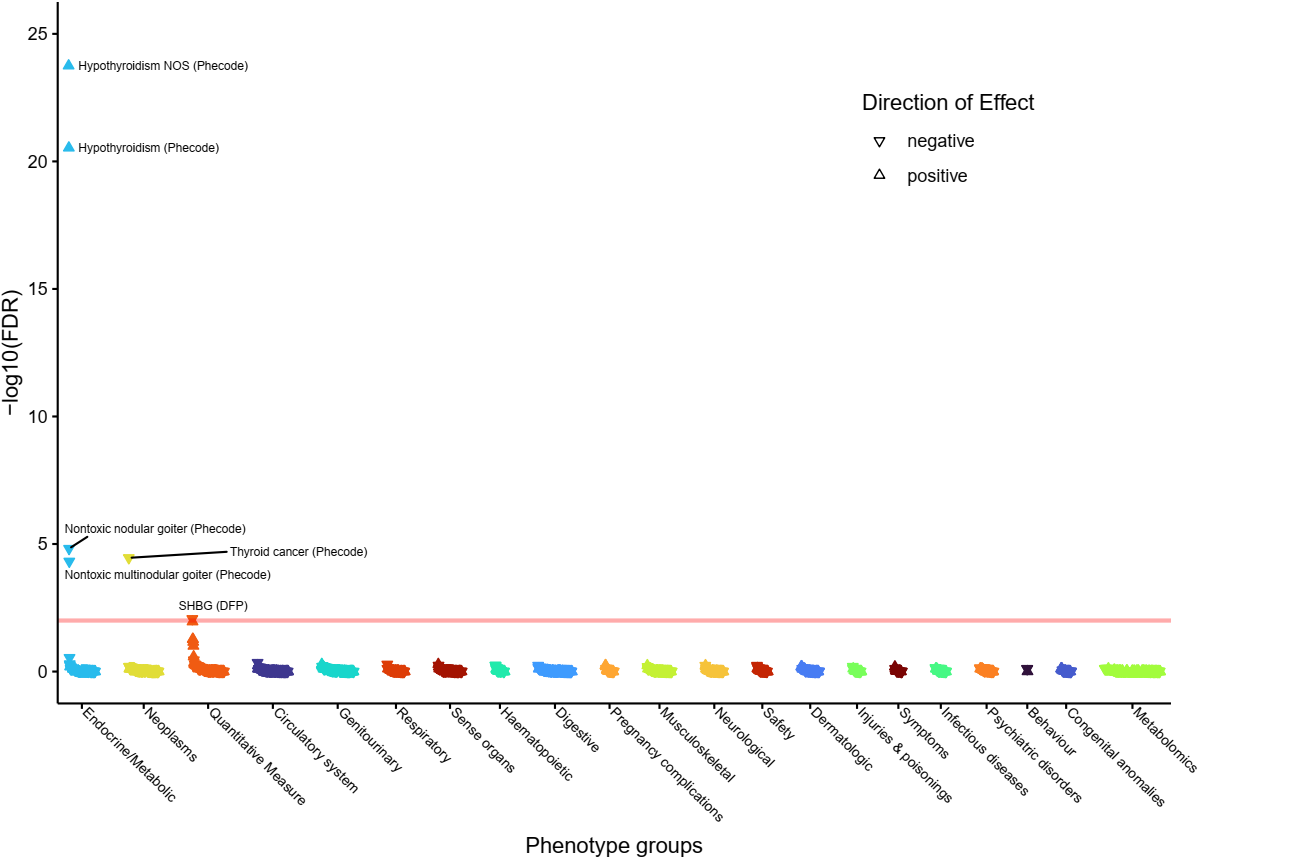

d) **Factors and pathways affecting insulin-like growth factor (IGF1)-Akt signaling pathway.**
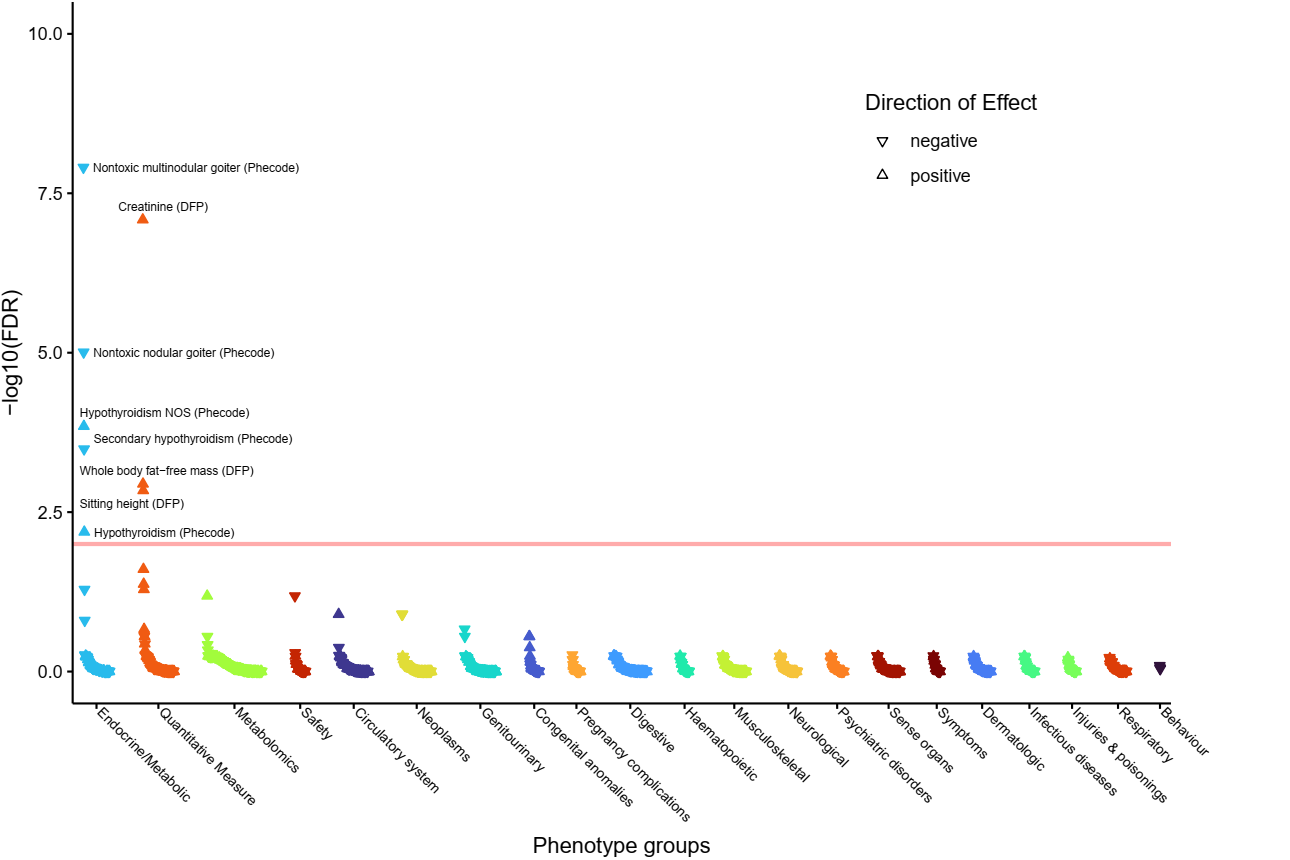

e) **FGFR3 signaling in chondrocyte proliferation and terminal differentiation pathway.**
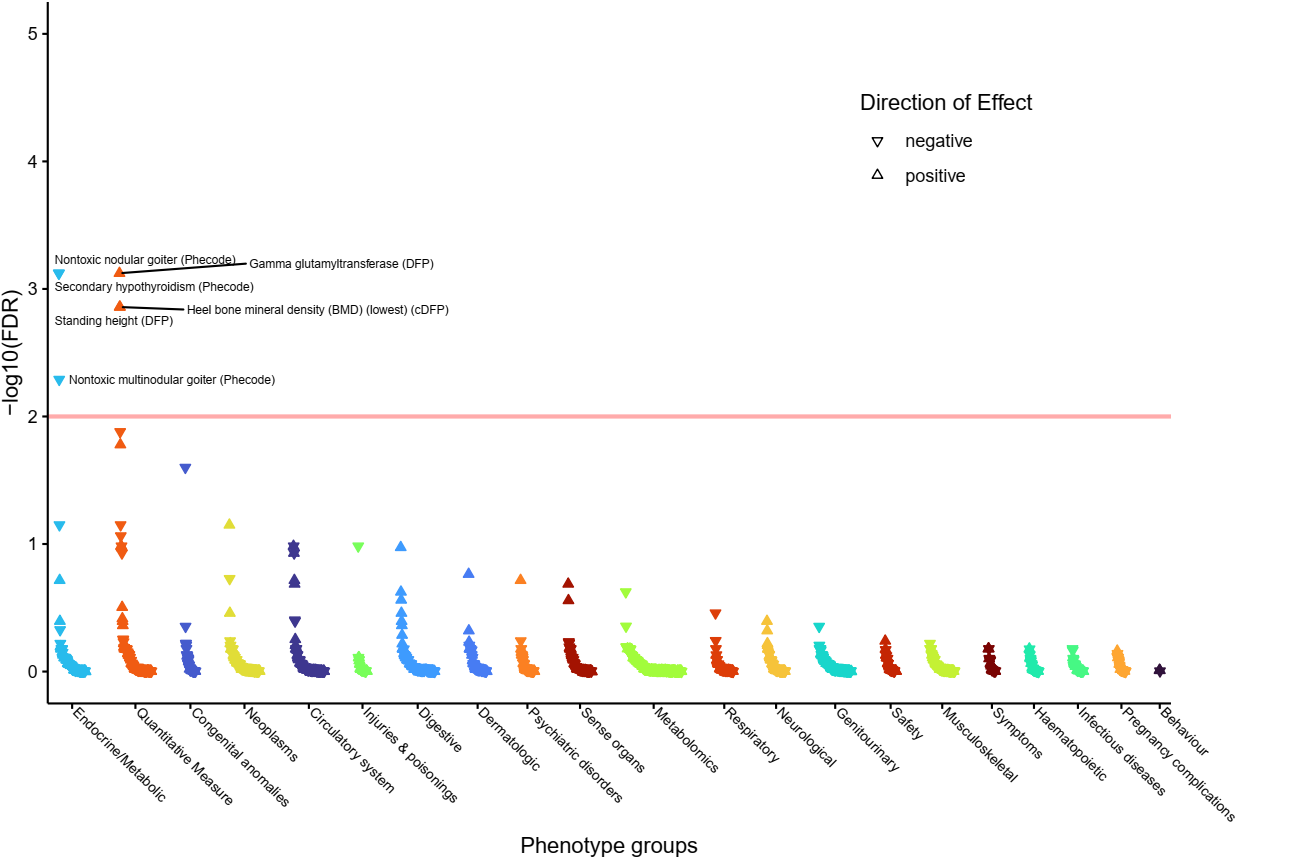

f) **Focal Adhesion-PI3K-Akt-mTOR-signaling pathway.**
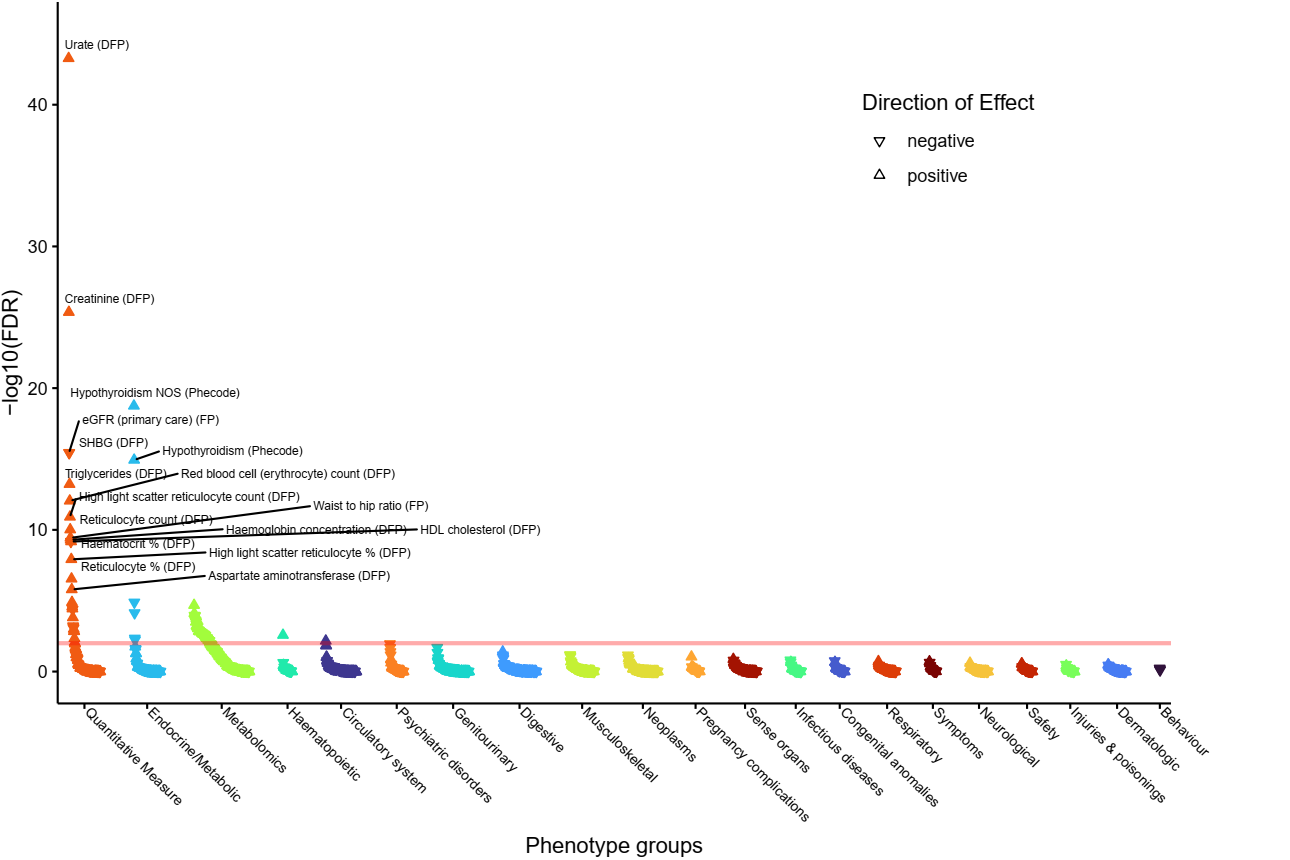

g) **G Protein Signaling Pathways pathway.**
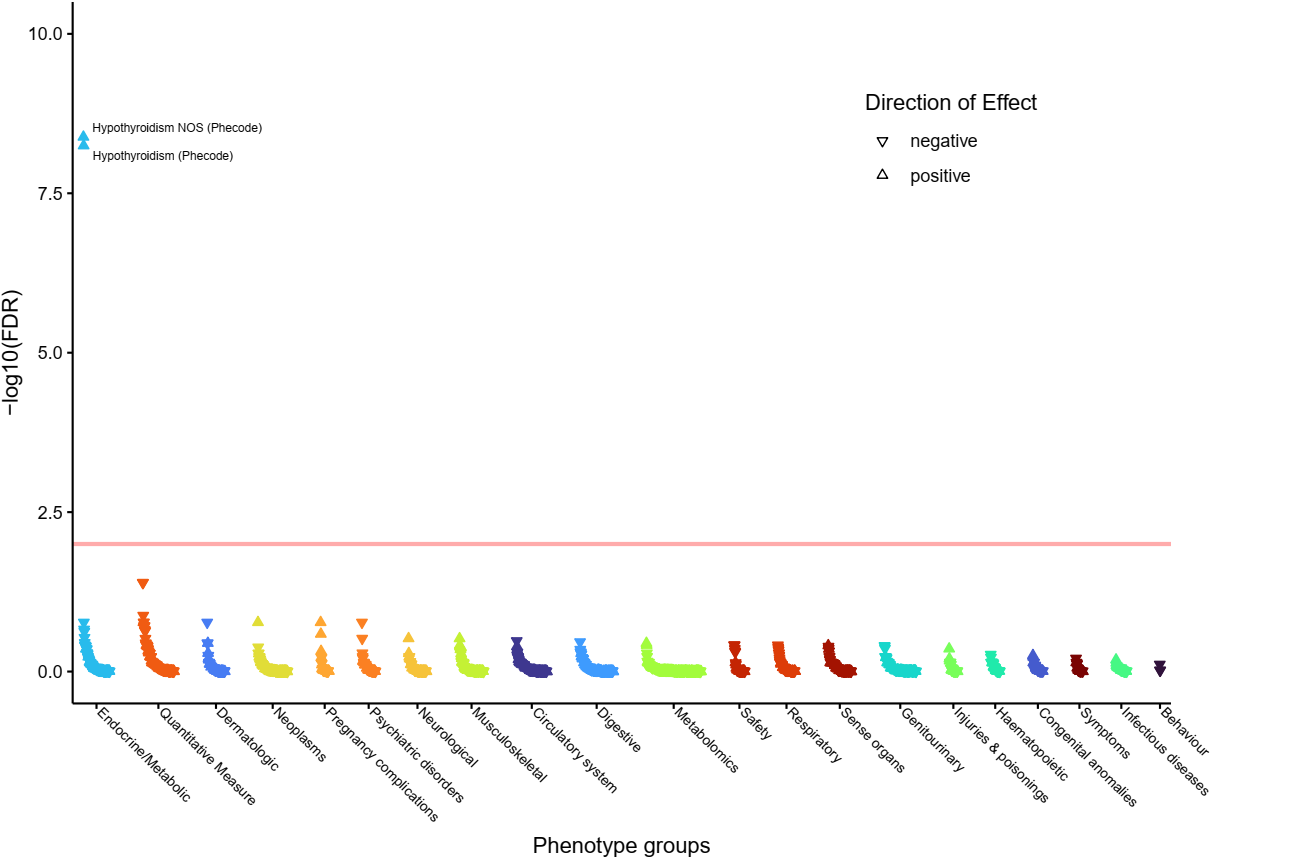

h) **Heart Development pathway.**
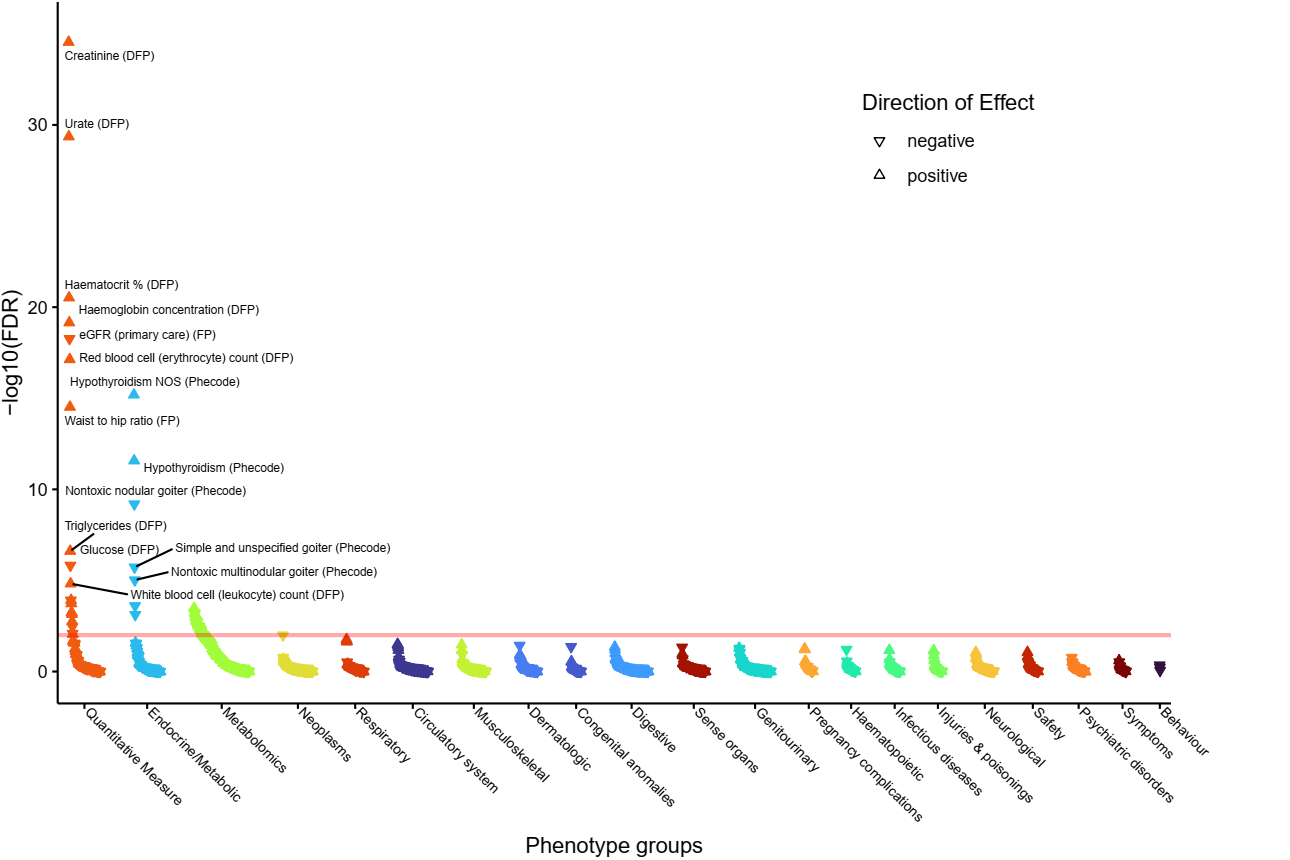

i) **Hemostasis pathway.**
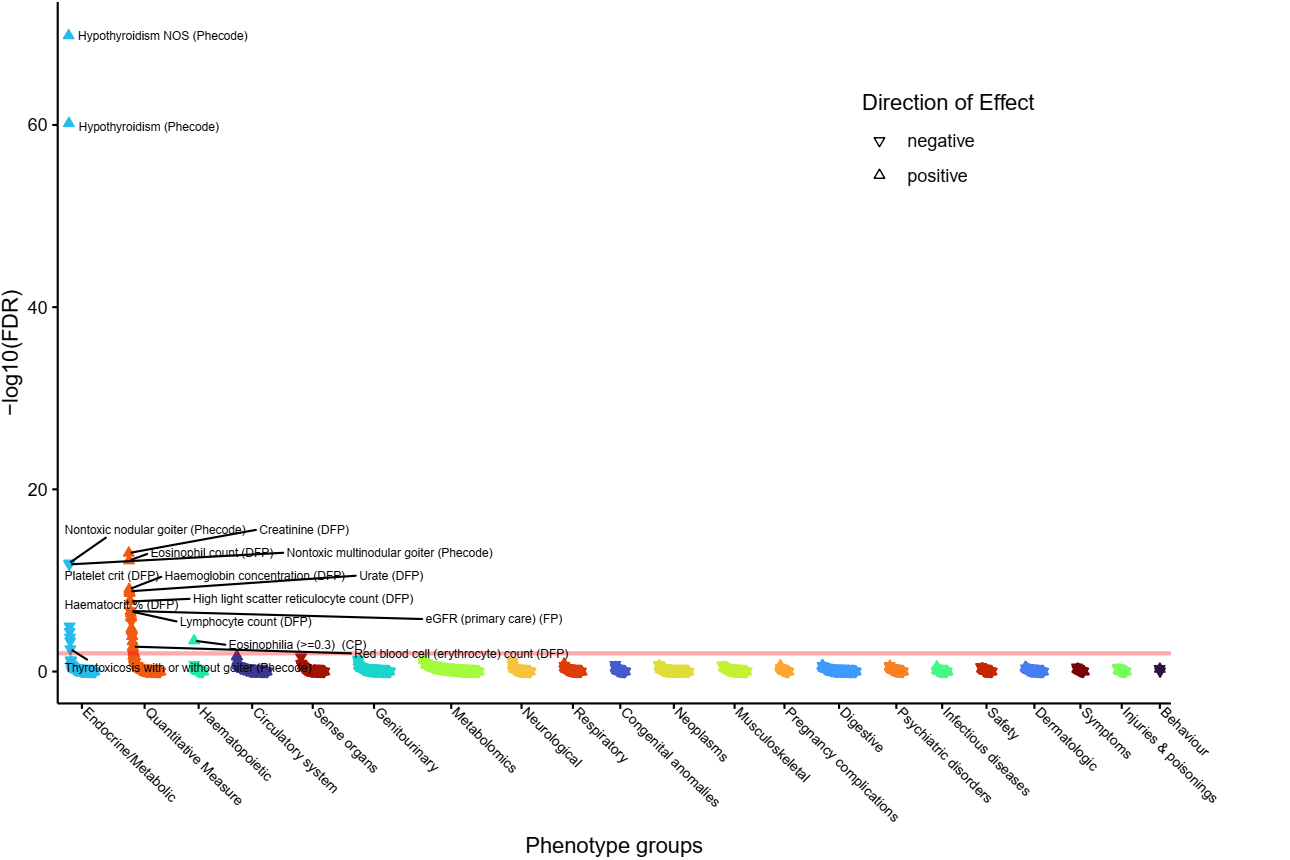

j) **Human T-cell leukemia virus 1 infection - Homo sapiens (human) pathway.**
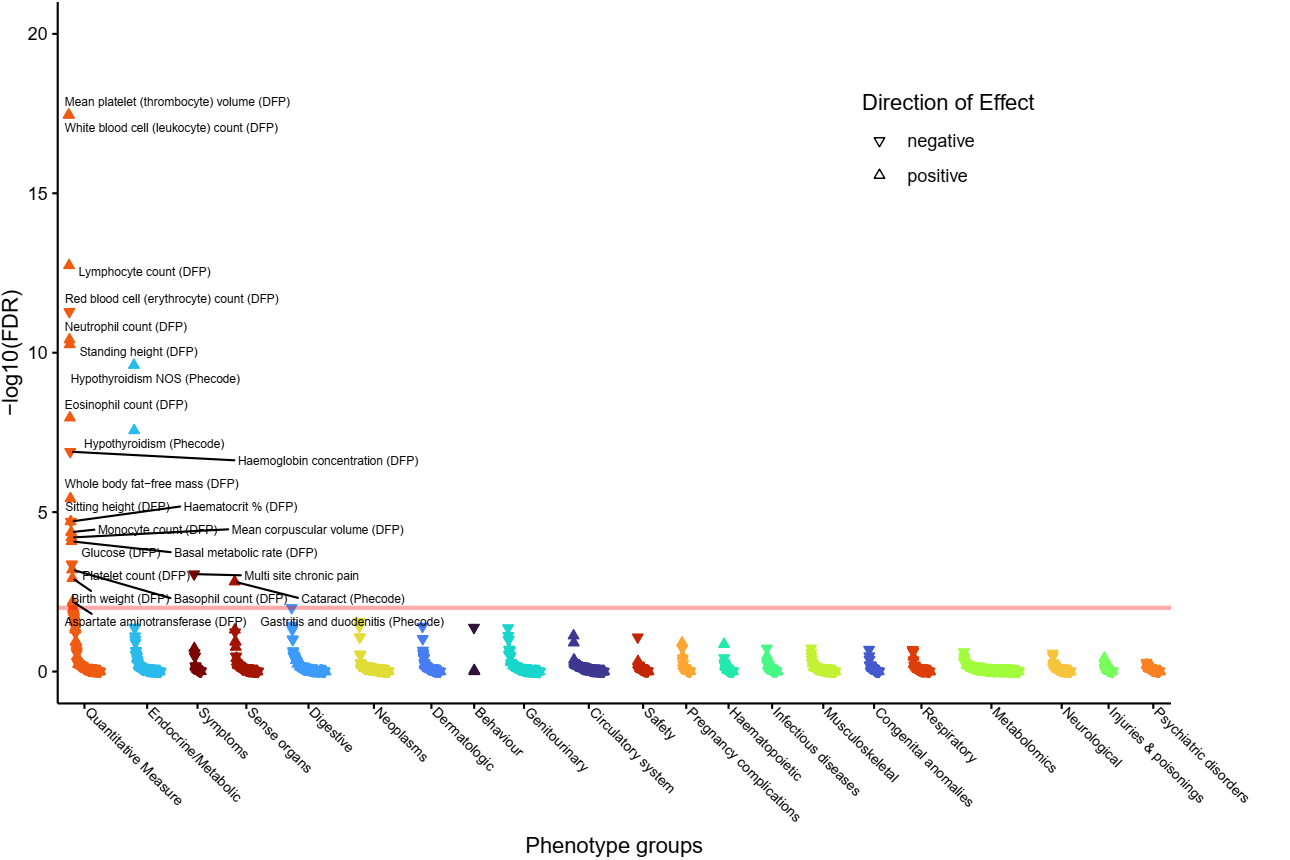

k) **Intracellular Signalling Through FSH Receptor and Follicle Stimulating Hormone pathway.**
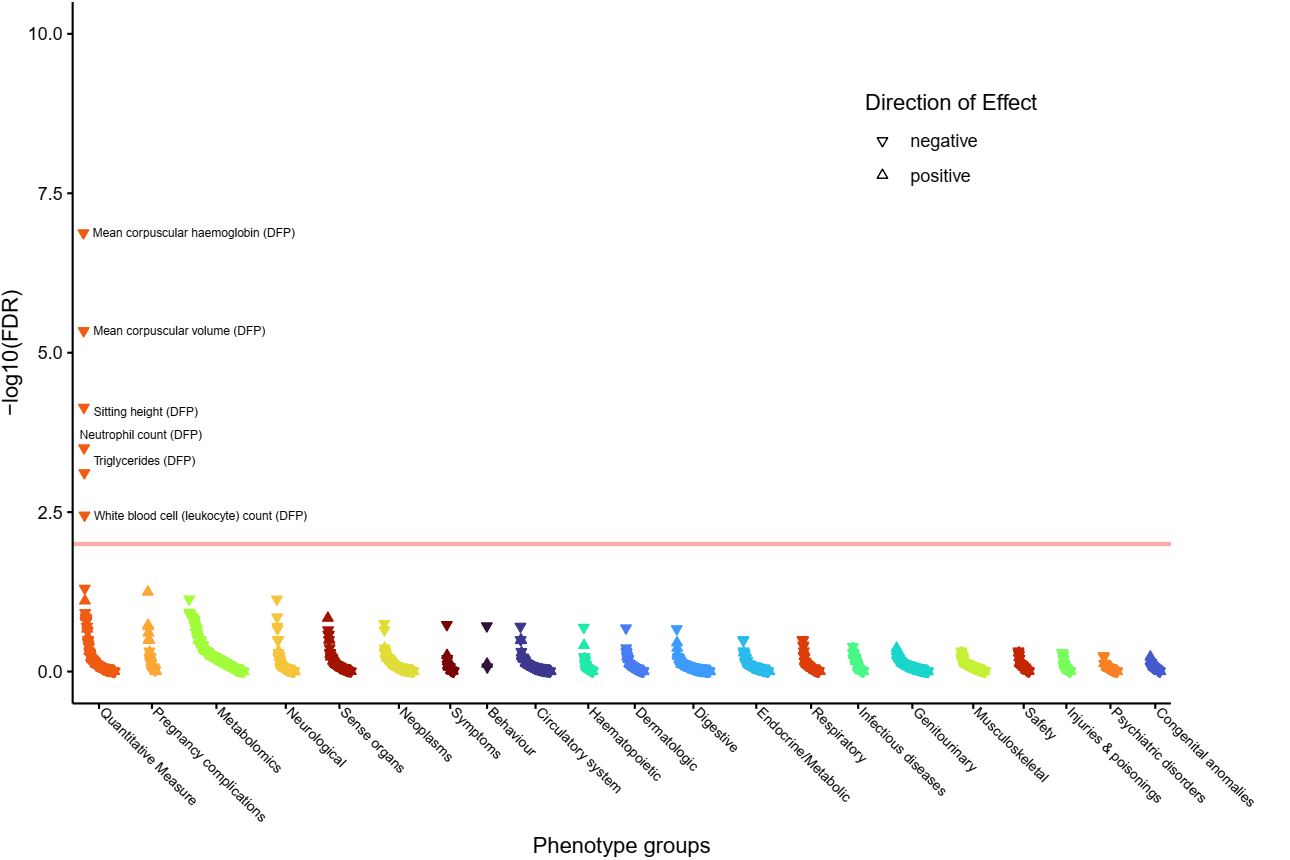

l) **Morphine addiction - Homo sapiens (human) pathway.**
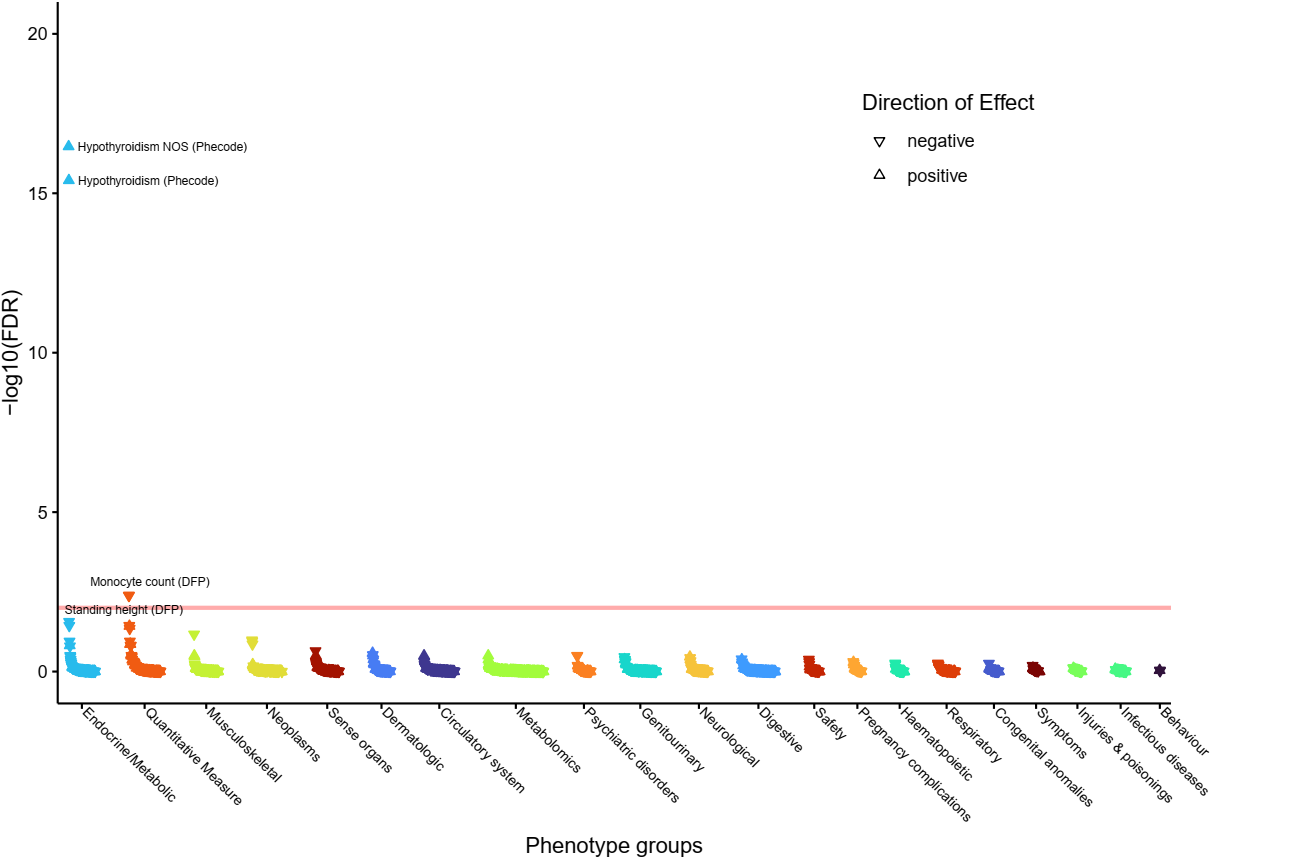

m) **Myometrial relaxation and contraction pathways.**
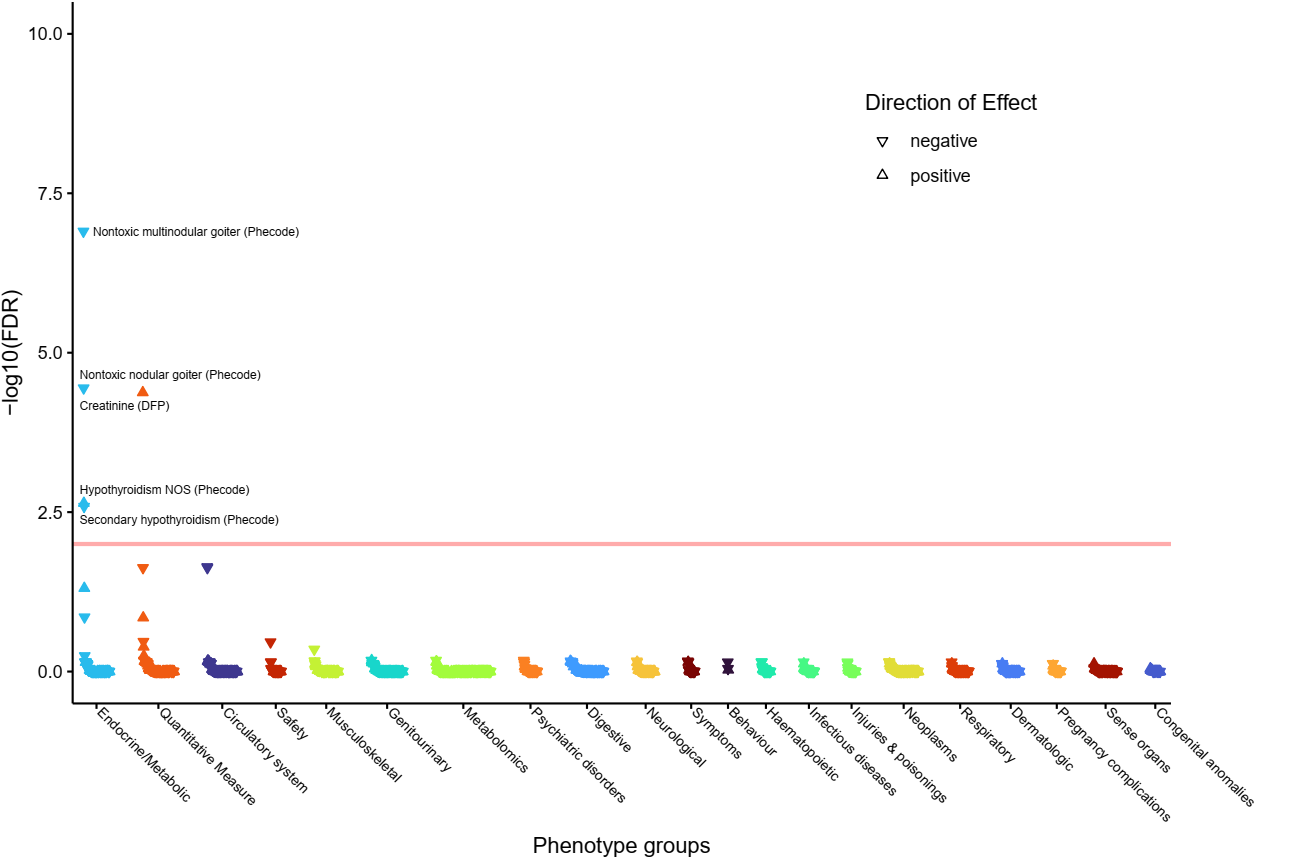

n) **Pathways in cancer - Homo sapiens (human) pathway.**
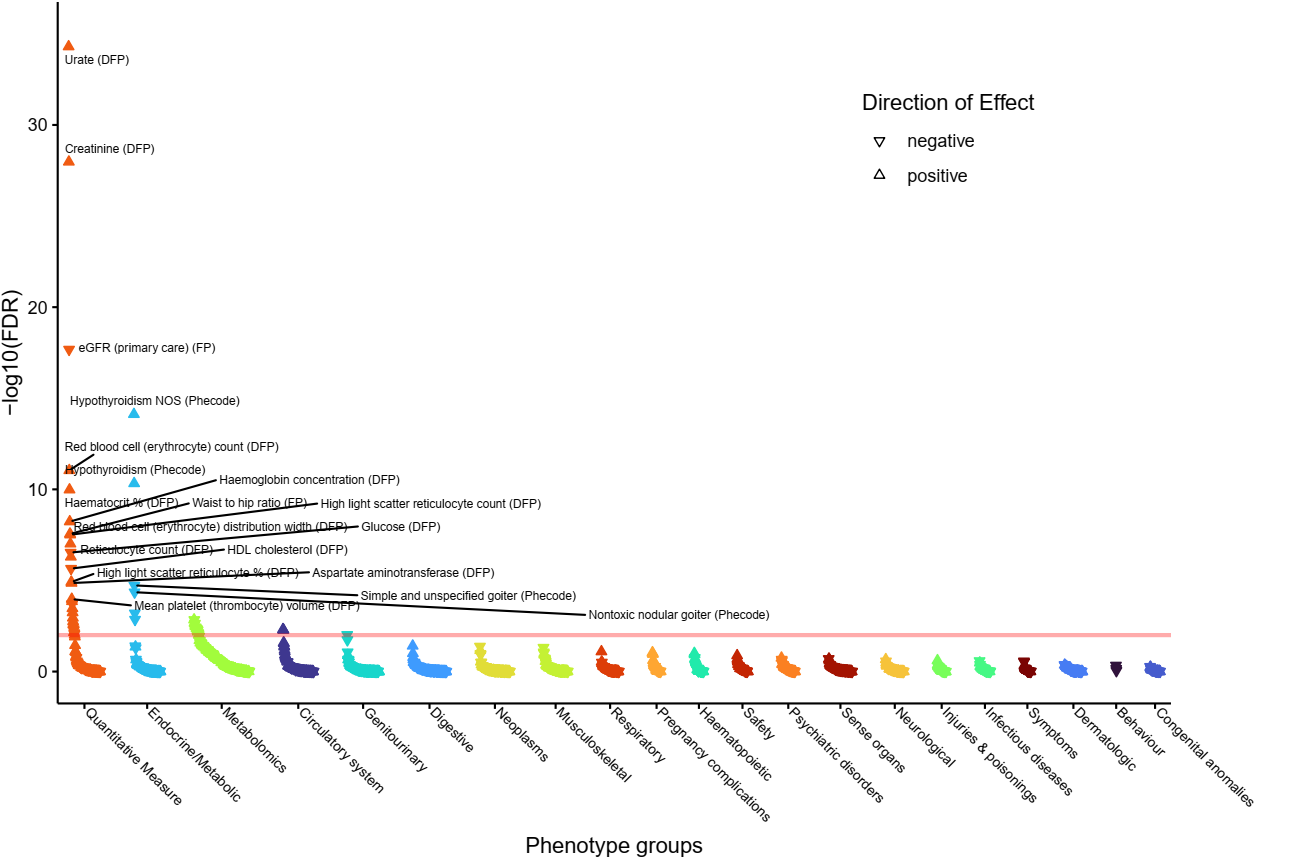

o) **Rap1 signaling pathway - Homo sapiens (human) pathway.**
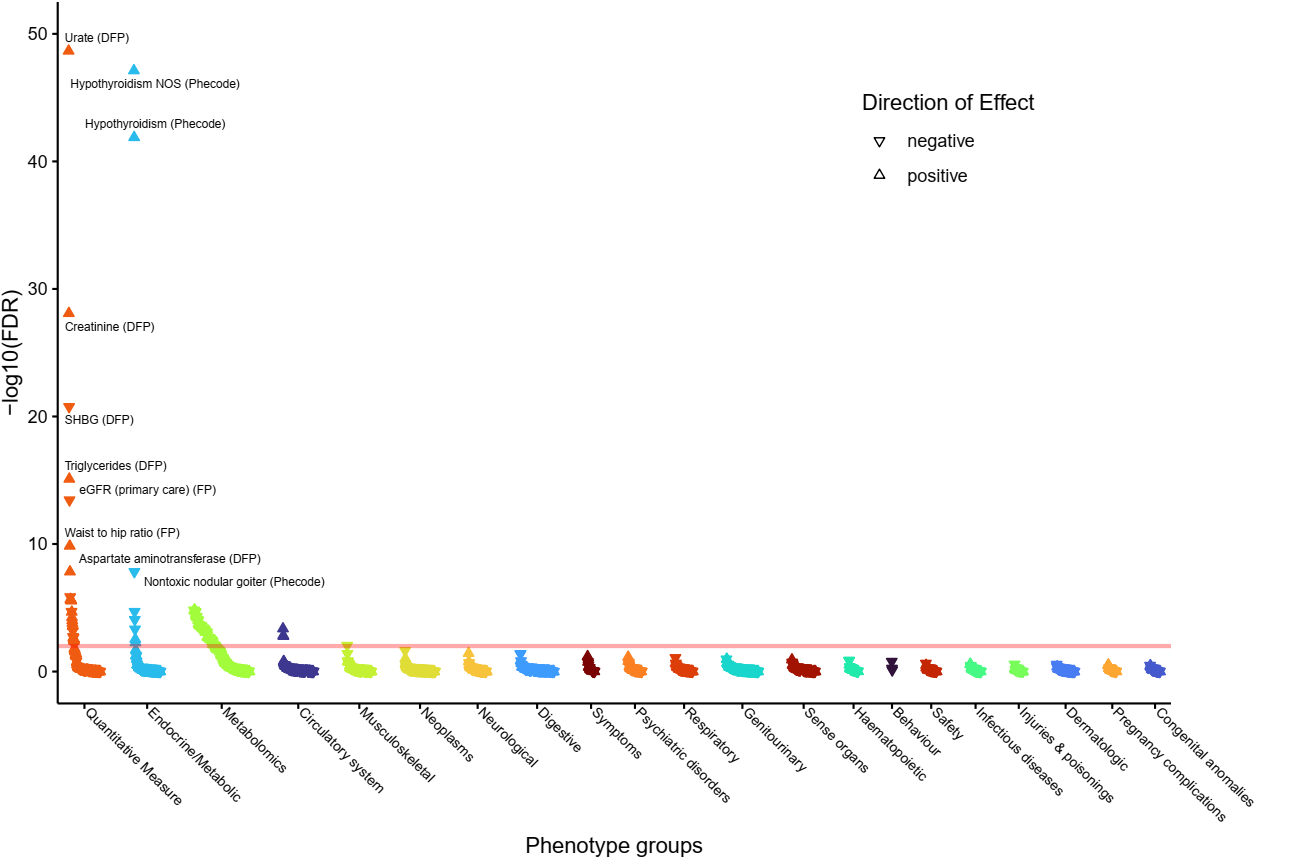

p) **Regulation of lipolysis in adipocytes - Homo sapiens (human) pathway.**
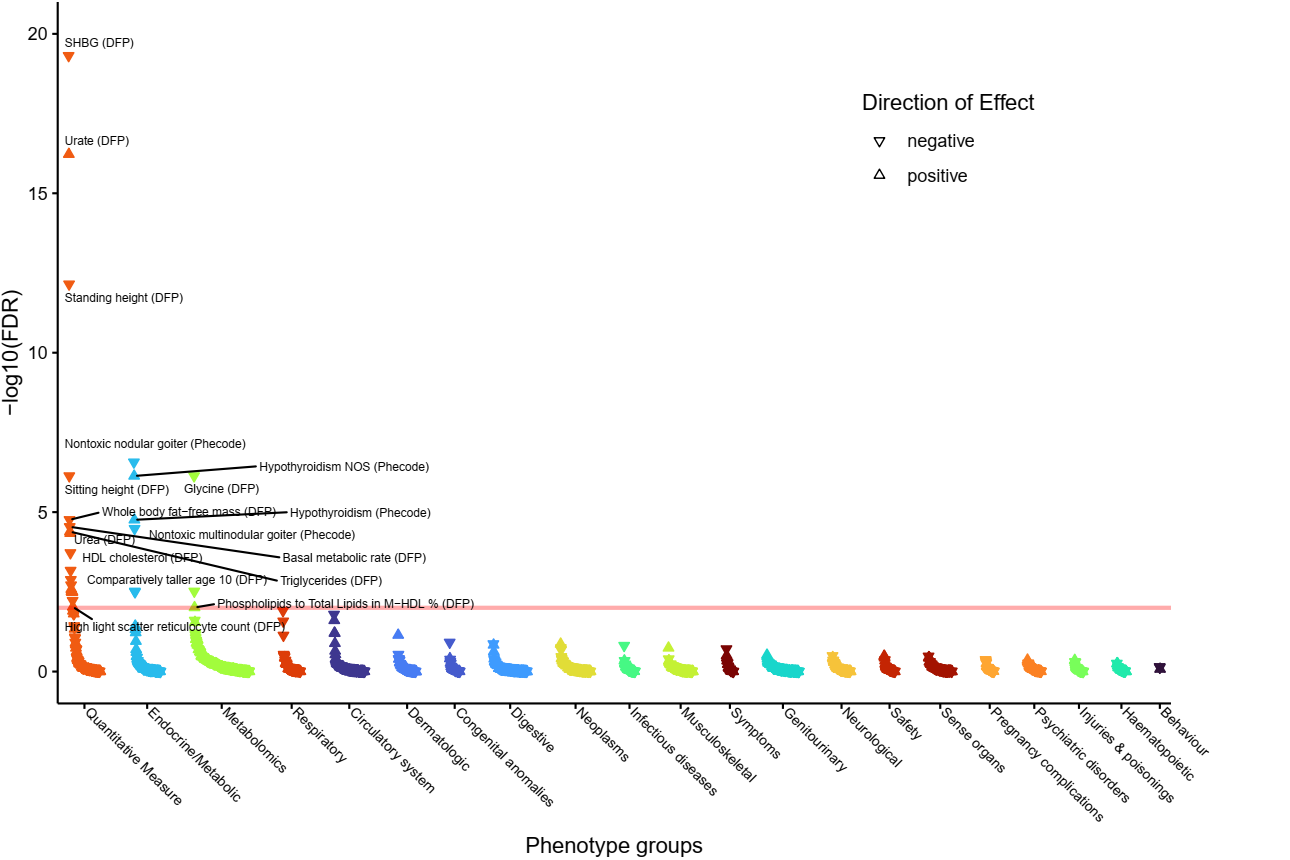

q) **Signal Transduction pathway.**

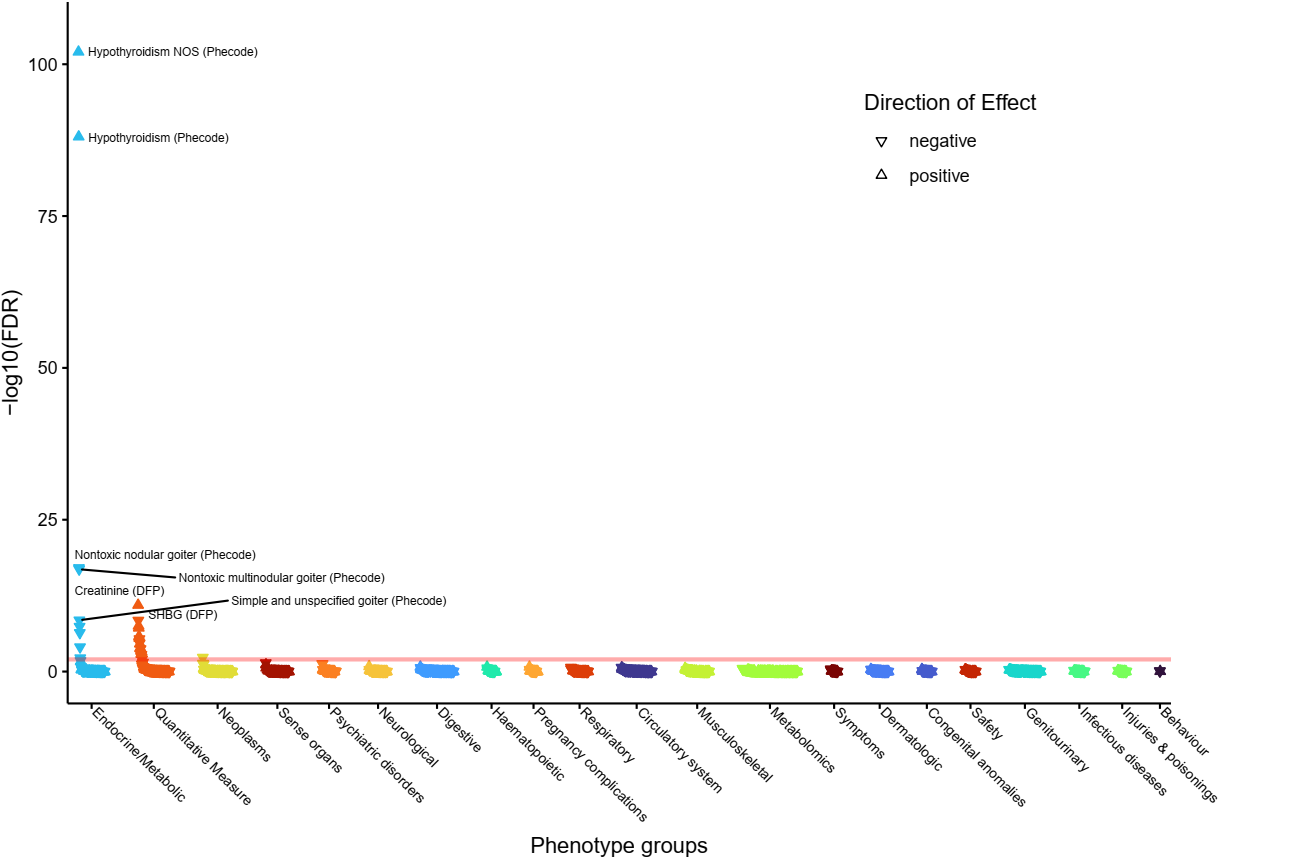

r) **Signalling by Receptor Tyrosine Kinases pathway.**
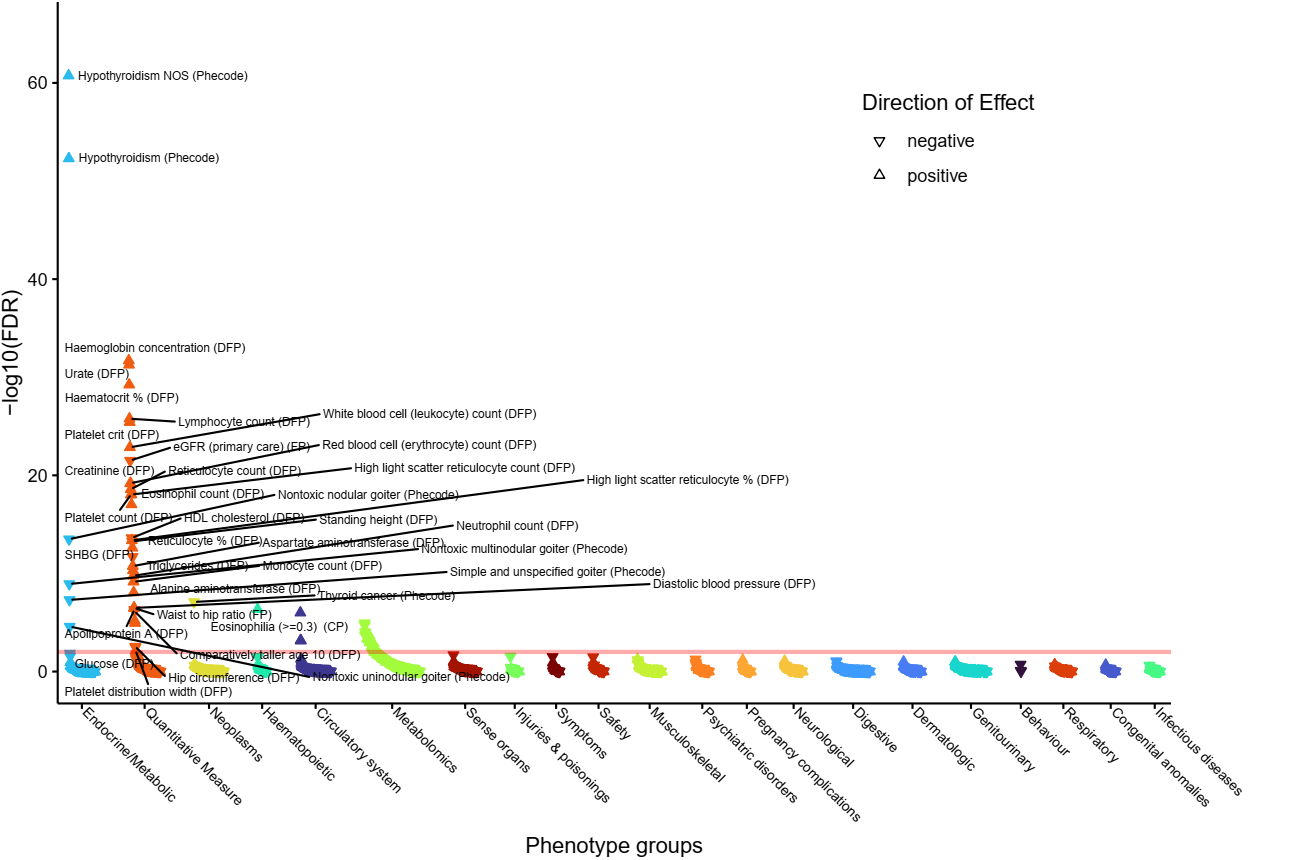

s) **Thyroxine (Thyroid Hormone) Production pathway.**
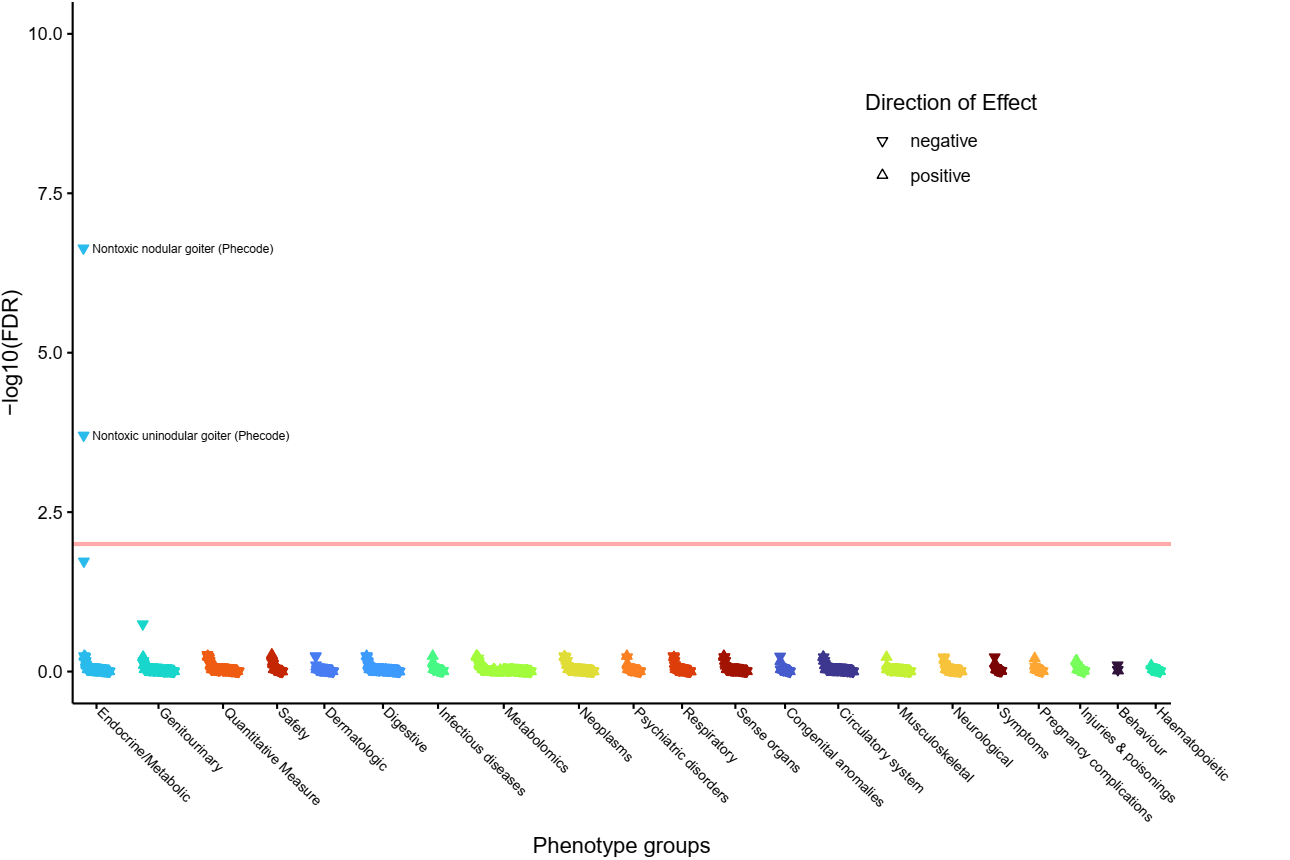

t) **Transcriptional regulation by RUNX2 pathway.**
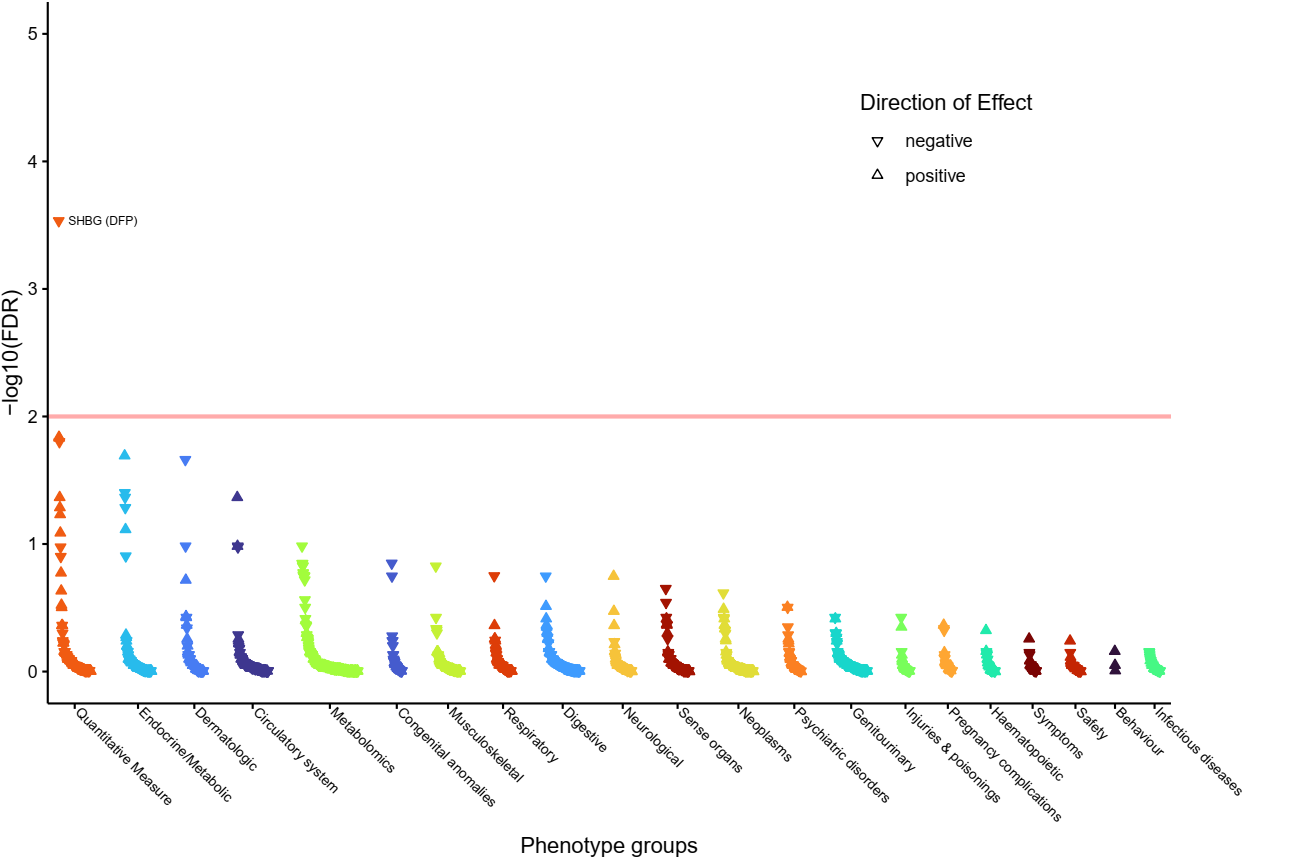

u) **Vegf hypoxia and angiogenesis pathway.**
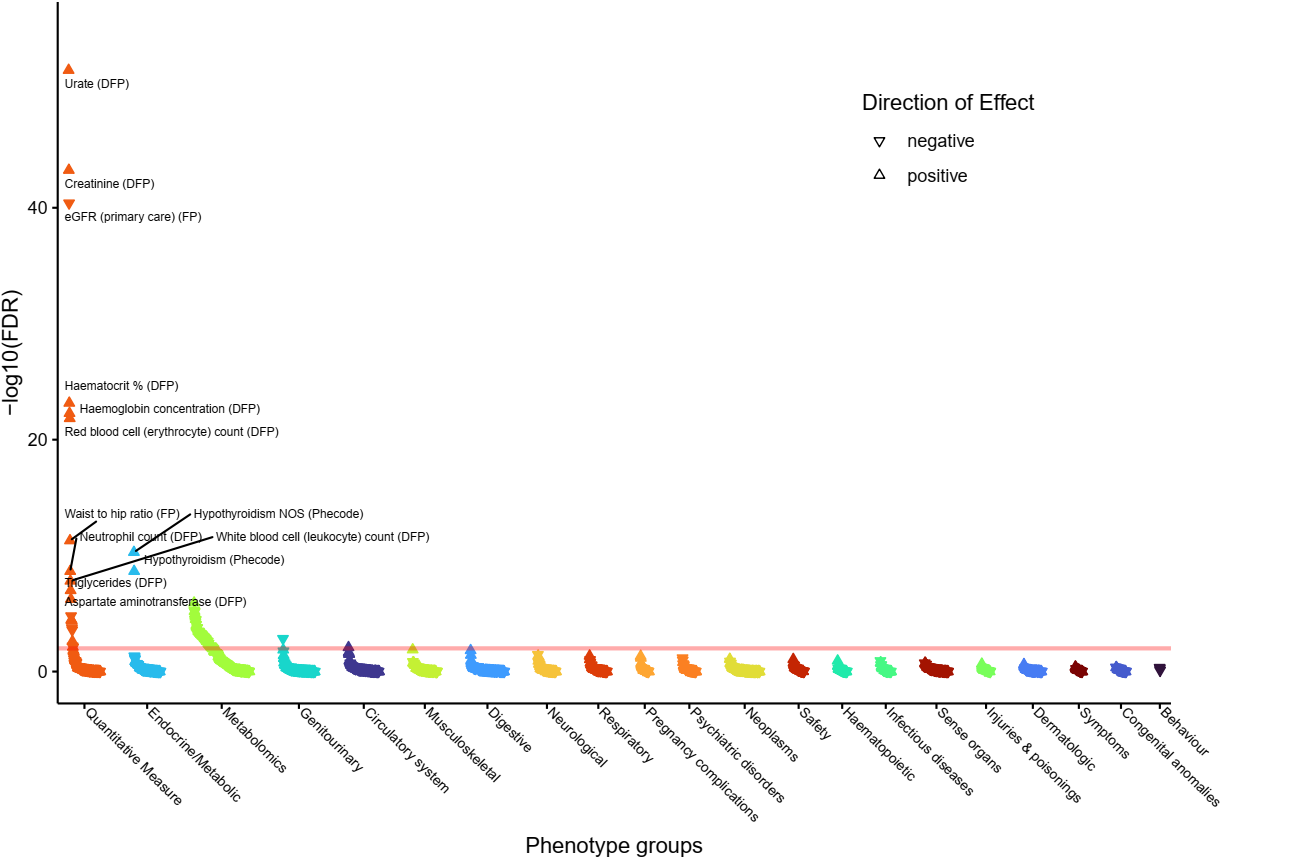

v) **VEGF ligand-receptor interactions pathway.**
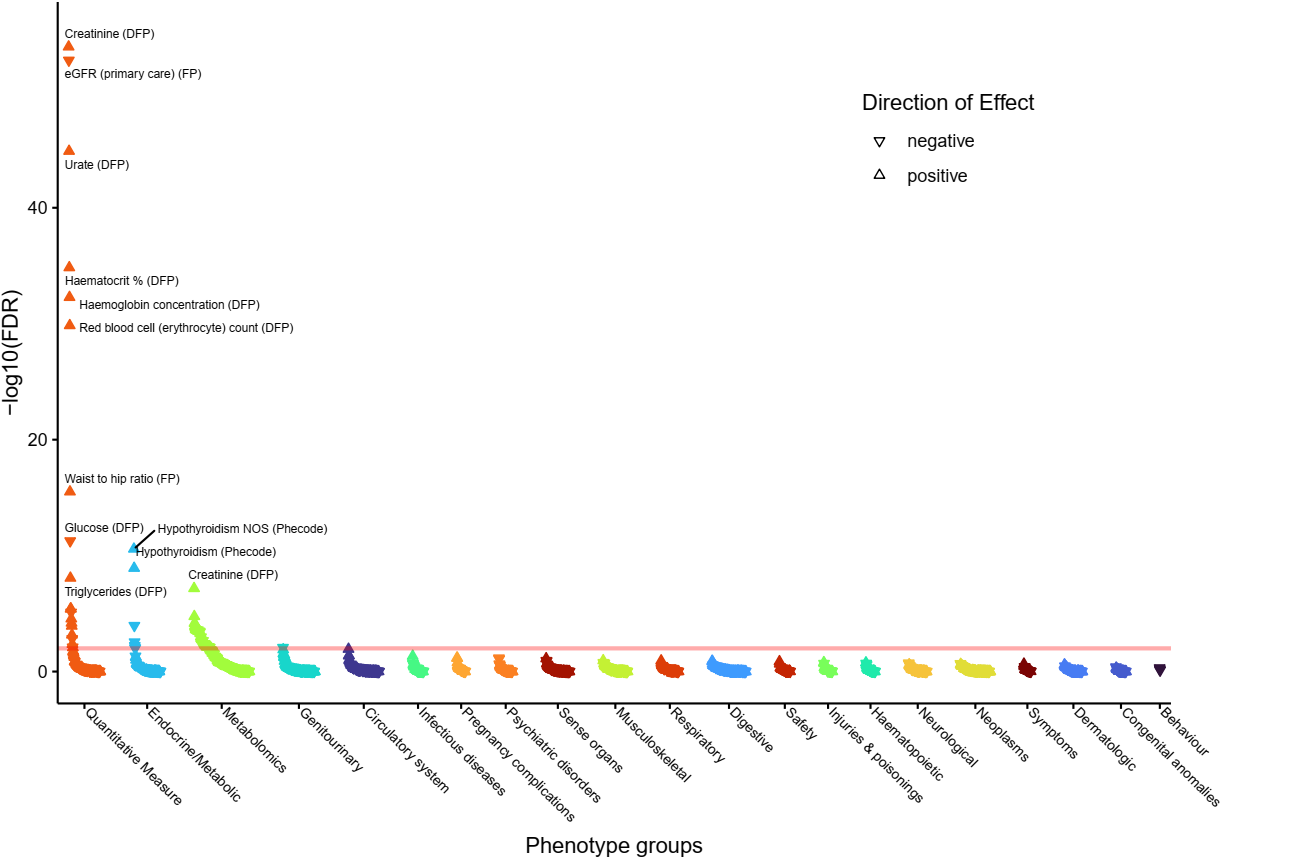

w) **VEGFA-VEGFR2 Signaling Pathway.**
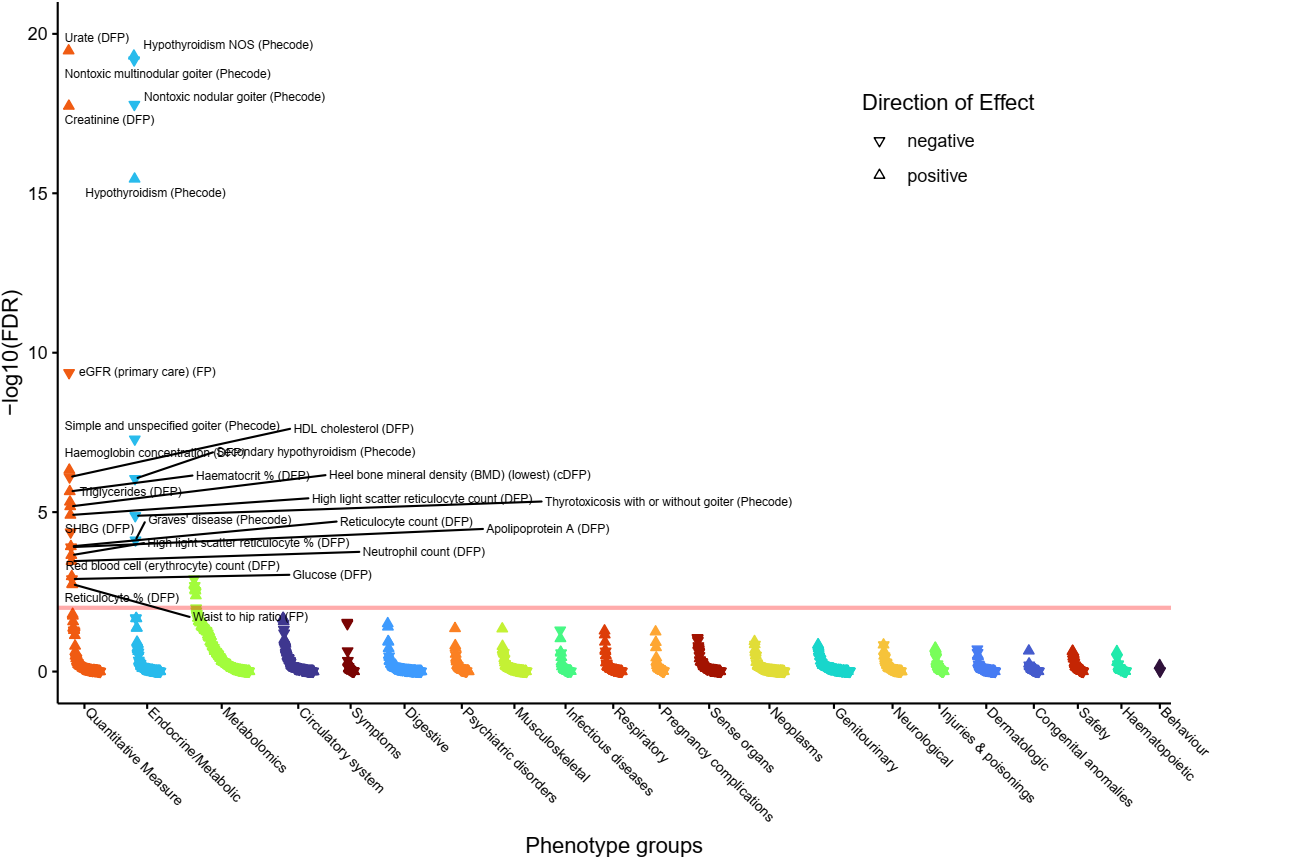

**Supplementary Figure 4**: Thyroid-stimulating hormone polygenic score PheWAS

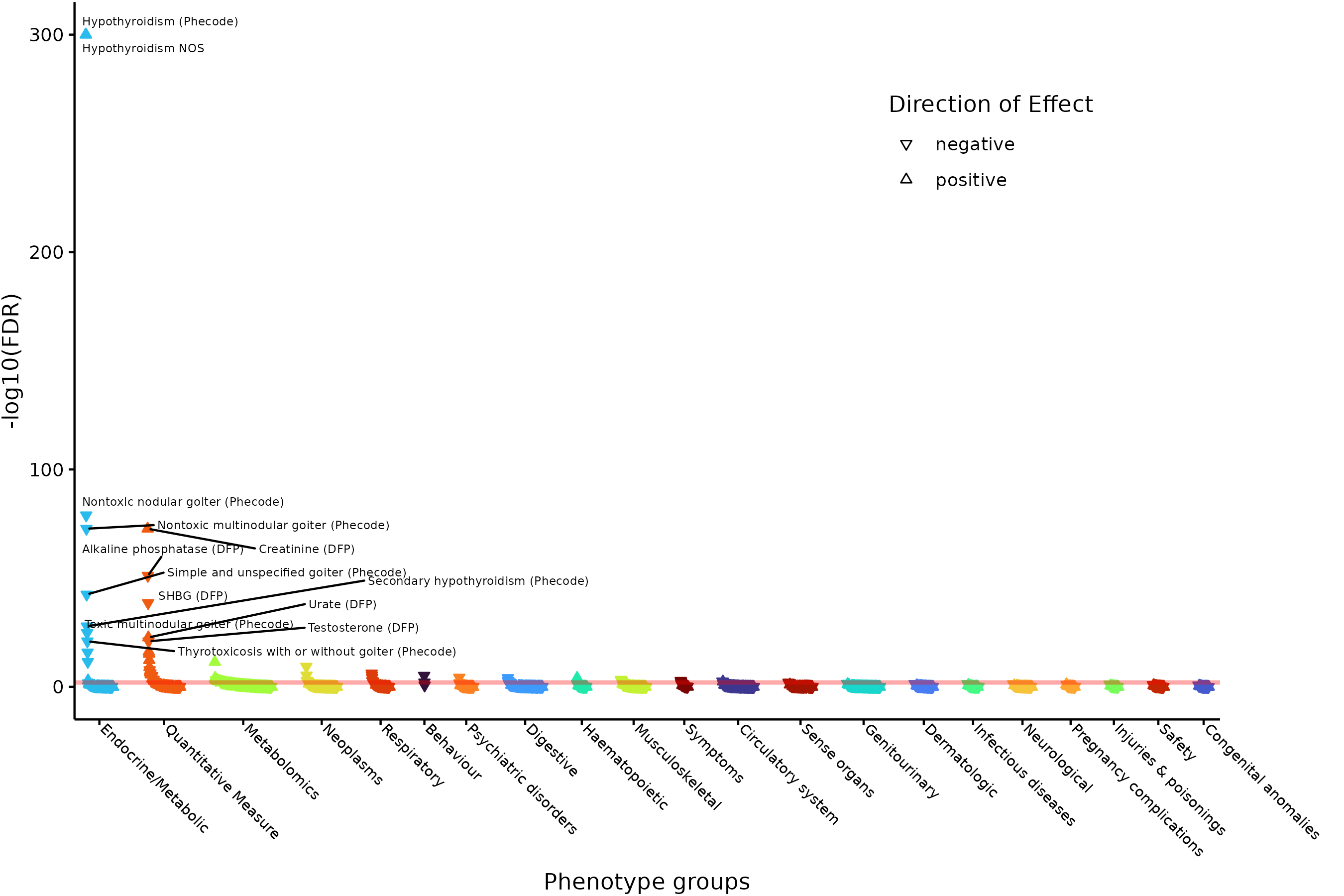

Supplementary Figure 5**: PGS performance across clinical diseases (sensitivity analysis free of overfitting).** Prediction performance of the TSH PGS for four clinical thyroid phenotypes – (**a, top left**) hypothyroidism, (**b, top right**) hyperthyroidism, **(c, bottom left**) thyroid cancer, and (**d, bottom right**) other thyroid disease. Error bars indicate 95% confidence intervals. The Mann-Kendall test is a test for monotonic trend.

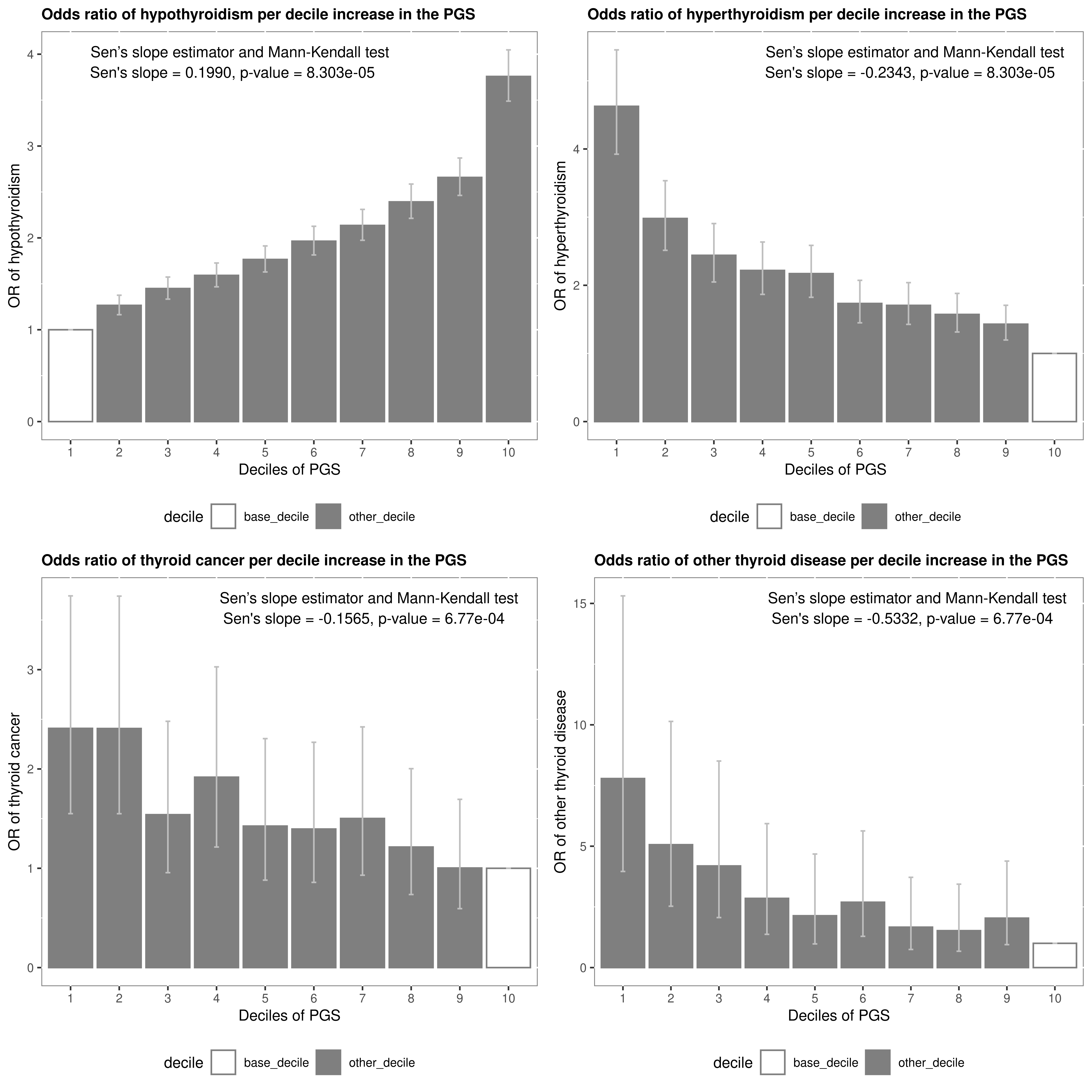

Supplementary Figure 6**: Age-of-onset analysis (sensitivity analysis free of overfitting).** Proportion of hypothyroidism (**a, left**) and hyperthyroidism (**b, right**) cases diagnosed by age stratified into lowest (grey), median (blue) and highest (yellow) decile for the TSH PGS. Shaded bands indicate 95% confidence intervals.

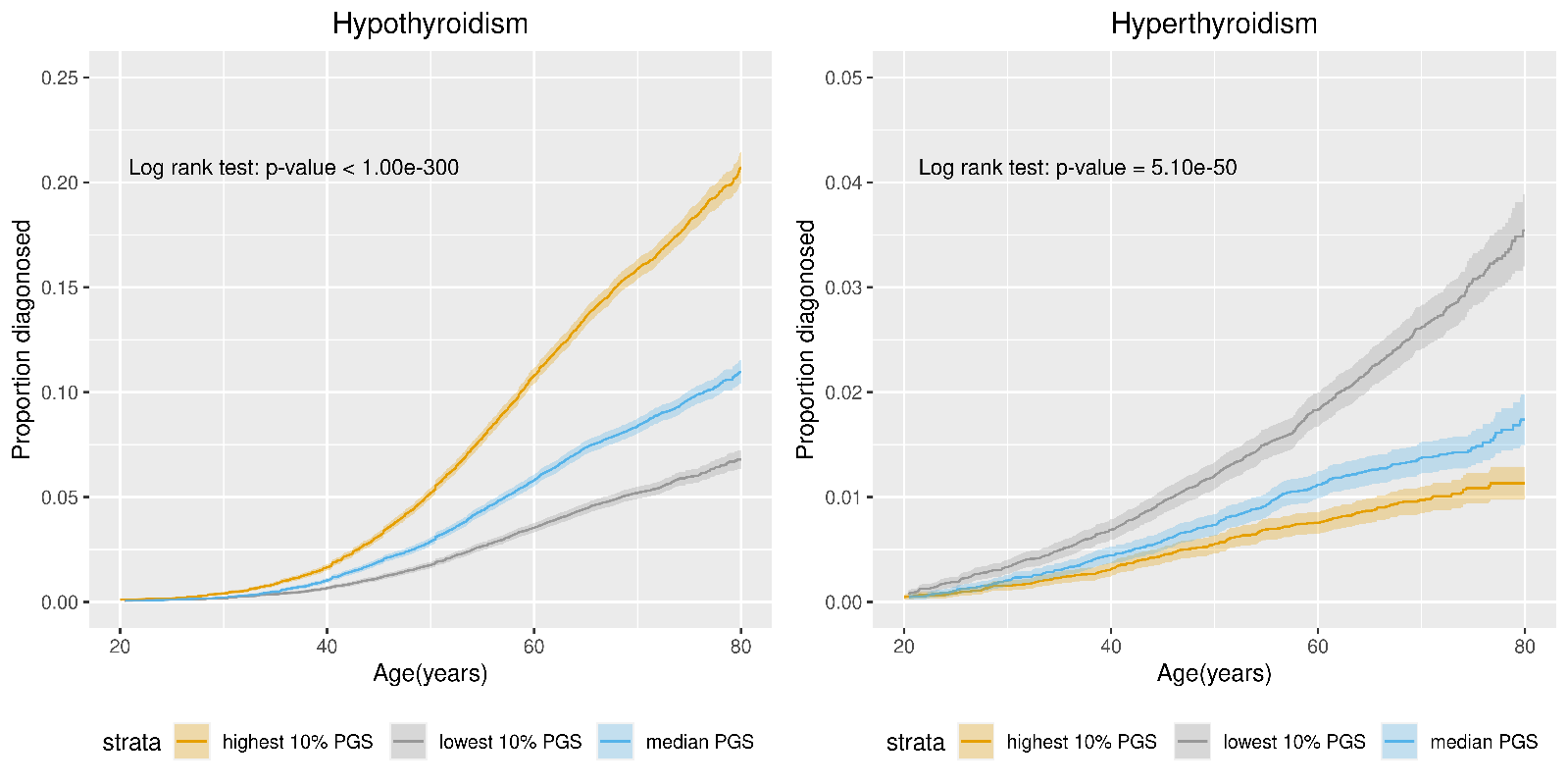

### Supplementary Tables

Supplementary Table 1: (Excel sheet) Signal selection results.
Each row represents the sentinel variant for an independent signal of association, the sentinel is the variant with the highest posterior probability (PIP column) returned from Polyfun Susie or Wakefield (method column). Allele1 is the coded/effect allele. The novel and novel locus columns indicated a previously unreported signal or locus respectively and the SNP reported, R2 and PMID columns give the previously reported SNP at the locus, it’s r^2^ with the signal reported here and the publication of the previously reported signal. The n_genes, genes and evidence columns give the genes implicated by the sentinel and the variant-to-gene evidence. Columns AA-AF give the VEP annotation for the sentinel and columns AG-AI give the most highly associated SNP in the locus. The study-level statistics from the 3 studies in the meta-analysis are in columns AJ-AR. The “consistent” column indicates signals that showed consistent direction of effect and P<0.01 in at least two of the three contributing datasets in addition to reaching P<5×10^-8^ in the meta-analysis.

Supplementary Table 2: (Excel sheet) Prioritised genes
List of prioritised genes according to the number of lines of variant-to-gene evidence implicates the gene (n_evidence column). The “Novel” column indicates whether the gene was previously reported. The n_novel_signals lists the number of novel signals implicating the gene.

Supplementary Table 3: (Excel sheet) Previously reported genes
Genes typically implicated by a single criterion and the reference for the previous publication reporting the gene.

Supplementary Table 4: (Excel sheet) Epidemiological associations
Epidemiological associations between the 257 top PIP variants available in UK Biobank and thyroxine (T4) levels or clinical thyroid diseases: hypothyroidism, hyperthyroidism, thyroid cancer, and other non-cancer thyroid diseases. Also shown are epidemiological associations with hypothyroidism due to medication defined using phecode 244.1 - these associations were only assessed in genetic variants implicating genes that met two or more variant-to-gene criteria.

Supplementary Table 5: (Excel sheet) Single-variant PheWAS associations
Phenome-wide association study (PheWAS) results for select TSH sentinel variants for associations at FDR <1% (European ancestry). N_ID = Number of participants in the analysis, FDR = false discovery rate, P = p value, OR = odds ratio, L95 = lower boundary of the 95% confidence interval, L95 = upper boundary of the 95% confidence interval, MAF = minor allele frequency, MAC = minor allele count, MAC_cases = minor allele count in cases, MAC_controls = minor allelecount in controls. Z_T_STAT = the Z or T STAT output from Plink2, SE = standard error. The P value is the P value from Plink2 for association for a linear model or logistic regression for the phenotype after adjusting for age, sex and first 10 principal components.

Supplementary Table 6: (Excel sheet) Variants selected at each gene for look up of epidemiological associations
For each gene the variant with the most significant association across either 6 or 7 binary traits is shown.

**Supplementary Table 7: (Excel sheet) Druggability**

Drug Gene Interaction Database (DGIDB) results. Columns:

- Drug; drug/compound name,
- ChEMBL_ID; drug/compound identification number from ChEMBL,
- Gene; mapped gene(s),
- Gene_source; line(s) of evidence for each mapped gene, and signal implicated (including the associated lung function trait),
- Indication(Phase); drug indication phase. Phase 1: Testing of drug on healthy volunteers for dose-ranging; Phase 2: Testing of drug on patients to assess efficacy and safety; Phase 3: Testing of drug on patients to assess efficacy, effectiveness and safety; and Phase 4: Approval of drug and post-marketing surveillance,
- Thyroid_related: whether the drug is used to for the treatment of thyroid conditions.
- Cancer_related: whether the drug is used to for the treatment of some form of cancer.

**Supplementary Table 8: (Excel sheet) ConsensuspathDB results**
ConsensuspathDB pathways enriched (FDR <5%) for genes from our 112 with at least 2 lines of evidence for being causal. The P value and FDR for enrichment of genes in pathways are those returned by ConsensuspathDB{Herwig, 2016 #78}.

**Supplementary Table 9: (Excel sheet) Pathway-based PheWAS results**
Results of 29 pathway-specific PheWAS for FDR <1%: FDR = false discovery rate, P = p value, OR = odds ratio, L95 = lower boundary of the 95% confidence interval, L95 = upper boundary of the 95% confidence interval.

**Supplementary Table 10: (Excel sheet) TSH polygenic risk score PheWAS results**
Results of TSH PGS PheWAS for FDR <1%: FDR = false discovery rate, P = p value, OR = odds ratio, L95 = lower boundary of the 95% confidence interval, L95 = upper boundary of the 95% confidence interval.

**Supplementary Table 11: (Excel sheet) Association results of TSH and free T4 with TSH PGS in ancestry groups in UK Biobank**

**Supplementary Table 12: (Excel sheet) Association results of diseases with TSH PGS in ancestry groups in UK Biobank**

**Supplementary Table 13: (Excel sheet) Association results of diseases with TSH PGS in ancestry groups in UK Biobank (winner’s curse free)**

**Supplementary Table 14: (Excel sheet) 95% credible sets**
All variants in the 95% credible sets are shown. The posterior probability (PIP column) returned from Polyfun Susie or Wakefield (method column). Allele1 is the coded/effect allele**.**

Supplementary Table 15: Clinical codes used to define thyroid-stimulating hormone (TSH), free T4, hypo- and hyperthyroidism, other thyroid diseases, and thyroid cancer.

| **TSH** | **T4** | **Hypothyroidism** | **Hyperthyroidism** | **Other thyroid diseases** | **Thyroid cancer** |
| --- | --- | --- | --- | --- | --- |
| 442..  442A.  442A0  442A1  442e.  442K.  442L.  442M.  442N.  442O.  442P.  442Q.  442R.  442S.  442T.  442W.  442X.  X80Gc  XaBj2  XaELV  XaELW  XaESa  XaESb  XaESc  XaESX  XaESY  XaESZ  XaET1  XaET2  XaET7  XaETX  XaIzE  XE27u  XE2wy | 4427.  442V.  442c.  4426.  XaERr  XaERs | C04..  C0430  C0431  C0432  C043z  C044.  C046.  C047.  C04y.  C04z.  C0A5.  Cyu11  PK251  X40HD  X40HE  X40HF  X40HG  X40HH  X40HI  X40HL  X40HM  X40HN  X40HO  X40HP  X40Hv  X40Hw  X40Hx  X40Hy  X40Hz  X40IB  X40IQ  X40OP  X50Fa  XE108  XE10A  XE124  Xa08g  Xa3ec  Xa3ed  XaJ9F | C02..  C020.  C0200  C0201  C020z  C021.  C0210  C0211  C021z  C022.  C0220  C0221  C022z  C023.  C0230  C0231  C023z  C024.  C0240  C0241  C024z  C025.  C02y.  C02y0  C02y1  C02y2  C02yz  C02z.  C02z0  C02z1  C02zz  C05y4  Cyu13  X40Gj  X40Gk  X40Gl  X40Gn  X40Go  X40Gt  X40Gv  X40Gw  X40Gx  X40Gy  X40Gz  X40H0  X40H1  X40H2  X40H4  X40H5  X40H7  X40Hd  X40I4  X40OI  X40OK  X40OO  XE104  XE105  XE106  XE122  XE1g7  Xa09v  Xa3eb  XaZtG | C0...  C00..  C000.  C00z.  C01..  C010.  C011.  C01z.  C05..  C050.  C0500  C0501  C050z  C051.  C052.  C053.  C054.  C05y.  C05z.  C061.  C0A3.  C0A4.  C0AX.  Cyu12  Cyu14  Cyu16  L1810  L1811  L1812  L1813  L1814  L1815  L181z  X40Dj  X40Ge  X40Gf  X40Gi  X40HR  X40HS  X40HT  X40HU  X40HV  X40HW  X40HY  X40HZ  X40Hc  X40He  X40Hf  X40Hg  X40Hh  X40Hu  X40IE  X40IG  X40II  X40IJ  X40OR  X76FB  XE101  XE102  XE103  XE10B  XE10C  XE10D  XE120  XE126  XM0Ap  Xa08f  Xa3ea | C73  193  V1087 |

**Supplementary Table 16: (Excel sheet) FUMA eQTL look up results**
Look up of eQTL results from the GTEx v8 (thyroid, hypothalamus and pituitary tissues) and eQTLGen (blood, cis- and trans-eQTLs) datasets for variants in our 95% credible sets.

**Supplementary Table 17: (Excel sheet) Polygenic Priority Score (PoPS) results - 500KB window**
Polygenic Priority Score (PoPS) results using a +/-250KB window

**Supplementary Table 18: (Excel sheet) Genes near our signals associated with rare Mendelian respiratory diseases**
34 genes associated with a rare Mendelian respiratory disease were implicated.

**Supplementary Table 19: (Excel sheet) Mouse ortholog genes near our signals associated with a respiratory disease**
6 genes were implicated

**Supplementary Table 20: (Excel sheet) Look up of UK Biobank WES variants**
Look up of UK Biobank WES variants within +/-500kb of TSH sentinels.

**Supplementary Table 21: (Excel sheet) Putative causal variants**
Variants from credible sets annotated as missense/damaging/deleterious and with a PIP>50%
